# From naive to foundation: benchmarking models for epidemic forecasting

**DOI:** 10.64898/2026.05.11.26352889

**Authors:** Douhan Wang, Yuhan Li, Nicola Perra

## Abstract

We systematically evaluate and compare the performance of classical statistical methods (ARIMA), mechanistic compartmental models (SEIR), modern deep learning architectures (LSTM, DLinear, Autoformer), and an emerging time-series foundation model (TabPFN-TS) to forecasts the incidence of Influenza-Like Illness (ILI) across nine European countries. The models are benchmarked against a naive baseline and a multi-model ensemble (RespiCast) created by an initiative of the ECDC. In line with the operational practice of existing forecasting hubs, our entire evaluation is explicitly optimized for short-term horizons (1 to 4 weeks ahead). Interestingly, we found that the foundation model TabPFN-TS allows for great zero-shot inference capabilities. Without any task-specific retraining, it successfully overcomes extreme data scarcity to consistently outperform all other individual architectures, frequently rivalling or surpassing the RespiCast ensemble. Our results highlight how deep learning architectures are severely constrained by extreme data scarcity, typical in epidemic forecasting, requiring targeted endogenous data augmentation to reduce predictive errors. Within the deep learning class of models, we observe that simpler architectures (such as DLinear and LSTM) frequently exhibit greater robustness and outperform complex, attention-based models (such as Autoformer) when data is constrained. Finally, our results show how a weighted ensemble, constructed by fusing all the models, delivers highly robust forecasts in all regions considered. Overall, our findings showcase the transformative potential of zero-shot foundation models in epidemic forecasting and confirm the importance of multi-model ensembles.

## Introduction

Epidemic forecasting is an important challenge in public health and epidemiology [1, 2]. Seasonal influenza alone leads to tens of millions of symptomatic cases and tens of thousands deaths annually in the European Union and European Economic Area [3, 4]. Accurate forecasts of key epidemic indicators, such as incidence of cases and hospitalizations, are essential for informing public health responses, including healthcare resource allocation, early warning systems, and intervention planning [5, 6]. Beyond operational utility, forecasts provide valuable insights into the variability of disease transmission dynamics across populations and time, thus contributing to a broader understanding of epidemic processes [7].

Mechanistic compartmental models, such as the susceptible-infectious-recovered framework, have been widely used for scenario analysis, policy evaluation, and forecasts [8–10]. These approaches describe transmission dynamics at different levels of abstractions modelling the transitions of individuals across various compartments [8, 10]. In recent times, another class of models, based instead on statistical and pattern recognition methods, has gained popularity within the epidemic forecasting community. Here we find traditional statistical frameworks for time-series like the ARIMA model as well as machine learning approaches based on deep learning (e.g., recurrent neural networks and transformers) [11–15]. Popular architectures such as Informer, Autoformer, and FEDformer have been specifically designed to address long-term time-series forecasting challenges [16–18]. More recently, foundation models for time-series forecasting, including Chronos, TimesFM, and TabPFN-TS, have demonstrated strong performance across diverse benchmark datasets, particularly in zero-shot or low-data settings [19–22]. In the context of machine learning, *zero-shot inference* refers to a model’s capacity to generate accurate predictions for a novel task or domain without undergoing any task-specific retraining or parameter fine-tuning. Unlike classical statistical methods or traditional deep learning architectures that require iterative weight optimization on historical observations to learn localized patterns, a zero-shot model relies entirely on a vast, task-agnostic pre-training phase. By pre-training on massive and heterogeneous collections of time-series, the model learns generalized temporal dynamics. Consequently, during the forecasting phase, the model’s internal parameters remain strictly frozen. It evaluates new epidemiological data and approximates the future predictive distribution in a single forward pass, which effectively bypasses the severe data scarcity limitations that typically cause traditional neural networks to over-fit [23]. In operational forecasting practice, ensemble approaches have become the dominant paradigm. In both the United States and Europe institutions such as the Centers for Disease Control and Prevention (CDC) and the European Center for Disease Control and Prevention (ECDC) routinely aggregate predictions from multiple models to improve robustness and accuracy [24–26].

Despite rapid methodological advances, the evaluation of forecasting models remains limited in several respects. First, existing benchmarks are often misaligned with the requirements of operational epidemiology. Many forecasting models, especially those based on deep learning, are optimized for long-horizon prediction accuracy on large-scale, generic datasets [27, 28], whereas, due to the nature of epidemic dynamics and human behaviour, short-term forecasts (typically 1 − 4 weeks ahead) are the standard in epidemiology [24–26, 29]. This mismatch can lead to model selection criteria that do not translate effectively to real-world deployment. Second, there are only few systematic evaluations of forecasting performances across model classes [22, 30–33]. More work is needed to draw generalizable conclusions about the relative strengths and limitations of different modelling paradigms.

In this study, we evaluate a range of forecasting approaches for epidemic prediction. By using weekly Influenza-Like Illness (ILI) data from nine European countries as target, we benchmark classical statistical models (ARIMA [34]), mechanistic compartmental models (SEIR [8, 35]), deep learning architectures (LSTM [36], DLinear [37], Autoformer [17]), and foundation models (TabPFN-TS [21]). We sample different contexts and epidemic variabilities by selecting Belgium, Czechia, Denmark, France, Ireland, Italy, Netherlands, Poland, and Romania. The models are evaluated alongside a naive baseline, the RespiCast ensemble (the ECDC multi-model forecasting hub [38]), and weighted ensemble, built by mixing all models, calibrated on historical probabilistic performance. We incorporate an empirical residual calibration strategy inspired by conformal prediction principles [39]. Additionally, we investigate strategies to address the data scarcity limitations commonly encountered when using data hungry models. In doing so, we introduce two complementary data expansion approaches: (i) a statistical augmentation strategy that generates synthetic time-series by shifting epidemic peaks and incorporating observational noise, thereby mimicking variability in epidemic dynamics and reporting processes; and (ii) a mechanistic augmentation strategy that incorporates simulated trajectories derived from an SEIR model calibrated to real data.

Our analysis reveals three insights for operational forecasting. First, the foundation model TabPFN-TS demonstrates exceptional zero-shot capabilities. Without any task-specific retraining, it successfully overcomes extreme data scarcity and consistently outperforms individual deep learning models and frequently rival the RespiCast ensemble. Second, we find that while neural networks are severely constrained by limited data, their predictive stability is substantially improved by targeted endogenous data augmentation. Finally, our performance-calibrated weighted ensemble effectively leverages these diverse architectural biases, delivering highly robust probabilistic forecasts across extended horizons and mitigating the volatility of individual models.

Overall, our analysis provides a systematic comparison of forecasting paradigms and offers practical insights into the conditions under which statistical, deep learning, and foundation models perform effectively, while also contextualizing their performance relative to ensemble systems that incorporate mechanistic modelling components.

## Results

In what follows, we first detail the taxonomy of the models considered. We then define the forecasting targets, outline the strategies employed for fitting and generating forecasts. Following this methodological setup, we present the comparative performance of the models, evaluating both their point forecasting accuracy and probabilistic reliability.

### Models’ taxonomy and baselines

To systematically evaluate forecasting performance across different methodological paradigms, we select a diverse set of models. As summarized in Table 1, these range from classical statistical methods and widely-used deep learning architectures to emerging foundation models and mechanistic epidemic models. Additionally, we included a naive baseline and the RespiCast ensemble as structural benchmark. The naive model consistently predicts the future by using the last observed value. As mentioned, RespiCast is a robust European multi-model operational forecasting system that integrates mechanistic, statistical, and machine learning approaches [38]. The classical statistical forecasting model we use is the AutoRegressive Integrated Moving Average (ARIMA) model [34], a widely used method for time-series forecasting. We included an age-stratified SEIR (Susceptible-Exposed-Infectious-Recovered) model with vaccinations calibrated via Approximate Bayesian Approximation methods as example of mechanistic epidemic frameworks [40, 41]. To represent deep learning methods, we evaluate three distinct architectures: Long Short-Term Memory (LSTM) networks [36], which are recurrent neural networks capable of capturing long-term temporal dependencies in sequence data; DLinear, a streamlined linear model optimized for time-series tasks via series decomposition into trend and remainder components [37]; Autoformer, a Transformer-based architecture featuring an auto-correlation mechanism designed to discover period-based temporal dependencies [17]. Finally, we included TabPFN-TS to represent emerging foundation models [21]. This pre-trained prior-data fitted model enables effective zero-shot forecasting without the need for task-specific training. To capitalize on the diverse biases of these paradigms, we also developed a performance-calibrated weighted ensemble. This integrative approach dynamically aggregates the predictions of the individual models by assigning weights based on their historical probabilistic accuracy (*IS*_80_). We refer the reader to the Materials and Methods as well as the Supplementary Information for more details about each model.

**Table 1:** Taxonomy and brief description of the forecasting models evaluated in this study.

| Model | Category | Short Description |
| --- | --- | --- |
| **Naive** | Baseline | Predicts future incidence exactly as the last observed value. |
| **RespiCast** | Benchmark | A robust European multi-model operational forecasting system integrating mechanistic, statistical, and machine learning approaches. |
| **ARIMA** | Statistical | Classical autoregressive integrated moving average model, widely used for linear time-series forecasting. |
| **SEIR** | Mechanistic | A mechanistic age-stratified compartmental model. |
| **LSTM** | Deep Learning | Recurrent neural network architecture. |
| **DLinear** | Deep Learning | A streamlined linear model with series decomposition (trend and remainder), optimized for time-series tasks. |
| **Autoformer** | Deep Learning | Transformer-based architecture featuring an auto-correlation mechanism for discovering period-based temporal dependencies. |
| **TabPFN-TS** | Foundation Model | Pre-trained prior-data fitted model enabling effective zero-shot forecasting without the need for task-specific training. |
| **Weighted ensemble** | Ensemble | A performance-calibrated aggregation that weights constituent models based on their historical probabilistic reliability ($IS_{80}$). |

### Data and forecasting targets

The forecasting target in our evaluation is the official weekly incidence of Influenza-Like Illness (ILI). We use retrospective surveillance data from nine European countries: Belgium, Czechia, Denmark, France, Ireland, Italy, Netherlands, Poland, and Romania. These countries provide a geographically diverse set of epidemic dynamics and enable comparison with the RespiCast ensemble.

We consider data from the 2017 − 2018, 2018 − 2019, 2023 − 2024, and 2024 − 2025 influenza seasons. The last season is used a target of our forecasts and the previous three for training. Each season is defined from ISO week 42 to week 14 of the following year, capturing the primary epidemic period. Seasons during the COVID-19 pandemic are excluded due to substantial disruptions to typical influenza transmission patterns, which introduce structural shifts in the time series.

### Model training and data augmentation

Different model architectures require distinct training protocols. Classical statistical models (e.g., ARIMA) are fitted directly to the historical data immediately preceding the forecasting window. These require no formal train-test partitioning. The SEIR model likewise bypasses conventional neural network training, and its parameters are calibrated via Approximate Bayesian Computation (ABC) [40]. Similarly, the foundation model, TabPFN-TS, is applied zero-shot. The predictions rely entirely on its task-agnostic pre-training, without any task-specific training.

Deep learning architectures (e.g., LSTM, DLinear, and Autoformer) are data-intensive. In standard neural network pipelines, data is rigorously partitioned into three distinct subsets: a training set to iteratively learn patterns and optimize internal weights, a validation set to tune hyper-parameters and prevent over-fitting (e.g., via early stopping), and a strictly sequestered test set for the final out-of-sample evaluation. Applying this standard protocol to seasonal epidemic forecasting, however, presents a critical challenge. A typical influenza season yields fewer than 30 observational data points. Due to this severe data scarcity, reserving a separate validation set is not feasible when evaluating models strictly on surveillance data, as doing so will critically deplete the already limited training pool. Therefore, these models are trained directly on the three available historical seasons (i.e., 2017 − 2018, 2018 − 2019, and 2023 − 2024), with the 2024 − 2025 season serving as the target for forecasting evaluation.

Relying solely on limited historical data makes neural networks highly susceptible to over-fitting. To alleviate this constraint and systematically investigate the impact of different data generation paradigms, we developed a data augmentation pipeline that enriches the training space. We generated two distinct streams of synthetic time series, each containing 1, 000 trajectories per season. While we refer the reader to the Material and Methods as well as to the Supplementary Information for more details, here we provide a high level summary of the two methods:

- **Endogenous Augmentation:** We generate 1, 000 synthetic trajectories per season by applying temporal translations to shift epidemic peaks and injecting realistic observational noise.
- **Exogenous Augmentation:** We calibrate the a SEIR model to the three seasons’ data separately (i.e., 2017 − 2018, 2018 − 2019, and 2023 − 2024) using the ABC-SMC (Approximate Bayesian Computation with Sequential Monte Carlo) algorithm [41]. For each season, we accept 1000 trajectories per season.

With the training space sufficiently enriched by either augmentation strategy, we restructured our dataset split to enable proper model validation and prevent data leakage. For these augmented experiments, deep learning models are trained on datasets comprising real data from the 2017 − 2018 and 2018 − 2019 seasons alongside the respective 1, 000 synthetic time-series (either endogenous or exogenous). To distinguish these configurations in our subsequent evaluation, we denote models trained on real data plus endogenous augmentation as the augmented (aug) setting, and those trained on real data plus exogenous augmentation as the combined (comb) setting. Crucially, the real data for 2023−2024 season is strictly reserved as a validation set for hyper-parameter tuning and early stopping, while the 2024 − 2025 season remains the unseen target for final forecasting evaluation.

### Forecasting setup and evaluation metrics

To simulate real-time operational forecasting settings, we implement a rolling-window strategy during the evaluation phase. We set the start of forecasts at ISO week 45 of the target season. For each forecasting week *w*, all models generate a 1 to 4 week-ahead forecasts. While the forecasting horizon is the same for all models, the historical look-back window used for inference varies according to model type. Specifically, the deep learning architectures (e.g., LSTM, DLinear, and Autoformer) employ a strictly fixed 4-week sliding window as their input sequence. In contrast, models relying on dynamic parameter calibration or in-context learning (i.e., ARIMA, SEIR model, and TabPFN-TS) use the cumulative observed data including the entire multi-season historical data up to the forecasting week *w*. The observation window is then sequentially advanced by one week, incorporating newly observed data, and the process is repeated until the end of the epidemic season.

To assess both point and probabilistic forecast accuracy, we evaluated models’ performance using three complementary metrics:

- **Mean Absolute Error (MAE):** Evaluates absolute point forecast accuracy.
- **Weighted Mean Absolute Percentage Error (WMAPE):** Provides a scale-independent measure of relative accuracy. This is essential for our multi-country framework, enabling equitable performance comparisons across regions with varying population sizes and baseline incidences.
- 80% **Interval Score (***IS*_80_): Evaluates probabilistic forecasting performance specifically for the 80% prediction interval. It assesses the quality of the predicted bounds by simultaneously penalizing excessively wide intervals (i.e., lack of precision) and instances where the true observation falls outside the interval (i.e., coverage failure). This balanced uncertainty quantification is critical for risk-aware public health decision-making.

### Point and probabilistic forecasting accuracy

To assess the predictive capabilities of the evaluated architectures, we focus the discussion that follows on the 4-week-ahead forecasts (horizon 4) which, being the farthest with respect to the last known data point, are the most challenging. We report the details of other horizons (i.e., 1, 2, 3) in the Supplementary Information.

We first evaluate probabilistic forecasting performance, using the Relative 80% Interval Score (Relative *IS*_80_) compared against the naive model which is used as baseline (see Fig. 1). In the figure, models with scores smaller than one outperform the baseline. We also report the mean *IS*_80_ values for all forecasting horizons in Table 2. The results reveal a pronounced disparity across models, and some clear trends in terms of performance. Interestingly, the RespiCast ensemble is confirmed as a strong benchmark, delivering robust and superior probabilistic forecasts in 6 out of the 9 evaluated countries. At the same time, the foundation model, TabPFN-TS, demonstrates great zero-shot probabilistic accuracy that rivals or even surpasses the RespiCast ensemble. Across all the countries except for Romania, its median Relative *IS*_80_ falls below 1.0, indicating substantial improvements over the naive model. For instance, TabPFN-TS demonstrated great forecasting stability in Ireland and Italy, achieving the lowest mean *IS*_80_. Specifically, Table 2 shows a mean *IS*_80_ of 41.2 for Ireland and 869.6 for Italy at horizon 4, significantly outperforming their respective naive baseline (155.7 and 1391) as well as the RespiCast ensemble (55.49 and 1405). The SEIR model exhibits high variability in terms of performance. While it exceeds the naive benchmark in Belgium (1118 vs 2164) and France (476.2 vs 783.3), it struggles in the others where it largely fails to outperform the naive baseline’s *IS*_80_. Similarly, ARIMA demonstrates mixed performance. It improves upon the naive baseline in several settings, such as France at horizon 4 (600.5 vs. 783.3) and the Netherlands (126.8 vs. 171.8). However, this advantage is not consistent across countries nor horizons. For example, ARIMA performs worse than the naive baseline in Romania at horizon 4 (110.3 vs. 69), and also shows limited gains in some short-horizon cases, such as Belgium at horizons 1–2 (663.9 vs. 653 and 1171 vs. 1132). This pattern is consistent with the relative IS results, where ARIMA often remains close to the naive baseline rather than consistently shifting below it. In contrast, the deep learning architectures (i.e., DLinear, LSTM, Autoformer) trained exclusively on real data exhibit high variance and frequent underperformance, often failing to beat the naive baseline. This limitation is common across all models in this category. However, the introduction of the data augmentation pipeline notably stabilizes these architectures and significantly reduces absolute prediction errors. As visually indicated by the left-ward shift and narrowed variance in the relative *IS*_80_ distributions, this stabilization translates into measurable performance gains. This improvement is explicitly supported by the quantitative metrics in Table 2. For instance, in Ireland, the augmentation strategy reduces the Autoformer’s horizon 4 mean *IS*_80_ from 119.7 (real) down to 73.27 (aug), while in France, it substantially improves LSTM’s performance by decreasing the error from 643 (real) to 518.9 (aug). When directly comparing the two data augmentation, the results reveal that the use of endogenous (aug) data consistently outperforms the use of exogenous data (comb) across the majority of architectures and forecasting horizons. For instance, in France, the DLinear model trained under the augmented setting achieves a mean *IS*_80_ of 518.2 at horizon 4, showing a distinct advantage over the combined setting (610.4). Similarly, in Ireland, the augmented strategy yields a substantially lower error for LSTM (80.93) compared to its combined counterpart (121.2). Although the combined approach occasionally produces lower errors in isolated scenarios, such as for LSTM in Belgium, the overall trend is clear. This suggests that while augmenting the training space is highly beneficial for deep learning architectures, an overly heterogeneous data pool (as introduced by the exogenous data) may risk diluting localized epidemic signals or injecting excessive noise. Consequently, the more targeted augmented strategy provides a more balance of signal enrichment and regularization. Finally, our weighted ensemble effectively leverages these diverse architectural biases delivering highly robust probabilistic forecasts. As visually indicated in Figure 1, the weighted ensemble frequently exhibits tighter variance and lower median relative *IS*_80_ values compared to deep learning models. By aggregating the predictive distributions, it successfully mitigates the volatility of individual architectures. The quantitative metrics in Table 2 explicitly confirm this stabilization; for instance, in Czechia at horizon 4, the weighted ensemble achieves a mean *IS*_80_ of 124.9, successfully outperforming all individual deep learning configurations (e.g., DLinear aug at 160.6 and LSTM aug at 144.8), TabPFN-TS (237.4) and offering a highly competitive benchmark.

**Table 2:** Probabilistic forecasting performance (Mean *IS*_80_) across nine European countries. The table details the Mean Interval Score for each model across four forecasting horizons 1 to 4. Lower values denote better probabilistic performance, reflecting sharper and better-calibrated predictive distributions. Training data strategies for deep learning models are denoted identically to those in Fig. 1.

| Country | Step | Naive | ARIMA | SEIR | DLinear |  |  | LSTM |  |  | Autoformer |  |  | TabPFN<sub>ts</sub> | Respicast | Ensemble |
| --- | --- | --- | --- | --- | --- | --- | --- | --- | --- | --- | --- | --- | --- | --- | --- | --- |
|  |  |  |  |  | real | aug | combined | real | aug | combined | real | aug | combined |  |  |  |
| Belgium | 1 | 653 | 663.9 | 668.9 | 1031 | 628.4 | 783.3 | 1276 | 829.2 | **503.4** | 1150 | 745.7 | 993.4 | 514.2 | 778.6 | 507.7 |
|  | 2 | 1132 | 1171 | 833.7 | 2439 | 991.4 | 1154 | 1423 | 1329 | 675.7 | 1447 | 1036 | 1205 | 676.1 | **639.6** | 653.3 |
|  | 3 | 1618 | 1483 | 1032 | 1459 | 1086 | 1345 | 1585 | 1734 | 854.6 | 1721 | 1051 | 1003 | 835.9 | **624.8** | 798.6 |
|  | 4 | 2164 | 1866 | 1118 | 2057 | 1475 | 1661 | 1716 | 1337 | 1049 | 2185 | 1138 | 1413 | 953.3 | **697.3** | 1004 |
| Czechia | 1 | 93.19 | 80.54 | 161.7 | 135.1 | 69 | 98.92 | 130.9 | 82.22 | 90.74 | 167.9 | 110.9 | 144.7 | **47.9** | 101.4 | 52.5 |
|  | 2 | 148.9 | 116.2 | 259 | 191.9 | 106.3 | 132.9 | 160.4 | 80.59 | 163.8 | 203.1 | 95.61 | 150.4 | 95.04 | 97.64 | **69.12** |
|  | 3 | 208.5 | 149.7 | 337.8 | 169.7 | 133.9 | 169.6 | 173.7 | 103 | 224.3 | 200 | 122.7 | 182.5 | 148.7 | 98.88 | **90.93** |
|  | 4 | 241.8 | 169.8 | 392.2 | 212.8 | 160.6 | 187.1 | 178.4 | 144.8 | 294.9 | 215.5 | 167.2 | 196.7 | 237.4 | **112** | 124.9 |
| Denmark | 1 | 105.3 | 111.8 | 426.8 | 114 | 103.6 | 202.7 | 125.8 | 117.2 | 138.5 | 141 | 154.5 | 152 | 108 | 98.85 | **96.52** |
|  | 2 | 111.3 | 115.8 | 533.2 | 249.9 | 109.4 | 244.2 | 152 | 148 | 206.4 | 171.3 | 173 | 155.3 | 119.8 | 99.75 | **99.09** |
|  | 3 | 148.8 | 145.6 | 643.9 | 204 | 119 | 253.6 | 186.9 | 121.5 | 200.9 | 198 | 235.2 | 263.1 | 139 | **106.8** | 117.1 |
|  | 4 | 169.1 | 165 | 779.7 | 190.7 | 131.3 | 262 | 200.6 | 189.7 | 215.6 | 240.9 | 210.4 | 348.5 | 162.3 | **119.9** | 140.7 |
| France | 1 | 253.1 | 215.6 | 291.1 | 360.7 | 209.5 | 287.9 | 412 | 243.4 | 224.5 | 424.5 | 313 | 336 | **163** | 277.8 | 163.1 |
|  | 2 | 433.6 | 381.5 | 352.2 | 774.7 | 391.8 | 442.4 | 502 | 412.9 | 426 | 480.7 | 312.9 | 405.2 | 216 | **178.8** | 217 |
|  | 3 | 600.5 | 502.9 | 411.6 | 511.5 | 430.5 | 497 | 569 | 507.6 | 514.3 | 683.2 | 466.2 | 751.4 | 283.7 | **192.6** | 288.9 |
|  | 4 | 783.3 | 600.5 | 476.2 | 708.1 | 518.2 | 610.4 | 643 | 518.9 | 607.9 | 668.8 | 494.6 | 1094 | 380.9 | **228.7** | 397.3 |
| Ireland | 1 | 56.5 | 49.28 | 103.5 | 67.55 | 54.59 | 89.29 | 67 | 48.16 | 56.97 | 78.21 | 57 | 41.65 | 41.43 | 55.55 | **40.06** |
|  | 2 | 101.9 | 67.69 | 129.9 | 79.52 | 99.57 | 129.9 | 72.46 | 38.47 | 84.92 | 74.18 | 78.56 | 86 | 40.79 | 55.48 | **38** |
|  | 3 | 126 | 74.96 | 164.8 | 71.49 | 108.6 | 167.6 | 74.8 | 44.54 | 85 | 89.07 | 58.91 | 137.5 | **36.78** | 46.07 | 38.22 |
|  | 4 | 155.7 | 81.52 | 201.3 | 103.7 | 129.9 | 186.7 | 78.24 | 80.93 | 121.2 | 119.7 | 73.27 | 151.1 | **41.2** | 55.49 | 49.48 |
| Italy | 1 | 435.8 | 301.3 | 1751 | 818.7 | 281.3 | 470.9 | 991.1 | 846.1 | 357.4 | 1161 | 500.9 | 471 | **273.9** | 638.3 | 310.7 |
|  | 2 | 826.7 | 629 | 2553 | 2552 | 541.9 | 810.7 | 1239 | 1102 | 780.7 | 1001 | 802 | 670.2 | **459.9** | 746.6 | 497.3 |
|  | 3 | 1166 | 1005 | 3494 | 1786 | 813.9 | 968.4 | 1613 | 1450 | 1138 | 1295 | 1077 | 861.4 | **692.7** | 947.1 | 710.1 |
|  | 4 | 1391 | 1348 | 4467 | 1526 | 978.5 | 1204 | 1568 | 1649 | 1571 | 1700 | 1212 | 1126 | **869.6** | 1405 | 993.2 |
| Netherlands | 1 | 48.02 | 49.18 | 80.61 | 72.33 | 39.39 | 50.63 | 86.12 | 66.42 | **30.86** | 87.75 | 74.34 | 70.48 | 39.54 | 43.21 | 36.9 |
|  | 2 | 78.81 | 74.75 | 112.1 | 225.6 | 65.16 | 89.74 | 95.37 | 98.96 | 74.3 | 105.3 | 109.6 | 108.5 | 45.75 | **30.24** | 56.79 |
|  | 3 | 122.5 | 95.12 | 143.7 | 118 | 87.4 | 110 | 99.35 | 124.5 | 111.9 | 122.7 | 117.6 | 163.1 | 56.64 | **37.94** | 67.69 |
|  | 4 | 171.8 | 126.8 | 168 | 116.9 | 118 | 155.4 | 102.7 | 150.8 | 156.2 | 105.3 | 185.5 | 127.8 | 69.81 | **48.57** | 81.75 |
| Poland | 1 | 355.2 | 312.7 | 897.2 | 327.6 | 358.2 | 600.9 | 397.3 | 556.3 | 330.8 | 394.6 | 345 | 479.9 | **281.1** | 300.5 | 296.9 |
|  | 2 | 433.2 | 404 | 1166 | 499.9 | 516.5 | 951.4 | 461.7 | 640.3 | 670.7 | 383.7 | 453.2 | 724.6 | 386.5 | **243.9** | 355.1 |
|  | 3 | 485.7 | 465.6 | 1414 | 414.3 | 525.6 | 1180 | 396.4 | 647.6 | 815.4 | 469.2 | 513.8 | 1113 | 524.3 | **311.2** | 410.9 |
|  | 4 | 621.1 | 555.5 | 1682 | 559.9 | 688.1 | 1300 | 406.7 | 826.3 | 1028 | 500.2 | 671.5 | 1717 | 714.5 | **346.5** | 474.1 |
| Romania | 1 | 27.36 | 25.72 | 51.81 | 52.09 | 22.22 | 23.22 | 75.54 | 53.92 | 33.14 | 63.56 | 24.97 | 27.53 | 25.13 | 31.45 | **21.03** |
|  | 2 | 42.95 | 49.52 | 67.2 | 123.6 | 44.18 | 39.79 | 81.45 | 59.37 | 45.91 | 72.32 | 45.46 | 37.82 | 32.64 | 31.99 | **28.91** |
|  | 3 | 57.79 | 77.11 | 89.55 | 90.46 | 53.94 | 61.22 | 99.22 | 51.61 | 54.24 | 94.84 | 60.45 | 60.13 | 57.33 | **34.82** | 45.07 |
|  | 4 | 69 | 110.3 | 116.2 | 105.1 | 61.08 | 78.66 | 106.2 | **50.21** | 82.89 | 122 | 68.13 | 70.65 | 109.9 | 54.15 | 62.99 |

**Figure 1:**
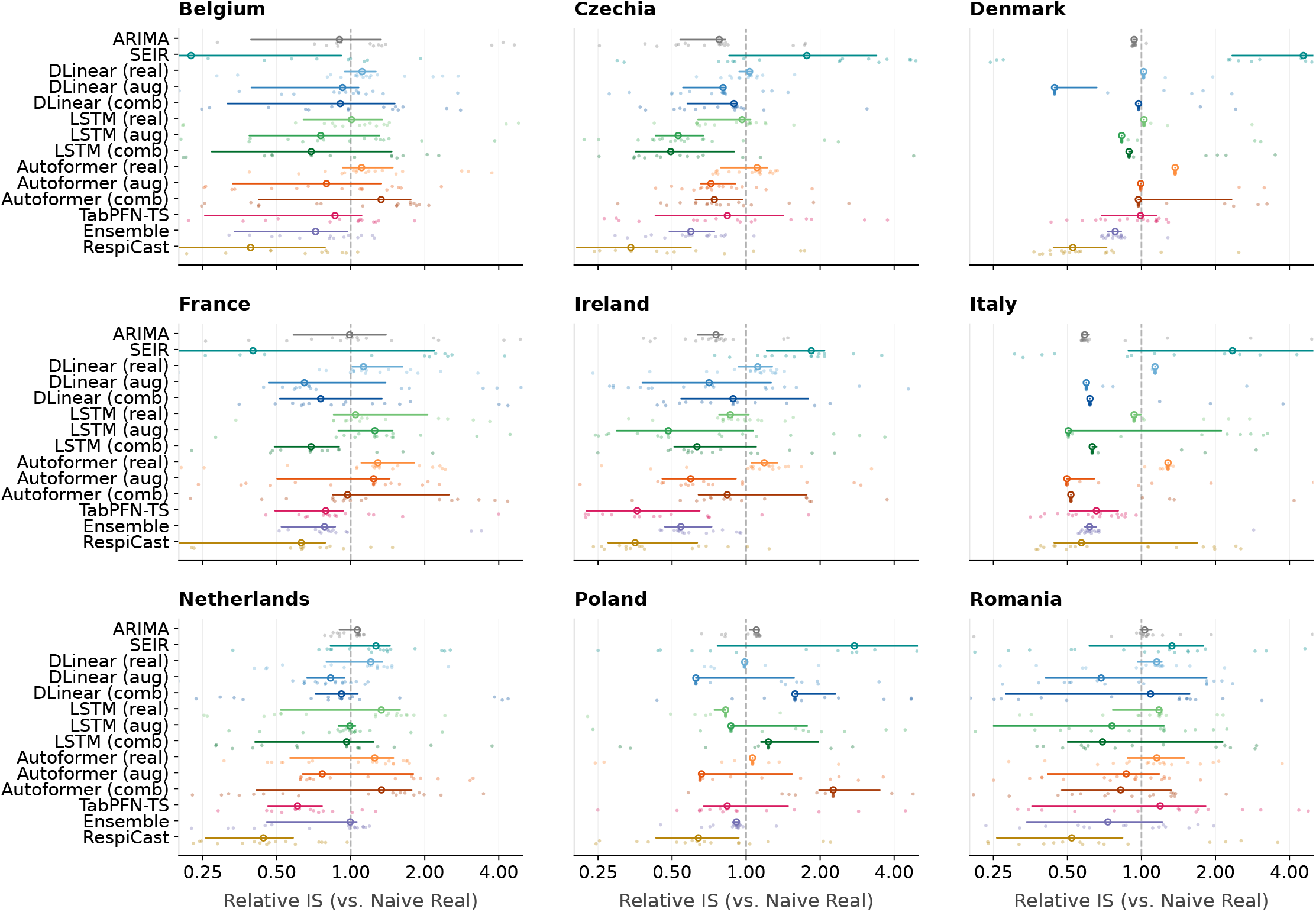
Probabilistic forecasting performance at horizon 4 (i.e., 4-week-ahead). Distribution of the Relative Interval Score (Relative IS) across nine European countries. The vertical dashed line at 1.0 represents the naive baseline; values *<* 1.0 indicate improved probabilistic accuracy. For deep learning models, suffixes denote training data strategies: **(real)** for real data only, **(aug)** for endogenous data, and **(comb)** for exogenous data. The x-axis is on a logarithmic scale.

Next, we evaluate the point forecast accuracy across all models using the Mean Absolute Error (MAE) and the Relative Absolute Error (RAE) with respect to the naive baseline (see Table 3 and Fig. 2, respectively). Consistently with our probabilistic evaluations, the RespiCast ensemble establishes a highly competitive benchmark, achieving a lowest MAE in 6 out of 9 countries at horizon 4. As shown in the RAE distributions, RespiCast frequently demonstrates tight error variances and median values consistently below 1.0, underlining its robustness across diverse epidemiological contexts. Again, the foundation model, TabPFN-TS, exhibits good zero-shot point forecasting capabilities. The RAE distributions reveal that TabPFN-TS achieves median errors well below the naive baseline in 6 countries. MAE metrics support this advantage: in Denmark, TabPFN-TS achieves the lowest MAE across all four horizons. Similarly, in Ireland, TabPFN-TS (MAE 9.054) maintains an advantage over RespiCast (10.1) and the naive baseline (24.33) at horizon 4. The SEIR model demonstrated a highly polarized performance. In specific regions like Belgium, France and Romania, it achieved some of lowest horizon 4 MAE scores. However, its accuracy degrades significantly in contexts like Italy and Poland. Similarly, ARIMA exhibits mixed point forecasting accuracy. While it successfully reduces the horizon 4 MAE compared to the naive baseline in regions like Belgium (304.3 vs 408.4), France (126 vs 164.9) and Italy (269.1 vs 397.4), it lacks robustness. In countries such as Denmark and Poland, ARIMA’s prediction errors actually exceeds the naive baseline (40.42 vs 38.87, and 135.6 vs 131.4, respectively), underscoring the limitations of traditional linear autoregressive methods in capturing complex epidemic trajectories. The deep learning architectures trained exclusively on the limited real data (real) exhibit high variance and frequent underperformance. At the horizon 4, the RAE distributions for these unaugmented models routinely span far above 1.0, indicating failures to beat even the naive baseline. The quantitative data strongly supports this visual trend. In Belgium, DLinear real reaches an MAE of 437.5, exceeding the naive model’s 408.4 at horizon 4. Likewise, in France, DLinear real yields an MAE of 193.4, which is notably worse than the naive baseline of 164.9. The integration of our data augmentation pipeline significantly mitigates this instability. The RAE plots illustrate a visible leftward shift, representing reduced relative error and tightened variance for augmented models compared to their real counterparts. When comparing the two enrichment strategies, the use of endogenous (aug) data is confirmed to yield more reliable improvements over the exogenous (comb) pool. For instance, in Belgium, the augmented strategy substantially reduces the MAE across all three deep learning architectures compared to the real training data. Building upon the optimized individual models, the weighted ensemble achieves competitive point forecasting performance, particularly at short-to-medium horizons, with the lowest MAE in several settings such as Czechia horizon 2 (15.15), France horizons 1 − 2 and (29.65, 40.77), Ireland horizons 1 and 3 (7.077, 7.641), and Italy horizons 1 and 2 (56.12, 99.85). However, its advantage is not evident at longer horizons where RespiCast and TabPFN-TS often perform better.

**Table 3:** Point forecast accuracy (MAE) across nine European countries. The table presents the MAE for all evaluated models across four forecasting horizons 1 to 4. Lower values indicate superior point forecasting accuracy. Training data strategies for deep learning models are denoted identically to those in Fig. 1.

| Country | Step | Naive | ARIMA | SEIR | DLinear |  |  | LSTM |  |  | Autoformer |  |  | TabPFN-ts | Respicast | Ensemble |
| --- | --- | --- | --- | --- | --- | --- | --- | --- | --- | --- | --- | --- | --- | --- | --- | --- |
|  |  |  |  |  | real | aug | combined | real | aug | combined | real | aug | combined |  |  |  |
| Belgium | 1 | 113.7 | 107.2 | 93.34 | 204.2 | 105.1 | 143.8 | 226.7 | 120.6 | **88.86** | 231.1 | 158 | 172.2 | 100.2 | 139.7 | 95.79 |
|  | 2 | 225.2 | 194.2 | 119.1 | 408 | 176.1 | 215.7 | 274.6 | 169.2 | 142.8 | 297.6 | 172 | 245 | **107.3** | 137 | 127.3 |
|  | 3 | 311.7 | 243.5 | 144.3 | 294.5 | 207.4 | 225.6 | 294.2 | 202.1 | 197.5 | 383.1 | 234 | 200.7 | 135.9 | **130.3** | 143.7 |
|  | 4 | 408.4 | 304.3 | 158.4 | 437.5 | 240.3 | 254.7 | 311.9 | 229.1 | 232.3 | 471.1 | 289.3 | 310.6 | 188.6 | **138.1** | 180.9 |
| Czechia | 1 | 18.3 | 15.14 | 21.14 | 33.17 | 14.33 | 16.13 | 36.49 | 12.58 | 15.6 | 38.57 | 23.36 | 26.19 | **9.101** | 16.47 | 11.6 |
|  | 2 | 33.31 | 25.73 | 33.44 | 61.15 | 25.39 | 28.04 | 39.92 | 19.02 | 28.67 | 47.03 | 21.27 | 31.02 | 19.99 | 19.19 | **15.15** |
|  | 3 | 48.17 | 39.4 | 45.24 | 41.67 | 38.91 | 40.59 | 48.54 | 31.69 | 43.43 | 63.16 | 28.66 | 40.9 | 36.78 | **21.29** | 24.81 |
|  | 4 | 64.17 | 46.47 | 57.31 | 61.76 | 47.74 | 52.63 | 49.29 | 45.7 | 57.49 | 68.52 | 40.46 | 46.48 | 52.12 | **23.47** | 38.38 |
| Denmark | 1 | 22 | 22.01 | 63.21 | 52.78 | 24.08 | 37.61 | 24.9 | 34.54 | 26.96 | 29.59 | 26.31 | 33.26 | **20.32** | 23.63 | 23.58 |
|  | 2 | 29.35 | 29.69 | 73.73 | 149.9 | 31.82 | 45.39 | 48.16 | 32.57 | 42.38 | 32.57 | 38.58 | 38.37 | **20.43** | 21.7 | 26.78 |
|  | 3 | 35.89 | 36.21 | 85.87 | 79.73 | 37.25 | 51.6 | 41.89 | 39.45 | 49.24 | 44.14 | 53.72 | 51.64 | **21.14** | 23.62 | 27.29 |
|  | 4 | 38.87 | 40.42 | 97.4 | 41.93 | 44.8 | 59.06 | 46.13 | 44.11 | 49.23 | 43.11 | 45.99 | 70.6 | **20.92** | 27.2 | 29.48 |
| France | 1 | 44.89 | 35.66 | 38.13 | 102.4 | 33.96 | 42.91 | 91.98 | 51.43 | 39.08 | 100.7 | 60.22 | 49.99 | 30.39 | 49.42 | **29.65** |
|  | 2 | 89.73 | 67.39 | 45.56 | 201.1 | 62.82 | 74.03 | 120.9 | 81.47 | 66.55 | 110 | 71.69 | 60.56 | 50.67 | 48.88 | **40.77** |
|  | 3 | 127.7 | 101.4 | 55.12 | 124.2 | 88.6 | 105.3 | 131.4 | 110.7 | 92.33 | 169.2 | 111.2 | 117.2 | 74.39 | **53.5** | 56.1 |
|  | 4 | 164.9 | 126 | 64.95 | 193.4 | 109.8 | 115.6 | 142.2 | 128.8 | 111.6 | 201.9 | 153.7 | 170.9 | 91.53 | **60.56** | 66.2 |
| Ireland | 1 | 8.443 | 7.321 | 11.81 | 13.61 | 8.795 | 12.99 | 12.36 | 9.176 | 9.144 | 16.78 | 12.35 | 8.13 | 7.13 | 9.185 | **7.077** |
|  | 2 | 14.31 | 10.99 | 14.82 | 21.39 | 12.77 | 16.13 | 12.82 | 8.824 | 14.69 | 17.18 | 12.15 | 12.78 | **7.888** | 9.381 | 8.229 |
|  | 3 | 18.58 | 12.82 | 19.04 | 14.96 | 15.55 | 20.8 | 14.74 | 8.345 | 17.59 | 22.13 | 11.47 | 19.33 | 8.031 | 9.419 | **7.641** |
|  | 4 | 24.33 | 15.46 | 23.09 | 24.53 | 18.18 | 22.92 | 16.39 | 11.72 | 23.72 | 28.48 | 15.95 | 24.43 | **9.054** | 10.1 | 10.26 |
| Italy | 1 | 106.4 | 57.73 | 313.1 | 339.2 | 69.45 | 86.91 | 246.5 | 200.8 | 69.82 | 261.4 | 89.6 | 105 | 57.79 | 124.5 | **56.12** |
|  | 2 | 207 | 121.5 | 406.2 | 956.6 | 157.1 | 173.8 | 387.8 | 283.6 | 152.1 | 272.7 | 139.1 | 136.7 | 100.8 | 148.8 | **99.85** |
|  | 3 | 305.8 | 188.5 | 505.4 | 535 | 297.8 | 282.8 | 412.1 | 395.3 | 244.5 | 351.3 | 196.5 | 179.4 | **153.1** | 201.7 | 159.6 |
|  | 4 | 397.4 | 269.1 | 605.8 | 459.2 | 451.2 | 470.3 | 440.6 | 505 | 382.9 | 466.4 | 233.9 | 226 | **219.4** | 265.2 | 244.9 |
| Netherlands | 1 | 10.61 | 10.57 | 11.91 | 16.78 | 11.36 | 12.52 | 23.41 | 14.26 | 9.286 | 19.14 | 13.24 | 16.16 | **8.612** | 11.69 | 8.938 |
|  | 2 | 16.69 | 16.68 | 15.96 | 32.79 | 17.96 | 16.32 | 23.6 | 21.47 | 13.29 | 25.47 | 20.69 | 22.54 | **8.493** | 10.25 | 12.51 |
|  | 3 | 24.55 | 24.6 | 21.07 | 23.27 | 22.81 | 21.29 | 28.09 | 25.06 | 24.82 | 30.26 | 24.53 | 31.11 | **10.2** | 10.63 | 17.97 |
|  | 4 | 31.34 | 31.44 | 25.31 | 32.44 | 28.86 | 26.67 | 27.96 | 31.29 | 35.78 | 29.58 | 29.3 | 35.92 | 14.61 | **10.44** | 22.53 |
| Poland | 1 | 63.72 | 67.94 | 145 | 97.82 | 62.34 | 111.2 | 85.98 | 91.33 | 64.89 | 93.81 | 72.27 | 95.48 | 58.56 | **56.05** | 59.15 |
|  | 2 | 108.3 | 108.7 | 177.8 | 210.6 | 93.58 | 171.8 | 121.8 | 121.6 | 111.7 | 101.4 | 97.48 | 149.3 | 78.46 | **57.24** | 100.1 |
|  | 3 | 121.6 | 124.5 | 201.3 | 131.8 | 97.2 | 194.7 | 112.9 | 113.9 | 164.8 | 117.1 | 119.6 | 235.4 | 111.9 | **59.25** | 106 |
|  | 4 | 131.4 | 135.6 | 235.7 | 129.6 | 102.7 | 213 | 124.2 | 110.2 | 223.9 | 147.5 | 132.3 | 359.9 | 148.2 | **63.02** | 119.1 |
| Romania | 1 | 4.9 | 4.682 | 6.186 | 9.633 | 4.842 | 4.859 | 12.53 | 8.751 | 6.231 | 11.56 | 6.34 | **4.498** | 5.567 | 6.007 | 4.786 |
|  | 2 | 9.825 | 8.932 | 7.805 | 18.77 | 8.828 | 7.646 | 14.91 | 11.27 | 8.128 | 13.09 | 8.169 | 7.762 | 9.826 | 6.303 | **6.087** |
|  | 3 | 14.16 | 13.27 | 10.04 | 16.21 | 13.04 | 12.15 | 16.78 | 11.53 | 10.52 | 16.64 | 11.52 | 12.16 | 14.71 | **7.706** | 11.18 |
|  | 4 | 18.61 | 18.41 | 12.75 | 17.93 | 16.17 | 14.36 | 17.5 | 13.06 | 12.76 | 20.88 | 14.61 | 15.17 | 18.31 | **9.891** | 13.75 |

**Figure 2:**
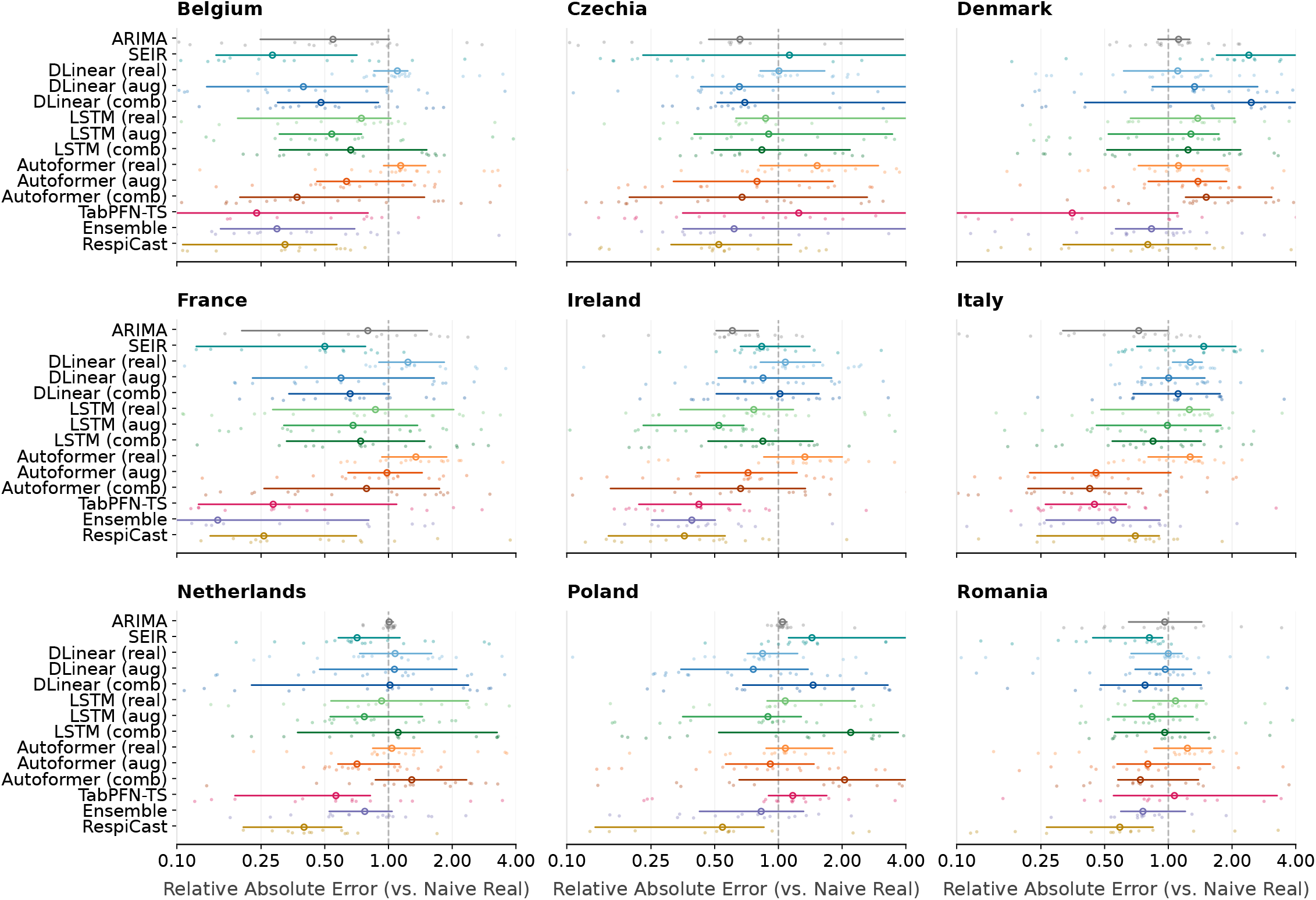
Point forecast accuracy at horizon 4 (i.e., 4-week-ahead). Distribution of the Relative Absolute Error (Relative AE) across nine European countries. The vertical dashed line at 1.0 marks the naive baseline; values *<* 1.0 indicate a reduction in absolute prediction error. Model suffix definitions and plotting conventions are identical to those in Fig. 1.

### Overall models’ performance across forecasting horizons

To show the performance of the models succinctly, in Fig. 3 we plot the cumulative *wins count* across all evaluation contexts (i.e., countries and weeks) for each forecasting horizon. A win is defined when a given model achieves the strictly lowest error in a specific evaluation context (across all countries and evaluation weeks). When considering point forecasts accuracy (see Fig. 3a) we observe a clear shift in models performance as the forecasting horizon extends. At shorter horizons (1 and 2), TabPFN-TS and our weighted ensemble are most competitive. Both achieve 3 out of 9 wins at horizon 1 and 4 out of 9 wins at horizon 2. However, at the more challenging extended horizons (i.e., 3 and 4), the RespiCast ensemble demonstrates superior performance, decisively overtaking other models securing the highest number of wins (5 out of 9 at horizon 3, and 6 out of 9 at horizon 4). In terms of probabilistic forecasting (see Fig. 3b), TabPFN-TS performs best at horizon 1 with 4 wins, while the weigthed ensemble and RespiCast jointly lead at horizon 2 with 4 wins each. Nevertheless, as the forecasting window increases, RespiCast shows its dominance, capturing the majority of wins (6 for both 3 and 4 horizons). Individual deep learning architectures (such as LSTM and Autoformer) rarely secured the strictly optimal score, recording only isolated single wins across all evaluation contexts.

**Figure 3:**
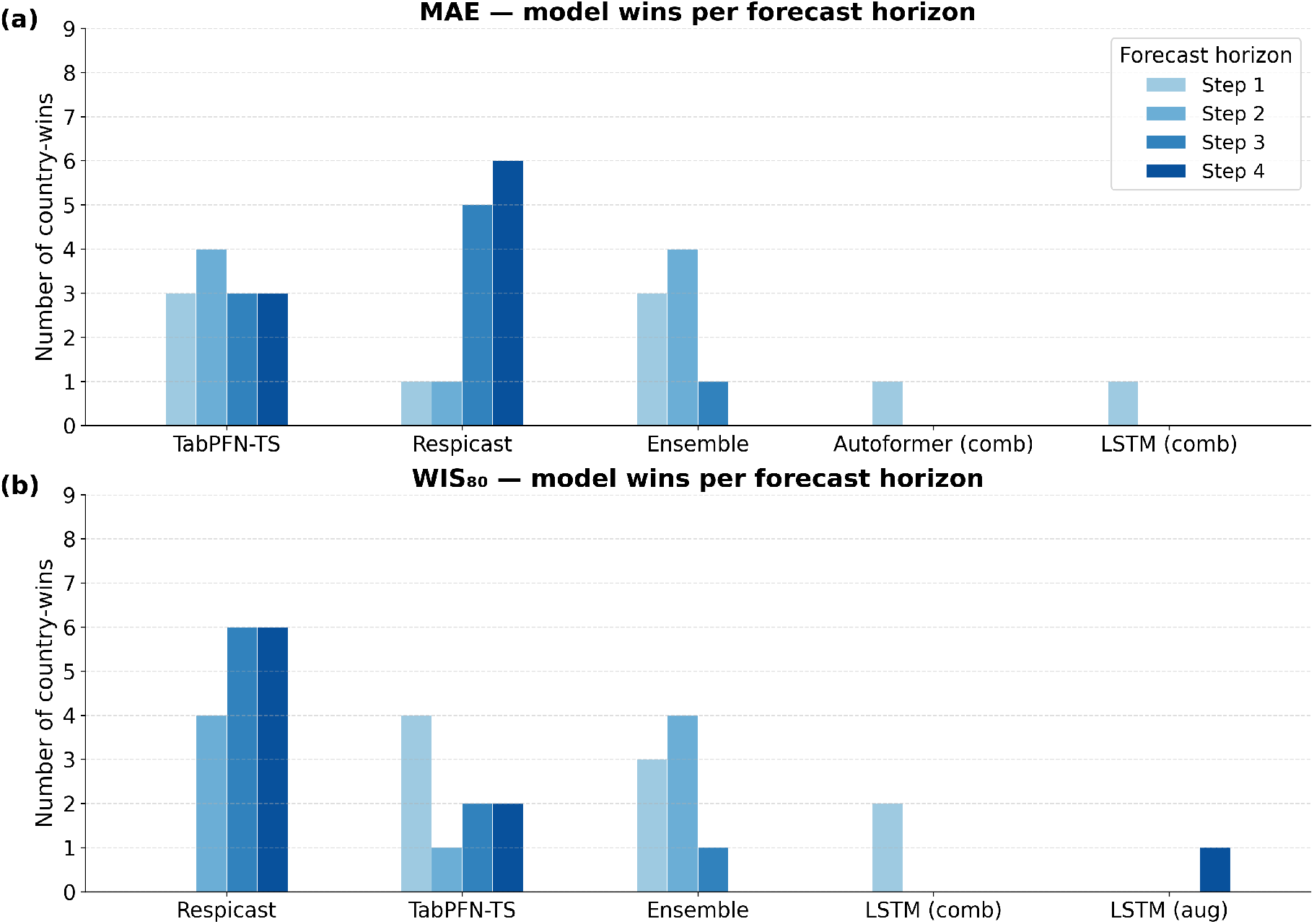
Cumulative wins counts across forecasting horizons and countries. Stacked bars illustrate the frequency with which each methodological paradigm emerges as the top performer. Panel (a) presents results for point forecast accuracy (by MAE), and Panel (b) presents results for probabilistic forecast accuracy (by *IS*_80_). The colors refer to different forecasting horizons.

### Epidemic trajectories

To complement our quantitative error metrics, we visually inspect the dynamics of the epidemic trajectories during the 2024 − 2025 Influenza season. Fig. 4 illustrates the 4-week-ahead forecasts (i.e., horizon 4) across all nine European countries. For each country we compare the RespiCast ensemble against the best-performing alternative model in each country, dynamically selected based on the lowest MAE. Observing the trajectories highlights the distinct challenges of epidemic forecasting, particularly during the rapid growth and peak phases of the season. We note how the best alternative model varies by region. Interestingly, the stand-alone SEIR model emerges as the most accurate point-forecaster in Belgium, France and Romania. Furthermore, the value of our data augmentation strategy is evident in Poland, where the endogenously augmented DLinear architecture (DLinear aug) successfully emerges as the top-performing alternative. In other regions, the foundation model TabPFN-TS and our ensemble demonstrate a remarkable capacity to reproduce the epidemic curves and maintain well-calibrated uncertainty bounds. While RespiCast remains a robust operational benchmark, these comparative trajectories underscore how tailored mechanistic priors and zero-shot foundation models can effectively capture distinct epidemiological shifts.

**Figure 4:**
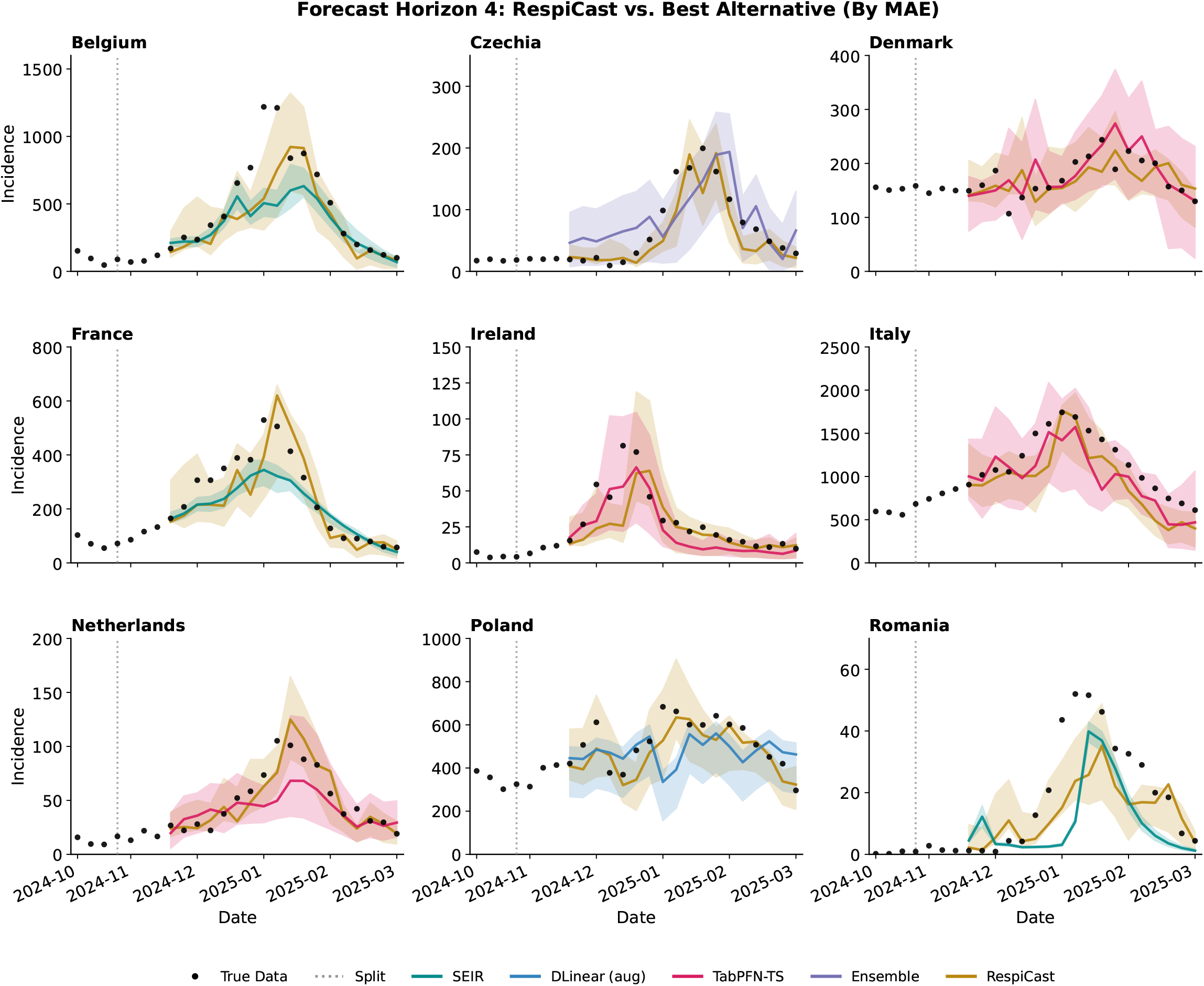
Epidemic trajectory forecasts at horizon 4 (4-week-ahead) across nine European countries during the 2024 − 2025 influenza season. The plots visually compare the predictions of the RespiCast ensemble against the best-performing alternative model for each respective country, selected based on the lowest Mean Absolute Error (MAE). Black dots represent the true observed weekly incidence of Influenza-Like Illness (ILI). The vertical dashed line marks the forecast origin (split). Solid lines denote the point forecasts, while the corresponding shaded regions represent the 80% prediction intervals, illustrating the models’ capacity to capture complex epidemic peaks and quantify uncertainty.

## Discussion

In this study, we systematically evaluated classical statistical methods, traditional epidemic compartmental models, deep learning architectures, and emerging foundation models for 1 to 4 week-ahead forecasts of Influenza-Like Illness (ILI) across nine European countries. By evaluating these methodologies alongside a naive baseline and the RespiCast ensemble, our framework bridges the gap between methodological innovation and the practical constraints of operational epidemiology.

Our analysis reveals three primary insights into the current landscape of epidemic forecasting. First, we demonstrate that data-intensive deep learning architectures—including DLinear, LSTM, and Autoformer—are constrained by the severe data scarcity inherent in epidemic forecasting. When trained exclusively on surveillance data, these models exhibit high variance and systematic instability, often failing to outperform even the naive baseline at 4-week-ahead horizons. Second, we find that this performance gap can be effectively bridged through targeted data augmentation strategies, though the source of synthetic data is critical. While exogenous augmentation using mechanistic epidemic simulations is theoretically attractive for embedding biological priors, our results reveal that it can inadvertently introduce biases. In contrast, our endogenous augmentation strategy, which enriches the training space via temporal translations and realistic observational noise, yields more robust and consistent improvements in predictive accuracy. Third, our study highlights the transformative potential of foundation models, which offer a paradigm shift in addressing data scarcity. The time-series foundation model, TabPFN-TS, demonstrates good zero-shot capabilities. By leveraging its task-agnostic pre-training, it performs robust Bayesian inference without the need for task-specific fine-tuning. TabPFN-TS consistently outperforms augmented deep learning architectures and rivals or surpasses the RespiCast ensemble in regions such as Ireland and Italy, both in point (MAE) and probabilistic (*IS*_80_) forecasting metrics. In this context, recent work by Kalahasti et al. [22] has highlighted the broader potential of time-series foundation models for policy evaluation and infectious disease forecasting, especially short-term. Their findings align with our observations regarding the robustness of pre-trained models in low-data settings, further underscoring the shift toward task-agnostic method in epidemiology.

Our study is subject to some limitations. First, to ensure a highly controlled evaluation environment, our rolling-window forecasting setup used clean and empirical historical data. While this retrospective data reflects real-world epidemic dynamics, it represents finalized surveillance records. This approach is standard for rigorous benchmarking, but it does not fully replicate the conditions of live operational forecasting. In real-time public health scenarios, recent epidemiological observations are frequently subject to reporting delays, right-truncation, and subsequent backfill revisions, which means past data points continually change over time. Our current evaluation does not explicitly stress-test how these models might degrade when fed this unstable data. We note how that the RespiCast ensemble was generated live during the respective seasons and inherently absorbed real-time data instabilities. Second, although we evaluated a diverse taxonomy of models, our selection within specific methodological families is representative rather than exhaustive. For instance, we evaluated Autoformer and TabPFN-TS as our singular representatives for transformer-based architectures and time-series foundation models, respectively. Moving beyond these specific architectures remains a priority. Finally, our geographical scope is explicitly constrained to nine European countries. While this provides a robust and diverse dataset it does not represent the global heterogeneity of influenza transmission. Epidemiological dynamics in other countries with vastly different surveillance capacities may present unique forecasting challenges. Consequently, the generalizability of our findings to a global context remains to be systematically verified.

Moving forward, extensions of this work could focus on integrating multivariate data streams, such as real-time mobility metrics, genomic surveillance, and climate data, to further constrain and guide data-hungry architectures. Furthermore, the zero-shot success of TabPFN-TS highlights a critical frontier for computational epidemiology: the development of an epidemic-specific foundation model. Current state-of-the-art foundation models are pre-trained on generic, heterogeneous time-series collections. Consequently, they do not inherently capture the laws of disease transmission. Future research should focus on designing foundation models pre-trained exclusively on massive epidemiological datasets [42]. By exposing foundation models to highly diverse, simulated epidemiological scenarios during pre-training, we could embed mechanistic priors directly into their latent space. Such an epidemic-specific foundation model would bridge the divide between data-driven and mechanistic paradigms, offering public health agencies a zero-shot forecasting tool that possesses both the adaptability of deep learning and the structural rigour of theoretical epidemiology. Furthermore, moving beyond pure forecasting, recent evidence suggests that conversational large language models (LLMs) can assist public health practitioners in co-designing transmission models and estimating critical parameters directly from prevalence data [43]. The use of interactive agents could help bridging the gap between complex mathematical modelling and accessible public health decision-making.

## Materials and Methods

### Data

Our forecasting target is the weekly incidence of Influenza-Like Illness (ILI) defined as incidence rate per 100, 000 population. We used epidemiological surveillance data from nine geographically diverse European countries: Belgium, Czechia, Denmark, France, Ireland, Italy, Netherlands, Poland, and Romania. The dataset was obtained from the European Respiratory Virus Surveillance Summary (ERVISS) platform, managed by the European Centre for Disease Prevention and Control (ECDC) and the World Health Organization (WHO) Regional Office for Europe. The data for the 2017 − 2018 and 2018 − 2019 seasons were obtained from the Flu Forecast Hub archive [44], while the data for the 2023 − 2024 and 2024 − 2025 seasons were retrieved from the RespiCast Syndromic Indicators repository [45] (both originally sourced from ERVISS). In accordance with standard epidemiological reporting, each season is defined as starting from ISO week 42 and concluding on ISO week 14 of the subsequent year. Surveillance data spanning the COVID-19 pandemic seasons were deliberately excluded from our analysis. The unprecedented non-pharmaceutical interventions (NPIs) implemented during this period caused substantial disruptions to typical seasonal influenza transmission patterns [46–48].

### Data Augmentation

Deep learning architectures require substantial training data to generalize effectively and avoid overfitting [49]. However, a standard influenza season yields fewer than 30 observational data points. To alleviate this severe data scarcity and investigate the impact of different data generation paradigms, we developed a data augmentation pipeline. We generated distinct streams of synthetic time series to enrich the training space.

The first is an endogenous strategy based on synthetic trajectories by directly augmenting the historical surveillance data. To mimic the natural variability in epidemic peak timing, we applied temporal translations by randomly shifting the empirical epidemic curves by a factor of *k* ∈ [− 4, 4] weeks. This specific temporal window was empirically derived to reflect the natural cross-regional variance in epidemic peak timing. To simulate the inherent heteroskedasticity and imperfections in public health reporting processes, we injected a combination of multiplicative and additive Gaussian noise into the shifted trajectories. Specifically, the multiplicative noise was drawn from 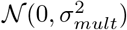, where *σ*_*mult*_ was uniformly sampled from [0.03, 0.12] and scaled by the local incidence magnitude. This noise scale was purposefully calibrated against the empirical surveillance data: across the evaluated countries, the historical incidence exhibited a median week-to-week relative variation consistently above 15% (ranging typically between 15% and 30%). Restricting the synthetic observational noise variance to a maximum of 12% ensures that it will not mask the underlying epidemiological signal. In addition to multiplicative variance, we incorporated an additive noise component scaled to 2% of the seasonal standard deviation. This accounts for the baseline fluctuations inherent in syndromic surveillance—such as the misclassification of other respiratory pathogens during off-peak weeks [50]. Finally, to maintain biological plausibility, a 3-to 5-week moving average filter was stochastically applied to the augmented trajectories with a 30% probability [51]. While smoothing is standard epidemiological practice to resolve single-week reporting anomalies (e.g., holiday backlogs), applying it stochastically acts as a structural regularizer. This ensures the training pool contains both volatile reporting scenarios and smooth transmission waves, preventing the models from over-fitting to specific noise patterns.

The second augmentation strategy is instead exogenous and obtained from a stochastic Susceptible-Exposed-Infectious-Recovered (SEIR) compartmental framework. The model was initially calibrated to the empirical incidence of each season via the ABC-SMC (Approximate Bayesian Computation with Sequential Monte Carlo) algorithm. For each country and each season, the algorithm selected 1000 trajectories with the lowest Weighted Mean Absolute Percentage Error (WMAPE) with respect to the data.

### Training configurations and data splitting

To systematically evaluate the impact of our data enrichment pipelines on the deep learning architectures, we defined three distinct training configurations. Models trained only on the historical surveillance data are denoted as *real*. Models trained on the real data supplemented with the synthetic trajectories generated via the statistical pipeline are designated as the augmented (aug) setting. Conversely, models using the real data combined with the simulated trajectories from the mechanistic SEIR pipeline are referred to as the combined (comb) setting.

Across all augmented configurations, to rigorously prevent data leakage, the datasets were partitioned such that the 2017–2018 and 2018–2019 seasons (plus their respective synthetic counterparts) served as the training set. The observed raw data from the 2023–2024 season was strictly used as a validation set for hyper-parameter tuning and early stopping, while the 2024–2025 season functioned solely as the unseen test set for the final out-of-sample evaluation.

### Forecasting setup

To mimic a real-time operational forecasting environment, we implemented a rolling-window strategy during the evaluation phase. The start of forecasts was initially set at ISO week 45 of each target season. At each weekly step *w*, all models were required to generate a 1 to 4 week-ahead forecast. Following each forecast generation, the observation window was sequentially advanced by one week to incorporate the most recent data point, and this process was repeated until the conclusion of the influenza season at ISO week 14. While the forecasting horizon of 4 weeks was strictly standardized across all evaluations to ensure equitable comparison, the length of the historical look-back window (*L*) used for inference was tailored to the specific architectural requirements of each method type:

- **Fixed-horizon sliding windows:** The deep learning architectures (LSTM, DLinear, and Autoformer) used a strictly fixed 4-week sliding window (*L* = 4) as their input sequence. This design ensures that these models focus on capturing immediate temporal dependencies and local epidemic gradients.
- **Expanding cumulative windows:** In contrast, methodologies relying on dynamic parameter calibration or extensive in-context learning (ARIMA, SEIR model, and the TabPFN-TS foundation model) employed an expanding cumulative window. This look-back strategy incorporated the entirety of the multi-season historical surveillance record in addition to the observed incidence of the current target season up to the first forecasting week. By leveraging the full cumulative history, these models were provided with the maximal epidemiological context necessary for stable parameter fitting or the construction of robust Bayesian support sets.

### Models

To comprehensively evaluate epidemic forecasting capabilities across different methodological paradigms, we employed a taxonomy of models. Let **y**_1:*t*_ = [*y*_1_, *y*_2_, …, *y*_*t*_] represent the historical incidence sequence, and our goal is to predict the future sequence **ŷ**_*t*+1:*t*+*H*_, where *H* is the forecasting horizon. In what follows, we provide a short summary of each model. We refer to the Supplementary Information for more details.

#### Baseline

As a minimal baseline, the naive model assumes the future incidence remains constant and equal to the last observed value: *ŷ*_*t*+*h*_ = *y*_*t*_ for all *h* ∈ [1, *H*]. In epidemiological forecasting, this serves as a critical diagnostic to ensure that complex models are genuinely anticipating transmission dynamics rather than merely echoing recent observations. Consequently, the naive model acts as the reference denominator for our relative performance metrics.

#### Benchmark

As performance benchmark, we considered RespiCast: a multi-model forecasting initiative funded by the ECDC [38]. It systematically aggregates outputs from diverse models developed by many teams. For our evaluation, we directly extracted the official 1-to 4-week-ahead predictive quantiles (10^*th*^ and 90^*th*^ percentiles) generated by the RespiCast hub ensemble for the respective target seasons. To ensure a complete, continuous evaluation and strict comparative parity with our models, particularly because publicly aggregated metrics can occasionally contain missing evaluations for specific target weeks, we independently computed the 80% Interval Score (*IS*_80_) for the RespiCast predictions using our rolling-window target dates and scoring functions.

#### Mechanistic epidemic model

As example of traditional epidemic compartmental model we considered an age-stratified Susceptible-Exposed-Infected-Recovered (SEIR) model [10, 35]. The model features vaccinations and is calibrated to the historical data using ABC-SMC algorithm [41]. The forecasts are generated using posterior distributions inferred from previous seasons. Within each forecasting window, model trajectories are evaluated against observed data, and those with the lowest wMAPE are selected.

#### Statistical model

As example of traditional statistical framework we considered the AutoRegressive Integrated Moving Average (ARIMA) [34]. It models the differenced stationary time series 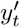 (where *d* is the order of differencing) as a linear combination of its past values and past forecast errors:

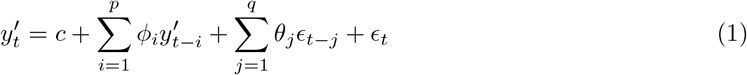

where *p* is the autoregressive order, *q* is the moving average order, *ϕ*_*i*_ and *θ*_*j*_ are the respective parameters, and *ϵ*_*t*_ is white noise. We used a non-seasonal auto-ARIMA procedure to select (*p, d, q*) independently at each rolling forecast origin. At each origin, the model was re-fitted using all observations available up to that time and then used to generate forecasts for horizons 1–4 weeks ahead. The candidate orders followed the standard auto-ARIMA search range used in common implementations: *p* ∈ [0, 5], *d* ∈ [0, 2], and *q* ∈ [0, 5], with the additional constraint *p* + *q* ≤ 5. The degree of differencing was selected using the KPSS stationarity test, and the final model order was chosen by minimizing the Akaike Information Criterion (AIC), following the automatic ARIMA selection approach of Hyndman and Khandakar [52].

#### Deep learning architectures

We considered three deep learning architectures: Long Short-Term Memory (LSTM) network, DLinear, and Autoformer.

The LSTM network is a specialized Recurrent Neural Network (RNN) designed to overcome gradients’ problem [36]. It uses a gating mechanism (forget, input, and output gates) to regulate the flow of information, allowing the network to capture complex, long-term temporal dependencies. To adapt this architecture for our multi-horizon evaluation, we implemented a 2-layer LSTM with dropout regularization. Rather than using a standard autoregressive rollout, we employed a direct multi-step forecasting strategy. Let 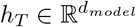 denote the final hidden state of the encoded historical sequence. To simultaneously predict the entire *H*-week forecasting horizon, *h*_*T*_ is processed through a layer normalization block and a linear projection head:

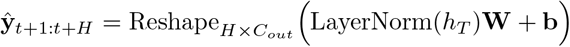

where 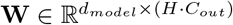 and 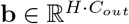 are the trainable weight and bias of the projection layer, and the output is reshaped to yield the multivariate predictions over the horizon *H*. By this projection, we explicitly mitigated compounding errors, establishing a stronger and more reliable pure-sequence benchmark.

DLinear is a highly efficient linear architecture optimized for time-series forecasting [37]. It decomposes the input sequence **X** into a moving-average trend component (**X**_*trend*_) and a seasonal remainder component (**X**_*seasonal*_). Two single-layer linear networks are then applied separately to these components to forecast the future:

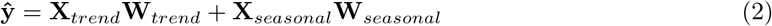

where **W**_*trend*_ and **W**_*seasonal*_ are the respective weight matrices. By stripping away non-linear activations and self-attention, DLinear enables us to empirically test whether seasonal influenza forecasting requires deep non-linear architectures, or if simple linear decomposition suffices.

Autoformer is a state-of-the-art deep learning architecture specifically designed for long-term time-series forecasting, offering significant structural and algorithmic advantages over standard attention-based models [17]. Unlike LSTM and DLinear, it uses both historical incidence sequences and exogenous temporal embeddings (time marks) to fully inform its attention layers. Typical transformers treat time series as uniform sequences of tokens. Instead, Autoformer assumes that empirical data inherently consists of hidden periodic patterns mixed with macroscopic trends. To capture this, it introduces a deep decomposition architecture that progressively isolates complex cyclical fluctuations from underlying trend dynamics throughout the network layers. Formally, for a hidden state representation ℋ^*l*^ at the *l*-th layer, the internal series decomposition block extracts the trend-cyclical part 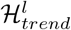 and the seasonal part 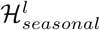 via an average pooling operation over a sliding window:

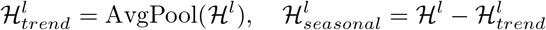

Beyond structural decomposition, Autoformer features a different the temporal modeling mechanism. In standard transformer architectures, sequence dependencies are captured via point-wise Self-Attention, where attention scores are calculated by taking the dot product between individual time steps across the Query (*Q*), Key (*K*), and Value (*V*) matrices [15]. However, point-wise attention is highly susceptible to localized noise and frequently fails to capture the macroscopic periodicity essential for modeling infectious disease outbreaks. To address this, Autoformer replaces standard self-attention with a novel auto-correlation mechanism. Rather than calculating isolated point-to-point similarities, auto-correlation operates at the sub-series level to discover period-based dependencies.

#### Foundation model

Foundation models (FMs) represent a paradigm shift in artificial intelligence [23]. Traditional approaches, such as LSTM and Autoformer, must optimize millions of parameters from scratch on limited local data. In contrast, FMs are pre-trained on massive datasets, enabling them to perform zero-shot inference on new domains without any retraining. While most FMs rely on massive real-world corpora (e.g., text or generic time-series databases), the Prior-Data Fitted Network (TabPFN) is uniquely pre-trained entirely on synthetically generated datasets drawn from a high-dimensional space of Bayesian priors [53]. It was explicitly designed to perform optimal zero-shot inference on extremely small tabular datasets. TabPFN-TS extends this paradigm to temporal data by framing forecasting as a tabular regression task, using historical sequences and temporal covariates as standard tabular features. Operating strictly as a zero-shot forecaster, it approximates Bayesian inference in a single forward pass, mapping the provided context window directly to a predictive distribution for the future horizon:

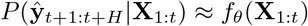

where *f*_*θ*_ represents the pre-trained transformer network operating exclusively in inference mode, and **ŷ** is the resultant multi-step forecast. TabPFN-TS restructures the time-series forecasting problem into a tabular in-context learning (ICL) task. Specifically, the historical lookback window is transformed into a Support Set 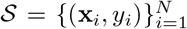, where *y*_*i*_ is the historical incidence and **x**_*i*_ is a constructed feature vector comprising autoregressive lags and exogenous temporal covariates (e.g., the epidemiological week). For a future forecasting horizon *H*, a Query Set 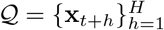 is generated.

#### Models’ ensemble

Every individual model offers distinct advantages, from DLinear’s structural decomposition to TabPFN-TS’s zero-shot capabilities. Despite these strengths, data scarcity and the inherent complexities of the dynamics under study make any single architecture vulnerable to critical forecasting failures. To mitigate architectural bias and enhance overall predictive robustness, we construct a heterogeneous, performance-calibrated ensemble. Rather than assigning naive uniform weights on test sets, our ensemble leverages the historical reliability of each constituent model. For deep learning baselines (LSTM, Autoformer and DLinear), the ensemble first automatically selects the optimal training data strategy (real, augmented, or combined) that minimizes the historical error for each specific country and forecasting horizon. These optimized candidates are then ensembled with our fixed baselines (ARIMA, SEIR and TabPFN-TS). The ensemble weights are calibrated based on the 80% Interval Score (*IS*_80_). Let ℳ = {*m*_1_, *m*_2_, …, *m*_*K*_} denote the set of our *K* selected forecasters. For each model *m*_*k*_, its weight *w*_*k*_ is calculated as the normalized inverse of its historical mean *IS*_80_:

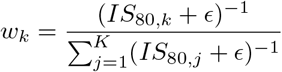

where *ϵ* = 10^−12^ is a strictly positive constant ensuring numerical stability. The final ensemble prediction **ŷ**_*ensemble*_, along with its 80% probabilistic bounds, is computed as the convex combination of the individual multi-step forecasts:

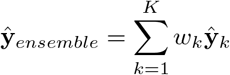

### Prediction interval calibration

To ensure a rigorous and equitable probabilistic evaluation across highly diverse modelling paradigms, we standardized the generation of (1 − *α*) × 100% prediction intervals. Several models within our framework natively output predictive distributions. Specifically, the ARIMA model derives parametric intervals directly from its internal error variance during the fitting process. The TabPFN foundation model outputs predictive distributions through Bayesian inference [21], while the mechanistic models and the RespiCast ensemble [38] inherently provide probabilistic bounds through their respective simulation and multi-model aggregation designs. Conversely, for the point-forecasting architectures, such as the naive baseline and the deep learning models, we implemented a horizon-specific empirical residual calibration strategy inspired by split conformal prediction principles [39, 54, 55]. Instead of imposing restrictive Gaussian assumptions, we evaluated the point forecasts *ŷ*_*t*+*h*_ on a held-out validation set of size *N* to compute the true historical residuals *ϵ*_*t*+*h*_ = *y*_*t*+*h*_ − *ŷ*_*t*+*h*_. Let ℰ_*h*_ = {*ϵ*_1+*h*_, *ϵ*_2+*h*_, …, *ϵ*_*N*+*h*_} denote the set of empirical residuals for a specific prediction horizon *h*. We defined 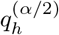 and 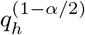 as the *α/*2-th and (1 − *α/*2)-th empirical quantiles of ℰ_*h*_, respectively. The non-parametric (1 − *α*) × 100% prediction interval, denoted as *Č*_*t*+*h*_, for an unseen test observation was then dynamically constructed by adding these horizon-calibrated quantiles to the base point prediction:

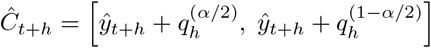

This fully data-driven procedure inherently captures the magnitude-dependent variance and skewness of epidemic trajectories, ensuring rigorous probabilistic evaluation without parametric bounds.

### Evaluation metrics

To rigorously assess both point and probabilistic forecasting performance, predictions were evaluated using three standard epidemiological scoring metrics. The evaluation was stratified by lead time (*h* ∈ {1, 2, 3, 4} weeks). Let *y*_*t*_ denote the true observed incidence and *ŷ*_*t*_ denote the point forecast.

The Mean Absolute Error (MAE) measures the average absolute magnitude of the point forecast errors, providing a straightforward quantification of incidence deviation:

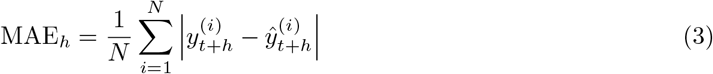

The Weighted Mean Absolute Percentage Error (WMAPE) provides a crucial scale-independent measure of relative accuracy. It weights the absolute errors by the total volume of true observations:

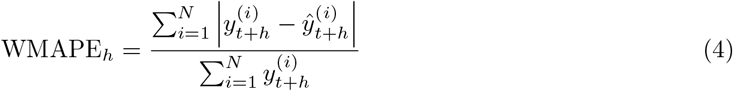

The Interval Score (IS_*α*_) is used to evaluate the calibration and sharpness of the probabilistic forecasts [56, 57]. For a central (1 − *α*) × 100% prediction interval defined by a lower bound *l* and an upper bound *u*, the score is calculated as:

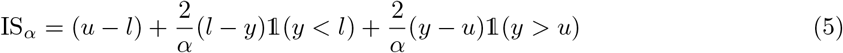

where 𝟙(*·*) is the indicator function. The first term (*u* − *l*) rewards sharpness (narrower intervals), while the subsequent penalty terms rigorously penalize the model when the true observation *y* falls outside the predicted bounds (coverage failure). In our study, we evaluate the 80% prediction interval, thus setting *α* = 0.2. Lower values of IS_80_ indicate superior probabilistic performance.

## Data Availability

The data and code to replicate the results can be found at: https://github.com/DouhanWang/epi4cast

## Supporting information

**S1 Text. Supplementary analyses for the models and results**. In this supplementary file (PDF), we present additional analyses and results of our work.

## Acknowledgments

All authors thank Nicolò Gozzi for useful discussion and the High Performance Computing facilities at Queen Mary University of London. D.W and Y.L. acknowledge support from the China Scholarship Council (CSC). The funders had no role in study design, data collection and analysis, decision to publish, or preparation of the manuscript.

## Author Contributions

D.W. and N.P. designed the research. D.W. implemented and trained all statistical and machine learning models. Y.L. implemented and calibrated the compartmental model. D.W. and N.P. wrote the first draft of the manuscript. All authors contributed interpreting the data, editing and approving the manuscript.

## Data and code

The data and code to replicate the results can be found at: https://github.com/DouhanWang/epi4cast

## References

[1] Madhav Marathe and Anil Kumar S Vullikanti. Computational epidemiology. Communications of the ACM, 56(7):88–96, 2013.

[2] Simon Pollett, Michael A Johansson, Nicholas G Reich, David Brett-Major, Sara Y Del Valle, Srinivasan Venkatramanan, Rachel Lowe, Travis Porco, Irina Maljkovic Berry, Alina Deshpande, et al. Recommended reporting items for epidemic forecasting and prediction research: The epiforge 2020 guidelines. PLoS medicine, 18(10):e1003793, 2021.

[3] John Paget, A Danielle Iuliano, Robert J Taylor, Lone Simonsen, Cecile Viboud, Peter Spreeuwen-berg, et al. Estimates of mortality associated with seasonal influenza for the european union from the glamor project. Vaccine, 40(9):1361–1369, 2022.

[4] European Centre for Disease Prevention and Control (ECDC). Factsheet about seasonal in-fluenza. https://www.ecdc.europa.eu/en/seasonal-influenza/facts/factsheet, 2022. Accessed: 2026-04-25.

[5] Jeffrey Shaman and Alicia Karspeck. Forecasting seasonal outbreaks of influenza. Proceedings of the National Academy of Sciences, 109(50):20425–20430, 2012.

[6] Nicholas G Reich, Logan C Brooks, Spencer J Fox, Sasikiran Kandula, Craig J McGowan, Evan Moore, Dave Osthus, Evan L Ray, Abhinav Tushar, Teresa K Yamana, et al. A collaborative multiyear, multimodel assessment of seasonal influenza forecasting in the united states. Proceedings of the National Academy of Sciences, 116(8):3146–3154, 2019.

[7] James D Munday, Sam Abbott, Sophie Meakin, and Sebastian Funk. Evaluating the use of social contact data to produce age-specific short-term forecasts of sars-cov-2 incidence in england. PLoS Computational Biology, 19(9):e1011453, 2023.

[8] Roy M Anderson and Robert M May. Infectious diseases of humans: dynamics and control. Oxford university press, 1991.

[9] Brian J Coburn, Bradley G Wagner, and Sally Blower. Modeling influenza epidemics and pandemics: insights into the future of swine flu (h1n1). BMC medicine, 7(1):30, 2009.

[10] Matt J Keeling and Pejman Rohani. Modeling infectious diseases in humans and animals. Princeton university press, 2008.

[11] Bryan Lim and Stefan Zohren. Time-series forecasting with deep learning: a survey. Philosophical transactions of the royal society a: mathematical, physical and engineering sciences, 379(2194), 2021.

[12] Andrea Freyer Dugas, Mehdi Jalalpour, Yulia Gel, Scott Levin, Fred Torcaso, Takeru Igusa, and Richard E Rothman. Influenza forecasting with google flu trends. PloS one, 8(2):e56176, 2013.

[13] Xianglei Zhu, Bofeng Fu, Yaodong Yang, Yu Ma, Jianye Hao, Siqi Chen, Shuang Liu, Tiegang Li, Sen Liu, Weiming Guo, et al. Attention-based recurrent neural network for influenza epidemic prediction. BMC bioinformatics, 20(Suppl 18):575, 2019.

[14] Rui Zhang, Zhen Guo, Yujie Meng, Songwang Wang, Shaoqiong Li, Ran Niu, Yu Wang, Qing Guo, and Yonghong Li. Comparison of arima and lstm in forecasting the incidence of hfmd combined and uncombined with exogenous meteorological variables in ningbo, china. International journal of environmental research and public health, 18(11):6174, 2021.

[15] Ashish Vaswani, Noam Shazeer, Niki Parmar, Jakob Uszkoreit, Llion Jones, Aidan N Gomez, Lukasz Kaiser, and Illia Polosukhin. Attention is all you need. Advances in neural information processing systems, 30, 2017.

[16] Haoyi Zhou, Shanghang Zhang, Jieqi Peng, Shuai Zhang, Jianxin Li, Hui Xiong, and Wancai Zhang. Informer: Beyond efficient transformer for long sequence time-series forecasting. In Proceedings of the AAAI conference on artificial intelligence, volume 35, pages 11106–11115, 2021.

[17] Haixu Wu, Jiehui Xu, Jianmin Wang, and Mingsheng Long. Autoformer: Decomposition transformers with auto-correlation for long-term series forecasting. Advances in neural information processing systems, 34:22419–22430, 2021.

[18] Tian Zhou, Ziqing Ma, Qingsong Wen, Xue Wang, Liang Sun, and Rong Jin. Fedformer: Frequency enhanced decomposed transformer for long-term series forecasting. In International conference on machine learning, pages 27268–27286. PMLR, 2022.

[19] Abdul Fatir Ansari, Lorenzo Stella, Caner Turkmen, Xiyuan Zhang, Pedro Mercado, Huibin Shen, Oleksandr Shchur, Syama Sundar Rangapuram, Sebastian Pineda Arango, Shubham Kapoor, et al. Chronos: Learning the language of time series. arXiv preprint arXiv:2403.07815, 2024.

[20] Abhimanyu Das, Weihao Kong, Rajat Sen, and Yichen Zhou. A decoder-only foundation model for time-series forecasting. arXiv preprint arXiv:2310.10688, 2023.

[21] Shi Bin Hoo, Samuel Müller, David Salinas, and Frank Hutter. From tables to time: Extending tabpfn-v2 to time series forecasting. arXiv preprint arXiv:2501.02945, 2025.

[22] Suprabhath Kalahasti, Benjamin Faucher, Boxuan Wang, Claudio Ascione, Ricardo Carbajal, Maxime Enault, Christophe Vincent Cassis, Titouan Launay, Caroline Guerrisi, Pierre-Yves Böelle, Federico Baldo, and Eugenio Valdano. Foundation models for time series forecasting and policy evaluation in infectious disease epidemics. Epidemics, 55:100916, 2026.

[23] Rishi Bommasani, Drew A Hudson, Ehsan Adeli, Russ Altman, Simran Arora, Sydney von Arx, Michael S Bernstein, Jeannette Bohg, Antoine Bosselut, Emma Brunskill, et al. On the opportunities and risks of foundation models. arXiv preprint arXiv:2108.07258, 2021.

[24] Estee Y Cramer, Evan L Ray, Velma K Lopez, Johannes Bracher, Andrea Brennen, Alvaro J Castro Rivadeneira, Aaron Gerding, Tilmann Gneiting, Katie H House, Yuxin Huang, et al. Evaluation of individual and ensemble probabilistic forecasts of covid-19 mortality in the united states. Proceedings of the National Academy of Sciences, 119(15):e2113561119, 2022.

[25] Katharine Sherratt, Hugo Gruson, Rok Grah, Helen Johnson, Rene Niehus, Bastian Prasse, Frank Sandmann, Jannik Deuschel, Daniel Wolffram, Sam Abbott, et al. Predictive performance of multi-model ensemble forecasts of covid-19 across european nations. Elife, 12:e81916, 2023.

[26] Stefania Fiandrino, Andrea Bizzotto, Giorgio Guzzetta, Stefano Merler, Federico Baldo, Eugenio Valdano, Alberto Mateo Urdiales, Antonino Bella, Francesco Celino, Lorenzo Zino, et al. Collaborative forecasting of influenza-like illness in italy: The influcast experience. Epidemics, 50:100819, 2025.

[27] Yuqi Nie, Nam H Nguyen, Phanwadee Sinthong, and Jayant Kalagnanam. A time series is worth 64 words: Long-term forecasting with transformers. arXiv preprint arXiv:2211.14730, 2022.

[28] Yong Liu, Tengge Hu, Haoran Zhang, Haixu Wu, Shiyu Wang, Lintao Ma, and Mingsheng Long. itransformer: Inverted transformers are effective for time series forecasting. arXiv preprint arXiv:2310.06625, 2023.

[29] Inga Holmdahl and Caroline Buckee. Wrong but useful—what covid-19 epidemiologic models can and cannot tell us. New England Journal of Medicine, 383(4):303–305, 2020.

[30] Katharine Sherratt, Rok Grah, Bastian Prasse, Friederike Becker, Jamie McLean, Sam Abbott, and Sebastian Funk. The influence of model structure and geographic specificity on predictive accuracy among european covid-19 forecasts. medRxiv, 2025.

[31] Matthew Biggerstaff, Michael Johansson, David Alper, Logan C. Brooks, Prithwish Chakraborty, David C. Farrow, Sangwon Hyun, Sasikiran Kandula, Craig McGowan, Naren Ramakrishnan, Roni Rosenfeld, Jeffrey Shaman, Rob Tibshirani, Ryan J. Tibshirani, Alessandro Vespignani, Wan Yang, Qian Zhang, and Carrie Reed. Results from the second year of a collaborative effort to forecast influenza seasons in the united states. Epidemics, 24:26–33, 2018.

[32] Abdelhafid Zeroual, Fouzi Harrou, Abdelkader Dairi, and Ying Sun. Deep learning methods for forecasting covid-19 time-series data: A comparative study. Chaos, solitons & fractals, 140:110121, 2020.

[33] Laure Wynants, Ben Van Calster, Gary S Collins, Richard D Riley, Georg Heinze, Ewoud Schuit, Elena Albu, Banafsheh Arshi, Vanesa Bellou, Marc MJ Bonten, et al. Prediction models for diagnosis and prognosis of covid-19: systematic review and critical appraisal. bmj, 369, 2020.

[34] George EP Box, Gwilym M Jenkins, Gregory C Reinsel, and Greta M Ljung. Time series analysis: forecasting and control. John Wiley & Sons, 2015.

[35] William Ogilvy Kermack and Anderson G McKendrick. A contribution to the mathematical theory of epidemics. Proceedings of the royal society of london. Series A, Containing papers of a mathematical and physical character, 115(772):700–721, 1927.

[36] Sepp Hochreiter and Jürgen Schmidhuber. Long short-term memory. Neural computation, 9(8):1735–1780, 1997.

[37] Ailing Zeng, Muxi Chen, Lei Zhang, and Qiang Xu. Are transformers effective for time series forecasting? In Proceedings of the AAAI conference on artificial intelligence, volume 37, pages 11121–11128, 2023.

[38] European Modelling Hubs. Respicast-syndromicindicators: Model output for Respi-Cast hub ensemble (ili/ari 2024/25). https://github.com/european-modelling-hubs/RespiCast-SyndromicIndicators/tree/main/model-output/respicast-hubEnsemble, 2024. Accessed: [2026-04-10].

[39] Anastasios N. Angelopoulos and Stephen Bates. A gentle introduction to conformal prediction and distribution-free uncertainty quantification, 2022.

[40] Amanda Minter and Renata Retkute. Approximate Bayesian Computation for infectious disease modelling. Epidemics, 29:100368, 2019.

[41] Tina Toni, David Welch, Natalja Strelkowa, Andreas Ipsen, and Michael PH Stumpf. Approximate bayesian computation scheme for parameter inference and model selection in dynamical systems. Journal of the Royal Society Interface, 6(31):187–202, 2009.

[42] Max SY Lau, C Jessica E Metcalf, Zewen Liu, Bryan T Grenfell, and Wei Jin. Toward ai foundation models for epidemics: Promise, challenges, and paths forward. Proceedings of the National Academy of Sciences, 123(13):e2526192123, 2026.

[43] Kin On Kwok, Tom Huynh, Wan In Wei, Samuel YS Wong, Steven Riley, and Arthur Tang. Utilizing large language models in infectious disease transmission modelling for public health preparedness. Computational and structural biotechnology journal, 23:3254–3257, 2024.

[44] European Modelling Hubs. Flu forecast hub archive: Target data for ili incidence (seasons 2017-2018, 2018-2019). https://github.com/european-modelling-hubs/flu-forecast-hub_archive/blob/main/target-data/latest-ILI_incidence.csv, 2024. Data sourced from ERVISS. Accessed: [2026-04-10].

[45] European Modelling Hubs. Respicast-syndromicindicators: Target data for ili incidence (seasons 2023-2024, 2024-2025). https://github.com/european-modelling-hubs/RespiCast-SyndromicIndicators/blob/main/target-data/latest-ILI_incidence.csv, 2024. Data sourced from ERVISS. Accessed: [2026-04-10].

[46] Yuchen Qi, Jeffrey Shaman, and Sen Pei. Quantifying the impact of covid-19 nonpharmaceutical interventions on influenza transmission in the united states. The Journal of infectious diseases, 224(9):1500–1508, 2021.

[47] Wenyi Zhang, Yao Wu, Bo Wen, Yongming Zhang, Yong Wang, Wenwu Yin, Shanhua Sun, Xianyu Wei, Hailong Sun, Zhijie Zhang, et al. Non-pharmaceutical interventions for covid-19 reduced the incidence of infectious diseases: a controlled interrupted time-series study. Infectious Diseases of Poverty, 12(02):60–71, 2023.

[48] Shuxuan Song, Qian Li, Li Shen, Minghao Sun, Zurong Yang, Nuoya Wang, Jifeng Liu, Kun Liu, and Zhongjun Shao. From outbreak to near disappearance: how did non-pharmaceutical interventions against covid-19 affect the transmission of influenza virus? Frontiers in Public Health, 10:863522, 2022.

[49] Qingsong Wen, Liang Sun, Fan Yang, Xiaomin Song, Jingkun Gao, Xue Wang, and Huan Xu. Time series data augmentation for deep learning: A survey. arXiv preprint arXiv:2002.12478, 2020.

[50] S Nickbakhsh, F Thorburn, B Von Wissmann, J McMenamin, RN Gunson, and PR Murcia. Extensive multiplex pcr diagnostics reveal new insights into the epidemiology of viral respiratory infections. Epidemiology & Infection, 144(10):2064–2076, 2016.

[51] Dave Osthus. Fast and accurate influenza forecasting in the united states with inferno. PLoS computational biology, 18(1):e1008651, 2022.

[52] Rob J Hyndman and Yeasmin Khandakar. Automatic time series forecasting: the forecast package for r. Journal of statistical software, 27:1–22, 2008.

[53] Noah Hollmann, Samuel Müller, Katharina Eggensperger, and Frank Hutter. Tabpfn: A transformer that solves small tabular classification problems in a second. arXiv preprint arXiv:2207.01848, 2022.

[54] Rob J Hyndman and George Athanasopoulos. Forecasting: principles and practice. OTexts, 2018.

[55] Vladimir Vovk, Alexander Gammerman, and Glenn Shafer. Algorithmic learning in a random world. Springer, 2005.

[56] Tilmann Gneiting and Adrian E Raftery. Strictly proper scoring rules, prediction, and estimation. Journal of the American statistical Association, 102(477):359–378, 2007.

[57] Johannes Bracher, Evan L Ray, Tilmann Gneiting, and Nicholas G Reich. Evaluating epidemic forecasts in an interval format. PLoS computational biology, 17(2):e1008618, 2021.

