## Supplementary Information for "From naive to foundation: benchmarking models for epidemic forecasting"

#### S1. Targets and data overview

Our forecasting target is the weekly incidence of Influenza-Like Illness (ILI) defined as incidence rate per $100,000$ population. Using incidence rates rather than absolute case counts is crucial for our multi-country benchmark, as it normalizes the data against varying national population sizes which supports equitable cross-regional performance comparisons. To achieve this, we used retrospective epidemiological surveillance data from nine geographically diverse European countries: Belgium, Czechia, Denmark, France, Ireland, Italy, Netherlands, Poland, and Romania. The dataset was obtained from the European Respiratory Virus Surveillance Summary (ERVISS) platform, managed by the European Centre for Disease Prevention and Control (ECDC) and the World Health Organization (WHO) Regional Office for Europe. The data for the 2017-2018 and 2018-2019 seasons were obtained from the Flu Forecast Hub archive [1], while the data for the 2023-2024 and 2024-2025 seasons were retrieved from the current RespiCast Syndromic Indicators repository [2] (both originally sourced from the European Respiratory Virus Surveillance Summary, ERVISS). These nine countries were selected to capture diverse spatial transmission dynamics and to ensure direct comparability with the RespiCast ensemble's forecasts [3].

To capture stable and representative transmission dynamics, we constructed our datasets using data from four distinct influenza seasons: 2017-2018, 2018-2019, 2023-2024, and 2024-2025, as illustrated in Figure S1. In accordance with standard epidemiological reporting, each season is defined as starting from ISO week 42 and concluding on ISO week 14 of the subsequent year, safely encapsulating the primary epidemic growth and peak periods. Crucially, surveillance data spanning the COVID-19 pandemic seasons were deliberately excluded from our analysis. The unprecedented non-pharmaceutical interventions (NPIs) implemented during this period caused substantial disruptions to typical seasonal influenza transmission patterns [4–6]. These disruptions introduced structural anomalies and non-stationarity that would confound model training and evaluation.

The surveillance records obtained from ERVISS were historically complete and contained no missing values for the selected seasons which obviates the need for missing data imputation. However, prior to model training, the time-series data underwent rigorous preprocessing to optimize numerical stability. To ensure stable convergence and gradient flow for the deep learning architectures, all historical incidence trajectories were normalized using Standard Scaler (Z-score normalization) before being fed into the neural networks. For augmented datasets, the scaler was strictly fitted on the original, real training data to prevent data leakage from the synthetic trajectories. All predictions were subsequently inverse-transformed to their original scale prior to error evaluation.

*

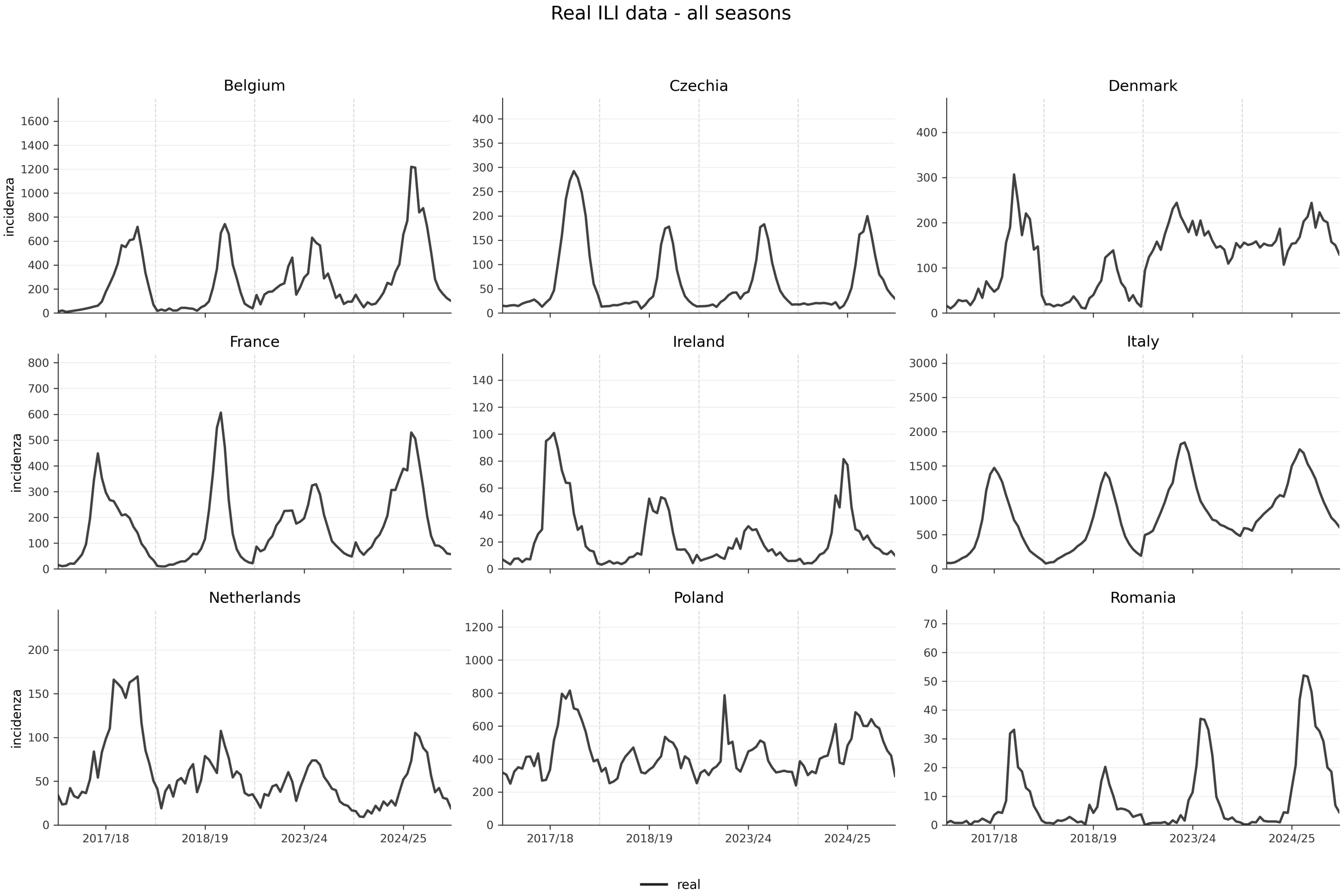


Figure S1: Trajectories of the complete, historical time-series data for all nine countries across the evaluated seasons, illustrating the natural variance in peak timing and incidence magnitude.

### S2. Data augmentation: algorithms and parameters

Deep learning architectures require substantial training data to generalize effectively and avoid over-fitting [7]. However, a standard influenza season yields fewer than 30 observational data points. To alleviate this severe data scarcity and systematically investigate the impact of different data generation paradigms, we developed a comprehensive data augmentation pipeline. We generated distinct streams of synthetic time series to enrich the training space.

#### S2.1 Endogenous augmentation

Our endogenous strategy generates synthetic trajectories by directly augmenting the historical surveillance data. To mimic the natural variability in epidemic peak timing, we applied temporal translations by randomly shifting the empirical epidemic curves by a factor of $k \in [-4, 4]$ weeks. This specific temporal window was empirically derived to reflect the natural cross-regional variance in epidemic peak timing; our analysis of the historical multi-country data showed that peak weeks typically fluctuate within an 8-week span. A standard shift of $\pm 4$ weeks optimally accommodates this variance while maintaining a biologically plausible flu season frame. To prevent artificial boundary discontinuities, missing data points resulting from these temporal shifts were imputed using edge reflection.

Furthermore, to simulate the inherent heteroskedasticity and imperfections in public health reporting processes, we injected a combination of multiplicative and additive Gaussian noise into the shifted trajectories. Specifically, the multiplicative noise was drawn from $\mathcal{N}(0, \sigma^2_{mult})$, where $\sigma_{mult}$ was uniformly sampled from $[0.03, 0.12]$ and scaled by the local incidence magnitude. This noise scale was purposefully calibrated against the empirical surveillance data: across the evaluated countries, the historical incidence exhibited a median week-to-week relative variation consistently above 15% (ranging typically between 15% and 30%). Restricting the synthetic observational noise variance to a maximum of 12% ensures that it will not mask the underlying epidemiological signal. In addition to multiplicative variance, we incorporated an additive noise component scaled to 2% of the seasonal standard deviation. This accounts for the baseline fluctuations inherent in syndromic surveillance—such as the misclassification of other respiratory pathogens during off-peak weeks [8]—without artificially inflating the epidemic baseline. Finally, to maintain biological plausibility, a 3- to 5-week moving average filter was stochastically applied to the augmented trajectories with a 30% probability [9]. While smoothing is standard epidemiological practice to resolve single-week reporting anomalies (e.g., holiday backlogs), applying it stochastically acts as a structural regularizer. This ensures the training pool contains both volatile reporting scenarios and smooth transmission waves, preventing the models from over-fitting to specific noise patterns. The resulting synthetic trajectories generated through this endogenous pipeline plus real data is defined as augmented (aug) setting, as visualized in Figure S2.

#### S2.2 Exogenous augmentation

The second augmentation strategy is instead exogenous and obtained from a stochastic Susceptible-Exposed-Infectious-Recovered (SEIR) compartmental framework. The model was initially calibrated to the empirical incidence of each season via the ABC-SMC (Approximate Bayesian Computation with Sequential Monte Carlo) algorithm. For each country and each season, the algorithm selects 1000 trajectories with the lowest Weighted Mean Absolute Percentage Error (WMAPE) with respect to the data. The resulting synthetic trajectories generated through this exogenous pipeline plus real data is defined as combined (comb) setting, as visualized in Figure S3.

### S3. Model training protocols and forecasting setup

To systematically evaluate the impact of our data enrichment pipelines, we defined three distinct training configurations for the machine learning based models. Models trained only on the historical surveillance data are denoted as *real*. Models trained on the real data supplemented with the synthetic trajectories generated via the statistical pipeline are designated as *augmented* (aug). Conversely, models using the real data combined with the simulated trajectories from the mechanistic SEIR pipeline are referred to as *combined* (comb).

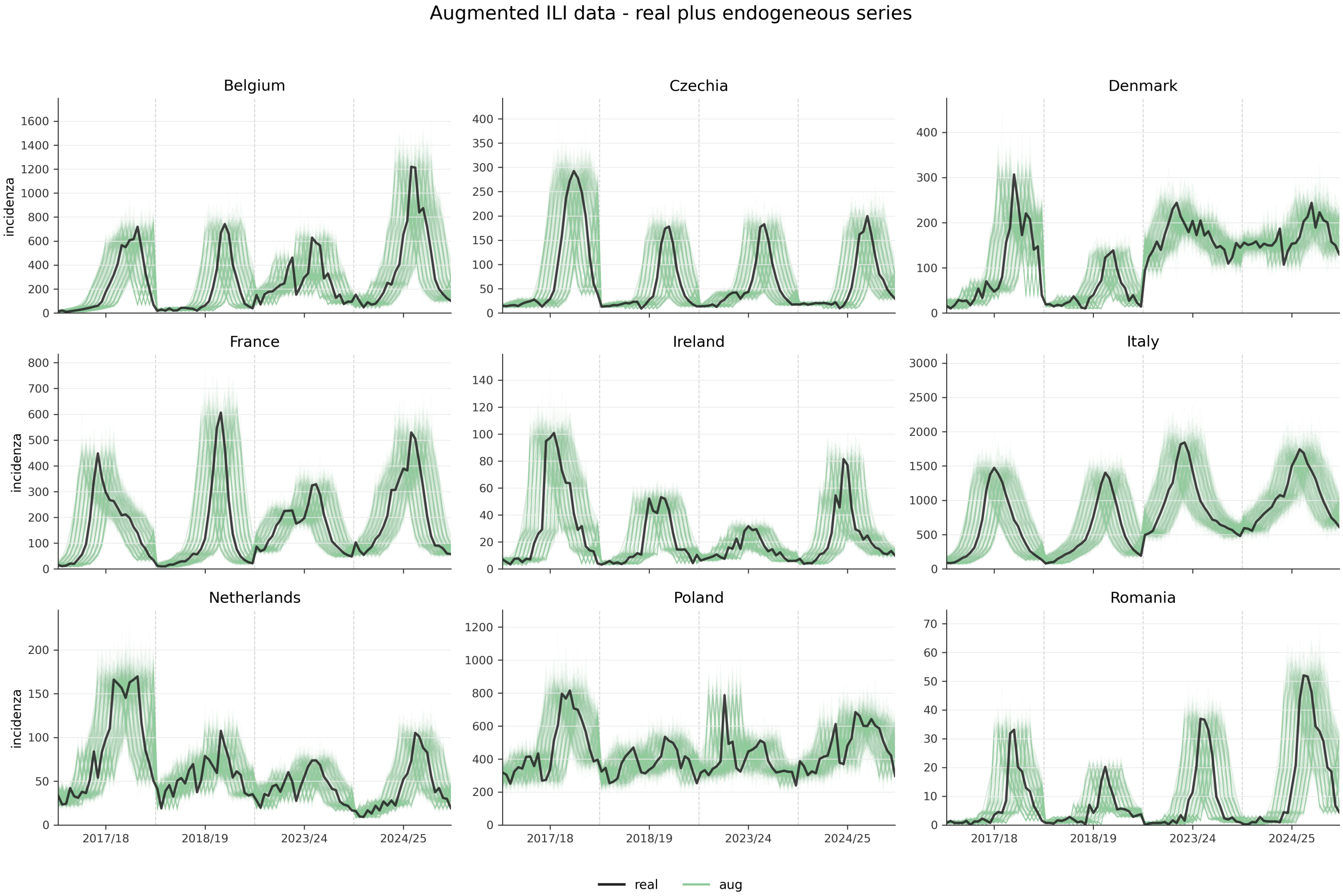


Figure S2: Trajectories of endogenous augmentation for all nine countries across the evaluated seasons, illustrating the natural variance in peak timing and incidence magnitude.

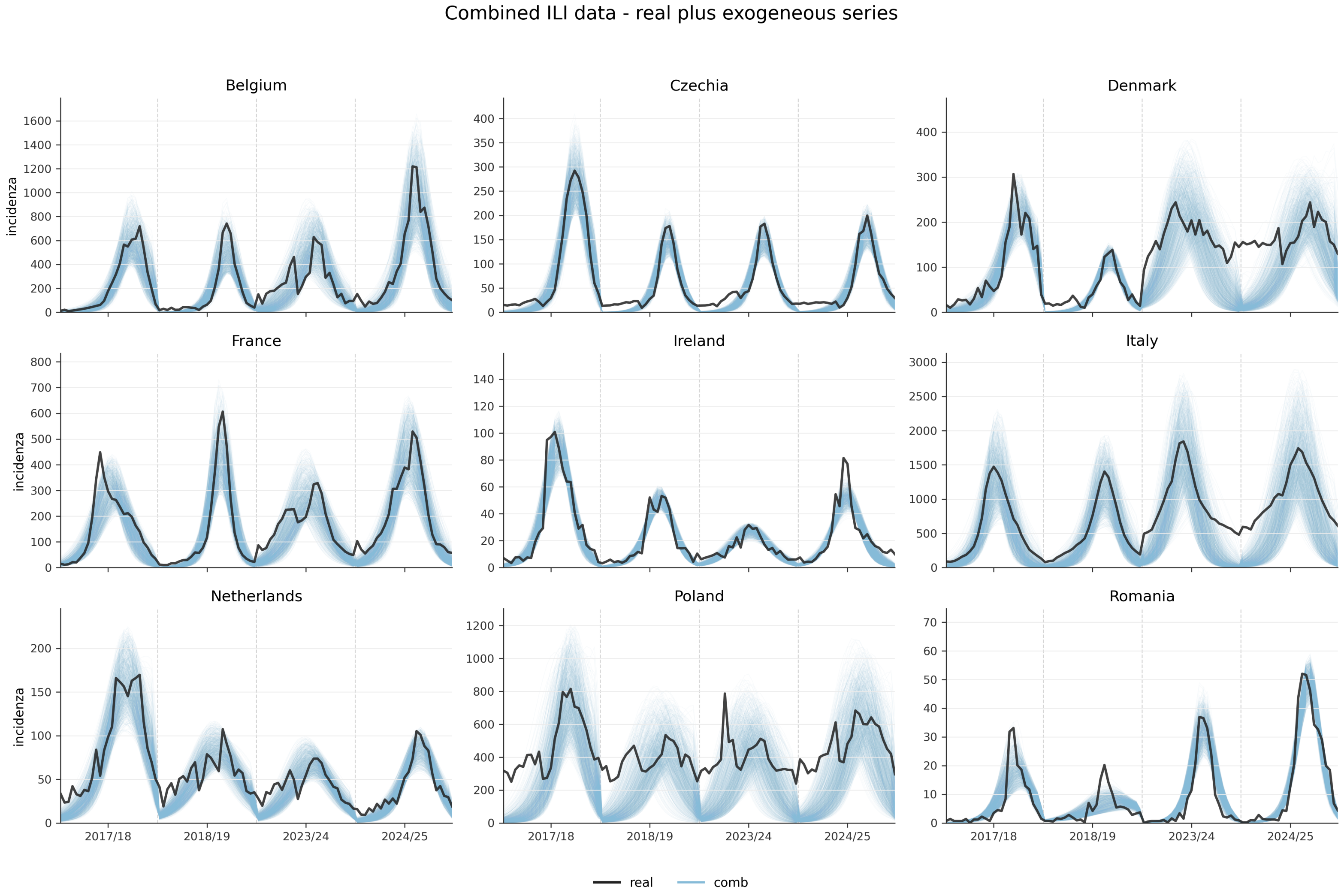


Figure S3: Trajectories of the exogenous augmentation for all nine countries across the evaluated seasons, illustrating the natural variance in peak timing and incidence magnitude.

In the real setting, the 2017–2018, 2018–2019, and 2023–2024 seasons were used for model fitting, while the 2024–2025 season was held out as the unseen test season for final out-of-sample evaluation. Because the real-only setting contains very limited historical seasons, we did not reserve a separate validation season in this configuration. Across all augmented configurations, to rigorously prevent data leakage, the datasets were partitioned such that the 2017–2018 and 2018–2019 seasons, together with their respective synthetic counterparts, served as the training set. The observed raw data from the 2023–2024 season was strictly used as the validation set for hyper-parameter tuning and early stopping, while the 2024–2025 season functioned solely as the unseen test set for the final out-of-sample evaluation.

To simulate a real-time operational forecasting environment, we implemented a rolling-window strategy during the evaluation phase. The forecast origin was initially set at ISO week 45 of each target season. At each weekly step $t$, all models were required to generate a 1- to 4-week-ahead forecast (e.g., predicting the incidence for weeks 46–49 based on information available at week 45). Following each forecast generation, the observation window was sequentially advanced by one week to incorporate the most recent data point, and this process was repeated until the conclusion of the influenza season at ISO week 14.

While the forecasting horizon ($H = 4$) was strictly standardized across all evaluations to ensure equitable comparison, the length of the historical look-back window ($L$) used for inference was tailored to the specific architectural requirements of each method type:

- **Fixed-horizon sliding windows:** The deep learning architectures (LSTM, DLinear, and Autoformer) used a strictly fixed 4-week sliding window ($L = 4$) as their input sequence. This design ensures that these models focus on capturing immediate temporal dependencies and local epidemic gradients.
- **Expanding cumulative windows:** In contrast, methodologies relying on dynamic parameter calibration or extensive in-context learning (ARIMA, SEIR model, and the TabPFN-TS foundation model) employed an expanding cumulative window. This look-back strategy incorporated the entirety of the multi-season historical surveillance record in addition to the observed incidence of the current target season up to the forecast origin $t$. By leveraging the full cumulative history, these models were provided with the maximal epidemiological context necessary for stable parameter fitting or the construction of robust Bayesian support sets.

### S4. Forecasting models and implementation details

To comprehensively evaluate epidemic forecasting capabilities across different methodological paradigms, we employed a diverse taxonomy of models. Let $\mathbf{y}_{1:t} = [y_1, y_2, \ldots, y_t]$ represent the historical incidence sequence, and our goal is to predict the future sequence $\hat{\mathbf{y}}_{t+1:t+H}$, where $H = 4$ is the forecasting horizon.

#### S4.1 Common implementation settings

All models were implemented and evaluated under a unified forecasting framework to ensure direct comparability across methodological families. The target variable was weekly ILI incidence, and all learning-based models used the univariate forecasting setting (`features=S`). The data frequency was weekly (`freq=w`), and all models used the same initial forecast origin after the first 4 observed weeks of the target season. The model-specific look-back strategies followed the fixed-window or expanding-window definitions described in Section S3. The forecasting horizon was set to $H = 4$, corresponding to 1–4 week-ahead forecasts.

For neural-network models, training used a fixed random seed of 2021, the Adam optimizer, and mean squared error (MSE) loss. Models were trained for a maximum of 30 epochs with early stopping where applicable. Unless otherwise specified, the mini-batch size was 32 and the early-stopping patience was 3. The ARIMA and naive baseline used a batch size of 1 and patience of 1, since they do not require iterative neural-network training in the same sense as the deep learning models.

Prediction intervals were standardized to the 80% level across all models. ARIMA and TabPFN-TS produced native predictive intervals, while deterministic point-forecasting models, including naive,

LSTM, DLinear, and Autoformer, used empirical residual quantile calibration to construct 80% prediction intervals. Probabilistic performance was evaluated using the 80% interval score (IS80; $\alpha = 0.2$), while point forecast accuracy was evaluated using MAE and WMAPE.

#### S4.2 Model-specific hyper-parameters

Table S1 summarizes the model-specific architecture and hyper-parameter settings used in the forecasting experiments. Hyper-parameters not listed in the table were kept at their implementation defaults.

Table S1: Model-specific implementation and hyper-parameter settings.

| Model | Hyperparameters / implementation settings |
|---|---|
| Naive | `seq_len`=4; `pred_len`=4; patience=1. |
| ARIMA | Non-seasonal; re-fitted at each rolling forecast origin; $p, q \in [0, 5]$; $d \in [0, 2]$; $p+q \leq 5$; AIC model selection; KPSS differencing test; $\alpha = 0.2$; batch size=1; patience=1. |
| DLinear | `seq_len`=4; `pred_len`=4; `enc_in`=1; moving-average decomposition kernel=3; `individual=False`; batch size=32; learning rate=0.005; epochs=30; patience=3. |
| LSTM | `seq_len`=4; `pred_len`=4; hidden size=512; layers=2; dropout=0.1; batch size=32; learning rate=0.005; epochs=30; patience=3. |
| Autoformer | `seq_len`=4; `label_len`=4; `pred_len`=4; `d_model`=32; `d_ff`=64; heads=2; encoder layers=2; decoder layers=1; moving average=5; factor=3; dropout=0.05; batch size=32; learning rate=0.0005; epochs=30; patience=3. |
| TabPFN-TS | `seq_len`=4; `pred_len`=4; `test_size`=`seq_len`+21; `freq_days`=7; mode=client; features: running index, calendar, auto-seasonal. |
| Ensemble | Weights proportional to $(\text{IS80} + \epsilon)^{-1}$, with $\epsilon = 10^{-12}$; weights calibrated using historical mean IS80. |

#### S4.3 Model architectures

##### Naive model

As a minimal benchmark, the naive model assumes the future incidence remains constant and equal to the last observed value: $\hat{y}_{t+h} = y_t$ for all $h \in [1, H]$. In epidemiological forecasting, this serves as a critical diagnostic to ensure that complex models are genuinely anticipating transmission dynamics rather than merely echoing recent observations. Consequently, the naive model acts as the reference denominator for our relative performance metrics (Relative $IS_{80}$ and Relative Absolute Error).

##### RespiCast ensemble

As the state-of-the-art operational baseline, we included RespiCast: a multi-model forecasting system managed by the European infectious disease hubs [3]. It systematically aggregates outputs from diverse mechanistic and statistical models developed by different teams. For our evaluation, we directly extracted the official 1- to 4-week-ahead predictive quantiles ($10^{th}$ and $90^{th}$ percentiles) generated by the RespiCast hub ensemble for the respective target seasons. To ensure a complete, continuous evaluation and strict comparative parity with our models, particularly because publicly aggregated metrics can occasionally contain missing evaluations for specific target weeks, we independently computed the 80% Interval Score ($IS_{80}$) for the RespiCast predictions using our exact rolling-window target dates and scoring functions. This provides a robust, standardized, and temporally aligned real-world probabilistic benchmark.

##### Mechanistic Model

We adopted a stochastic age-stratified Susceptible-Exposed-Infected-Recovered (SEIR) compartmental model with vaccination [10, 11]. The population is grouped by age and a contact matrix [12] is used for representing the contact patterns among age group. We also consider a seasonal transmission rate. Vaccination was explicitly included in the model framework. For each country and age group, individuals were vaccinated daily according to the average vaccine coverage observed over the previous three seasons. Vaccination was implemented prior to the compartmental transitions, following the dynamics:

$$S_k(t+\delta t) = S_k(t) - \text{Bin}(S_k(t), 1-e^{-\lambda_k(t)}) \tag{1}$$
$$E_k(t+\delta t) = E_k(t) + \text{Bin}(S_k(t), 1-e^{-\lambda_k(t)}) - \text{Bin}(E_k(t), \epsilon) \tag{2}$$
$$I_k(t+\delta t) = I_k(t) + \text{Bin}(E_k(t), \epsilon) - \text{Bin}(I_k(t), \mu) \tag{3}$$
$$R_k(t+\delta t) = R_k(t) + \text{Bin}(I_k(t), \mu) \tag{4}$$

$$S_k^V(t+\delta t) = S_k^V(t) - \text{Bin}(S_k^V(t), 1-e^{-(1-\text{VES})\lambda_k(t)}) \tag{5}$$
$$E_k^V(t+\delta t) = E_k^V(t) + \text{Bin}(S_k^V(t), 1-e^{-(1-\text{VES})\lambda_k(t)}) - \text{Bin}(E_k^V(t), \epsilon) \tag{6}$$
$$I_k^V(t+\delta t) = I_k^V(t) + \text{Bin}(E_k^V(t), \epsilon) - \text{Bin}(I_k^V(t), \mu) \tag{7}$$
$$R_k^V(t+\delta t) = R_k^V(t) + \text{Bin}(I_k^V(t), \mu) \tag{8}$$

where the force of infection is

$$\lambda_k(t) = \beta s(t) \sum_{j=1}^{K} \frac{\mathbf{C}_{kj}(I_j + I_j^V)}{N_j} \tag{9}$$

$\beta$ is the transmission rate computed from the reproductive number $R_0$, $\mathbf{C}$ is the contact matrix. The seasonality factor $s(t)$ modulates the transmission rate to capture seasonal variations in influenza transmissibility:

$$s(t) = s_{\min} + (1 - s_{\min}) \cdot \frac{1 + \cos\left(\frac{2\pi(t - t_{\max})}{365}\right)}{2}, \tag{10}$$

where $s_{\min} \in [0.5, 1]$ is the minimum seasonality scaling factor (calibrated), and $t_{\max}$ denotes the day of peak transmissibility (set to mid-January for the Northern Hemisphere). The factor satisfies $s(t) \in [s_{\min}, 1]$, with $s(t) = 1$ at the seasonal peak and $s(t) = s_{\min}$ at the trough. To avoid issues with transition probabilities large than one, we transform the rates $\lambda_k$ with the function $f(\lambda) = 1 - e^{-\lambda}$. $VES$ is the vaccine efficacy against infections. $Bin(X, p)$ describes a Binomial distribution.

For each country, we calibrate to the historical data in the past three seasons separately using ABC-SMC algorithm [13]. The goal of the ABC-SMC algorithm is to estimate the posterior distribution of the free parameters $\boldsymbol{\theta}$ from a prior distribution $P(\boldsymbol{\theta})$. Parameter sets $\boldsymbol{\theta}_i$ are sampled to generate model outputs $\mathbf{y}_i \sim f(\boldsymbol{\theta}_i)$, which are compared with the observed data $\mathbf{y}_{data}$ (weekly deaths) using a distance metric $d(\mathbf{y}_i, \mathbf{y}_{data})$. Parameters are accepted if the distance is below a tolerance threshold $\xi$. ABC-SMC iteratively updates the parameter distribution over $T$ generations, progressively reducing the tolerance threshold. In each generation, $M$ accepted particles are retained, and the accepted particles from the final generation approximate the posterior distribution. We implemented ABC-SMC using the Python package *pyabc* [14], with $\xi = 0.3$, $T = 10$, and $M = 1000$. We use weighted mean absolute percentage error (wMAPE) as the distance metric. The model's parameters are illustrated in Table S2.

The forecasts are generated using posterior distributions inferred from previous seasons. First, we calibrate the models via the ABC-SMC algorithm described above and obtain 1000 combined posteriors for each season. In order to account for variability across different seasons and uncertainty, we combine the posterior samples from the three past seasons into a single pooled posterior. Then, we draw samples from the combined posterior pool and perform a sufficiently large number (600,000) of stochastic simulations to extensively explore the range of plausible epidemic trajectories. Within each forecasting window, we select the simulated trajectories from the pool via the weighted mean absolute percentage error (wMAPE) between each simulated trajectory and the observed incidence data up to the current forecasting time point, and retain the trajectories with the lowest wMAPE values.

##### Statistical method

As example of traditional statistical framework we considered the AutoRegressive Integrated Moving Average (ARIMA) [17]. It models the differenced stationary time series $y'_t$ (where $d$ is the order of differencing) as a linear combination of its past values and past forecast errors:

$$y'_t = c + \sum_{i=1}^{p} \phi_i y'_{t-i} + \sum_{j=1}^{q} \theta_j \epsilon_{t-j} + \epsilon_t \tag{11}$$

Table S2: SEIR parameters.

| Parameter | Symbol | Value |
|---|---|---|
| Reproductive number | $R_0$ | Calibrated within $Uniform(1, 2)$ [15] |
| Latent period | $\epsilon^{-1}$ | 1.4 days |
| Infectious period | $\mu^{-1}$ | 3.8 days |
| Transmission rate | $\beta$ | Obtained from $R_0$ |
| Contact matrix | $\mathbf{C}$ | Ref. [12] |
| Initial fraction of infections | $i_0$ | Calibrated within $Uniform(5 \times 10^{-6}, 10^{-4})$ |
| Initial fraction of recoveries | $r_0$ | Calibrated within $Uniform(0, 0.5)$ |
| Relative infectivity of vaccinated individuals | $\alpha$ | Calibrated within Uniform(0, 1.0) |
| Adjustment factor for the infection trajectory | $\eta$ | Calibrated within $Uniform(0.01, 100)$ |
| Minimum seasonality scaling factor | $s_{\min}$ | Calibrated within $Uniform(0.5, 1.0)$ |
| Vaccine efficacy against infection | $VE_S$ | 0.4 [16] |
| Age-specific vaccine coverage | $r_k^V$ | Country-specific, from ECDC vaccination data |
| Population in age group $k$ | $N_k$ | Country-specific demographic data |

where $p$ is the autoregressive order, $q$ is the moving average order, $\phi_i$ and $\theta_j$ are the respective parameters, and $\epsilon_t$ is white noise. We used a non-seasonal auto-ARIMA procedure to select $(p, d, q)$ independently at each rolling forecast origin. At each origin, the model was re-fitted using all observations available up to that time and then used to generate forecasts for horizons 1–4 weeks ahead. The candidate orders followed the standard auto-ARIMA search range used in common implementations: $p \in [0, 5]$, $d \in [0, 2]$, and $q \in [0, 5]$, with the additional constraint $p + q \leq 5$. The degree of differencing was selected using the KPSS stationarity test, and the final model order was chosen by minimizing the Akaike Information Criterion (AIC), following the automatic ARIMA selection approach of Hyndman and Khandakar [18]. We used a non-seasonal ARIMA baseline rather than SARIMA because the forecasting data are constructed from influenza-season windows rather than uninterrupted full-year weekly time series. Imposing a fixed seasonal period in SARIMA could therefore introduce artificial seasonal dependence across truncated or non-observed off-season periods.

##### Deep Learning Architectures

**LSTM [19]:** The Long Short-Term Memory network is a specialized Recurrent Neural Network (RNN) designed to overcome the vanishing gradient problem. It uses a gating mechanism (forget, input, and output gates) to regulate the flow of information, allowing the network to capture complex, long-term temporal dependencies in the epidemiological sequence. To adapt this architecture for our multi-horizon evaluation, we implemented a 2-layer LSTM with dropout regularization. Crucially, rather than using a standard autoregressive rollout, we employed a direct multi-step forecasting strategy. Let $h_T \in \mathbb{R}^{d_{model}}$ denote the final hidden state of the encoded historical sequence. To simultaneously predict the entire $H$-week forecasting horizon, $h_T$ is processed through a Layer Normalization block and a linear projection head:

$$\hat{\mathbf{y}}_{t+1:t+H} = \text{Reshape}_{H \times C_{out}}\Big(\text{LayerNorm}(h_T)\mathbf{W} + \mathbf{b}\Big)$$

where $\mathbf{W} \in \mathbb{R}^{d_{model} \times (H \cdot C_{out})}$ and $\mathbf{b} \in \mathbb{R}^{H \cdot C_{out}}$ are the trainable weight and bias of the projection layer, and the output is reshaped to yield the multivariate predictions over the horizon $H$. By this projection, we explicitly mitigated compounding errors, establishing a stronger and more reliable pure-sequence benchmark. Furthermore, to serve as a rigorous pure sequence baseline, this implementation relies exclusively on the historical incidence values, intentionally bypassing the exogenous temporal embeddings (time marks) used in the more complex Transformer-based architectures.

**DLinear [20]:** DLinear is a highly efficient linear architecture optimized for time-series forecasting. It decomposes the input sequence $\mathbf{X}$ into a moving-average trend component ($\mathbf{X}_{trend}$) and a seasonal remainder component ($\mathbf{X}_{seasonal}$). Two single-layer linear networks are then applied separately to these components to forecast the future:

$$\hat{\mathbf{y}} = \mathbf{X}_{trend}\mathbf{W}_{trend} + \mathbf{X}_{seasonal}\mathbf{W}_{seasonal} \tag{12}$$

where $\mathbf{W}_{trend}$ and $\mathbf{W}_{seasonal}$ are the respective weight matrices. By stripping away non-linear activations and self-attention, DLinear enables us to empirically test whether seasonal influenza forecasting requires deep non-linear architectures, or if simple linear decomposition suffices.

**Autoformer [21]:** Autoformer is a state-of-the-art deep learning architecture specifically designed for long-term time-series forecasting, offering significant structural and algorithmic advantages over standard attention-based models. Unlike typical Transformers that treat time series as uniform sequences of tokens, Autoformer assumes that empirical data inherently consists of hidden periodic patterns mixed with macroscopic trends.

To capture this, it introduces a Deep Decomposition Architecture that progressively isolates complex cyclical fluctuations from underlying trend dynamics throughout the network layers. Formally, for a hidden state representation $\mathcal{H}^l$ at the $l$-th layer, the internal series decomposition block extracts the trend-cyclical part $\mathcal{H}^l_{trend}$ and the seasonal part $\mathcal{H}^l_{seasonal}$ via an average pooling operation over a sliding window:

$$\mathcal{H}^l_{trend} = \text{AvgPool}(\mathcal{H}^l), \quad \mathcal{H}^l_{seasonal} = \mathcal{H}^l - \mathcal{H}^l_{trend}$$

Beyond structural decomposition, Autoformer revolutionizes the temporal modeling mechanism. In standard Transformer architectures, sequence dependencies are captured via point-wise Self-Attention, where attention scores are calculated by taking the dot product between individual time steps across the Query ($Q$), Key ($K$), and Value ($V$) matrices [22]. However, point-wise attention is highly susceptible to localized epidemiological noise and frequently fails to capture the macroscopic periodicity essential for modeling infectious disease outbreaks.

To address this, Autoformer replaces standard Self-Attention with a novel Auto-Correlation Mechanism. Rather than calculating isolated point-to-point similarities, Auto-Correlation operates at the sub-series level to discover period-based dependencies. Let $Q, K, V \in \mathbb{R}^{L\times d}$ denote the query, key, and value matrices derived from the seasonal hidden states. Rooted in the Wiener-Khinchin theorem, the mechanism leverages Fast Fourier Transforms ($\mathcal{F}$) to computationally evaluate the temporal correlation $R_{Q,K}(\tau)$ between the Query and Key at all possible time delays $\tau$:

$$S_{Q,K}(f) = \mathcal{F}(Q) \odot \mathcal{F}(K)^*$$

$$R_{Q,K}(\tau) = \mathcal{F}^{-1}(S_{Q,K}(f))$$

where $\odot$ denotes the element-wise multiplication, and $*$ denotes the complex conjugate. This frequency-domain calculation efficiently identifies the most dominant periods within the historical epidemic sequence.

Once the top-$k$ most significant delays (denoted as $\tau_1, \dots, \tau_k$) are identified, the mechanism performs Time Delay Aggregation. It shifts (rolls) the Value matrix $V$ backward by these $\tau$ steps to align past cyclical patterns with the current time step, and aggregates them using the normalized correlation coefficients as weights:

$$\text{Auto-Correlation}(Q, K, V) = \sum_{i=1}^{k} \text{Softmax}(R_{Q,K}(\tau_i)) \cdot \text{Roll}(V, \tau_i)$$

In our study, Autoformer serves as the premier deep representation learning baseline. Unlike our LSTM and DLinear configuration, it uses both historical incidence sequences and exogenous temporal embeddings (time marks) to fully inform its attention layers. By comprehensively evaluating this architecture, we aim to empirically determine whether its internal decomposition and FFT-based macro-attention mechanisms provide a quantifiable advantage over pure linear extrapolation in epidemic forecasting.

##### Foundation Model: TabPFN-TS

Foundation Models (FMs) represent a paradigm shift in artificial intelligence [23]. Traditional baselines, such as our LSTM and Autoformer, must optimize millions of parameters from scratch on limited local data. In contrast, FMs are pre-trained on massive datasets, enabling them to perform zero-shot inference on new domains without any retraining.

While most foundation models rely on massive real-world corpora (e.g., text or generic time-series databases), the Prior-Data Fitted Network (TabPFN [24]) is uniquely pre-trained entirely on synthetically generated datasets drawn from a high-dimensional space of Bayesian priors. It was explicitly designed to perform optimal zero-shot inference on extremely small tabular datasets. TabPFN-TS extends this paradigm to temporal data by framing forecasting as a tabular regression task, using historical sequences and temporal covariates as standard tabular features [25].

Operating strictly as a zero-shot forecaster, it approximates Bayesian inference in a single forward pass, mapping the provided context window directly to a predictive distribution for the future horizon:

$$P(\hat{\mathbf{y}}_{t+1:t+H}|X_{1:t}) \approx f_\theta(X_{1:t})$$

where $f_\theta$ represents the pre-trained Transformer network operating exclusively in inference mode, and $\hat{Y}$ is the resultant multi-step forecast. TabPFN-TS fundamentally restructures the time-series forecasting problem into a tabular In-Context Learning (ICL) task. Specifically, the historical lookback window is transformed into a Support Set $\mathcal{S} = \{(\mathbf{x}_i, y_i)\}_{i=1}^N$, where $y_i$ is the historical incidence and $\mathbf{x}_i$ is a constructed feature vector comprising autoregressive lags and exogenous temporal covariates (e.g., the epidemiological week). For a future forecasting horizon $H$, a Query Set $\mathcal{Q} = \{\mathbf{x}_{t+h}\}_{h=1}^H$ is generated.

Instead of iteratively updating network weights via gradient descent, the frozen Transformer $f_\theta$ directly approximates the Bayesian posterior predictive distribution conditioned on the support set:

$$P(y_{t+h}|\mathbf{x}_{t+h}, \mathcal{S}) \approx f_\theta(\mathbf{x}_{t+h}, \mathcal{S})$$

This mathematical formulation perfectly aligns with the unique challenges of our study. Because our seasonal influenza surveillance spans only four epidemic waves, yielding a total of fewer than 100 historical observations ($N \approx 96$), it represents an extreme data scarcity. Conventional deep learning models attempting to optimize parameters over such a limited dataset are mathematically predisposed to severe overfitting. By replacing gradient-based optimization with forward-pass In-Context Learning, we leverage TabPFN-TS to empirically investigate the zero-shot forecasting capabilities of foundation models.

##### Weighted ensemble

Every individual model offers distinct advantages, from DLinear's structural decomposition to TabPFN-TS's zero-shot capabilities. Despite these strengths, data scarcity and the complexities of the dynamics under study make any single architecture vulnerable to critical forecasting failures. To mitigate architectural bias and enhance overall predictive robustness, we construct a heterogeneous, performance-calibrated ensemble.

Rather than assigning naive uniform weights on test sets, our ensemble strategically leverages the historical reliability of each constituent model. For deep learning baselines (LSTM, Autoformer and DLinear), the ensemble first automatically selects the optimal training data strategy (real, augmented, or combined) that minimizes the historical error for each specific country and forecasting horizon. These optimized candidates are then ensembled with our fixed baselines (ARIMA, SEIR and TabPFN-TS).

Crucially, the ensemble weights are calibrated based on the 80% Interval Score ($IS_{80}$), a rigorous epidemiological standard that simultaneously evaluates point accuracy and probabilistic uncertainty (balancing the sharpness of the prediction interval against calibration penalties; the formal mathematical definition is provided in the Evaluation Metrics section). Let $\mathcal{M} = \{m_1, m_2, \ldots, m_K\}$ denote the set of our $K$ selected forecasters. For each model $m_k$, its weight $w_k$ is calculated as the normalized inverse of its historical mean $IS_{80}$:

$$w_k = \frac{(IS_{80,k} + \epsilon)^{-1}}{\sum_{j=1}^{K}(IS_{80,j} + \epsilon)^{-1}}$$

where $\epsilon = 10^{-12}$ is a strictly positive constant ensuring numerical stability. The final ensemble prediction $\hat{Y}_{ensemble}$, along with its 80% probabilistic bounds, is computed as the convex combination of the individual multi-step forecasts:

$$\hat{\mathbf{y}}_{ensemble} = \sum_{k=1}^{K} w_k \hat{\mathbf{y}}_k$$

By fixing these empirically calibrated weights during the testing phase, the ensemble acts as a highly stabilized aggregation scheme. It explicitly rewards models that provide sharp, well-calibrated probabilistic distributions, effectively suppressing the volatile outputs of architectures that fail to generalize.

#### S4.4 Prediction interval calibration

To ensure a rigorous and equitable probabilistic evaluation across our highly diverse modelling paradigms, we standardized the generation of $(1-\alpha) \times 100\%$ prediction intervals.

Several models within our framework natively output predictive distributions. Specifically, the ARIMA model derives parametric intervals directly from its internal error variance during the fitting process. The TabPFN foundation model outputs predictive distributions through Bayesian inference [25], while the mechanistic models and the RespiCast ensemble [3] inherently provide probabilistic bounds through their respective simulation and multi-model aggregation designs.

The SEIR model generates probabilistic forecasts by leveraging its stochastic simulation design. Rather than producing a purely deterministic output, the model executes 1,000 independent simulations calibrated via Approximate Bayesian Computation (ABC). These trajectories inherently account for structural epidemiological uncertainties. We derive the $(1-\alpha) \times 100\%$ prediction intervals directly from the empirical distribution of these 1,000 simulated trajectories

**Empirical residual calibration:**
Conversely, for the point-forecasting architectures, such as the naive baseline and the deep learning models (DLinear, LSTM, Autoformer), we implemented a horizon-specific empirical residual calibration strategy inspired by split conformal prediction principles [26–28]. Instead of imposing restrictive Gaussian assumptions, we evaluated the point forecasts $\hat{y}_{t+h}$ on a held-out validation set of size $N$ to compute the true historical residuals $\epsilon_{t+h} = y_{t+h} - \hat{y}_{t+h}$. Let $\mathcal{E}_h = \{\epsilon_{1+h}, \epsilon_{2+h}, \ldots, \epsilon_{N+h}\}$ denote the set of empirical residuals for a specific prediction horizon $h$. We defined $q_h^{(\alpha/2)}$ and $q_h^{(1-\alpha/2)}$ as the $\alpha/2$-th and $(1-\alpha/2)$-th empirical quantiles of $\mathcal{E}_h$, respectively. The non-parametric $(1-\alpha) \times 100\%$ prediction interval, denoted as $\hat{C}_{t+h}$, for an unseen test observation was then dynamically constructed by adding these horizon-calibrated quantiles to the base point prediction:

$$\hat{C}_{t+h} = \left[\hat{y}_{t+h} + q_h^{(\alpha/2)},\ \hat{y}_{t+h} + q_h^{(1-\alpha/2)}\right]$$

This fully data-driven procedure inherently captures the magnitude-dependent variance and skewness of epidemic trajectories, ensuring rigorous probabilistic evaluation without parametric bounds.

### S5. Evaluation metrics

To rigorously assess both point and probabilistic forecasting performance, predictions were evaluated using three standard epidemiological scoring rules. The evaluation was stratified by lead time ($h \in \{1, 2, 3, 4\}$ weeks). Let $y_t$ denote the true observed incidence and $\hat{y}_t$ denote the point forecast.

**Mean Absolute Error (MAE):** MAE measures the average absolute magnitude of the point forecast errors, providing a straightforward quantification of incidence deviation:

$$\text{MAE}_h = \frac{1}{N}\sum_{i=1}^{N}\left|y_{t+h}^{(i)} - \hat{y}_{t+h}^{(i)}\right| \tag{13}$$

**Weighted Mean Absolute Percentage Error (WMAPE):** Given the substantial variation in baseline population and infection scale across the nine European countries, WMAPE provides a crucial scale-independent measure of relative accuracy. It weights the absolute errors by the total volume of true observations:

$$\text{WMAPE}_h = \frac{\sum_{i=1}^{N}\left|y_{t+h}^{(i)} - \hat{y}_{t+h}^{(i)}\right|}{\sum_{i=1}^{N} y_{t+h}^{(i)}} \tag{14}$$

**Interval Score (IS$_\alpha$):** To evaluate the calibration and sharpness of the probabilistic forecasts, we use the Interval Score, a strictly proper scoring rule widely adopted in epidemiological forecasting hubs[29, 30]. For a central $(1-\alpha)\times 100\%$ prediction interval defined by a lower bound $l$ and an upper bound $u$, the score is calculated as:

$$\text{IS}_\alpha = (u-l) + \frac{2}{\alpha}(l-y)\mathbb{1}(y<l) + \frac{2}{\alpha}(y-u)\mathbb{1}(y>u) \tag{15}$$

where $\mathbb{1}(\cdot)$ is the indicator function. The first term $(u-l)$ rewards sharpness (narrower intervals), while the subsequent penalty terms rigorously penalize the model when the true observation $y$ falls outside the predicted bounds (coverage failure). In our study, we evaluate the 80% prediction interval, thus setting $\alpha = 0.2$. Lower values of IS$_{80}$ indicate superior probabilistic performance.

### S6. Extended results

In the main text, we primarily focus our discussion on the 4-week-ahead forecasts (Horizon 4), where the best-performing alternative model for each country was selected based on the lowest Mean Absolute Error (MAE). To complement this with a probabilistic perspective, we present the Horizon 4 trajectory forecasts in Figure S4, where the best alternative models are instead identified based on the lowest Interval Score ($IS_{80}$). This metric change may alter the country-specific model selected for comparison, because $IS_{80}$ jointly rewards sharp predictive intervals and penalizes coverage failures rather than evaluating point accuracy alone. Compared with the MAE-based trajectory comparison in Figure 4 of the main text, selecting the best alternative by $IS_{80}$ in Figure S4 changes the highlighted model in several countries: Belgium and France shift from SEIR to TabPFN-TS, Denmark from TabPFN-TS to DLinear (aug), Poland from DLinear (aug) to LSTM (real), and Romania from SEIR to LSTM (aug). Czechia remains best represented by the weighted ensemble, while Ireland, Italy, and the Netherlands remain best represented by TabPFN-TS. These changes indicate that the model with the lowest point error is not always the one with the best-calibrated and sharpest uncertainty intervals. Importantly, however, the overall interpretation remains consistent: TabPFN-TS is still frequently selected under the probabilistic criterion, confirming that its strong zero-shot performance extends to uncertainty quantification at the 4-week-ahead horizon.

To provide a comprehensive evaluation of model behaviour across all timescales, we also present the extended quantitative results for all shorter horizons. Figures S5 to S10 illustrate the distribution of the Relative Interval Score ($IS_{80}$) and the Relative Absolute Error (RAE) across all nine European countries for Steps 1, 2, and 3. Consistent with the Horizon 4 results, these figures demonstrate how the relative advantage of different methodological paradigms shifts as the forecasting window extends. Furthermore, to complement these relative performance distributions, Table S3 provides the comprehensive Weighted Mean Absolute Percentage Error (WMAPE) for all evaluated models across the complete set of countries and forecasting horizons (Steps 1 through 4). As detailed in Section S5, WMAPE serves as a crucial scale-independent measure of relative accuracy.

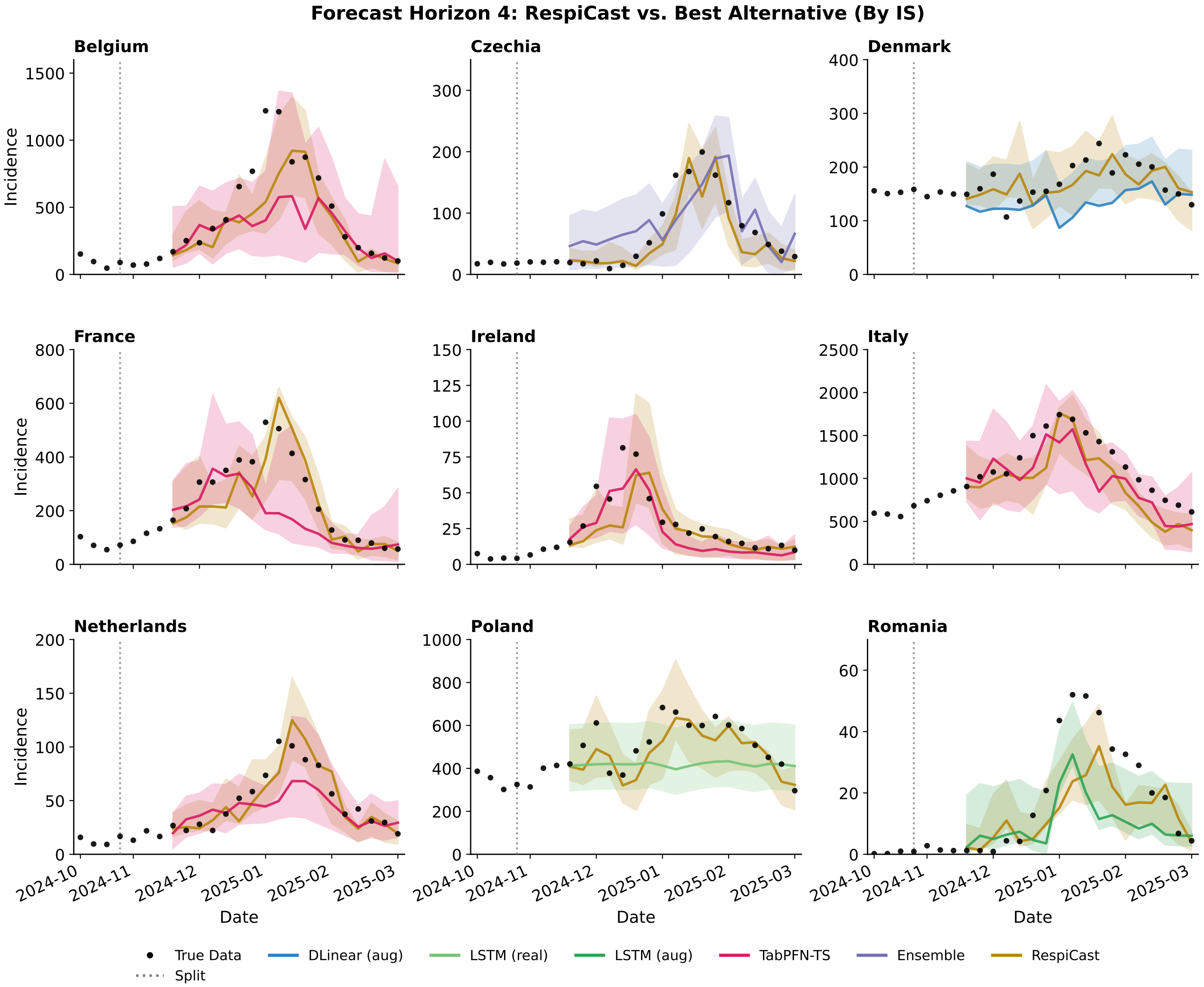


Figure S4: Epidemic trajectory forecasts at Horizon 4 (4-week-ahead) across nine European countries during the 2024-2025 influenza season. The plots visually compare the predictions of the RespiCast ensemble against the best-performing alternative model for each respective country, selected based on the lowest mean Interval Score ($IS_{80}$). Black dots represent the true observed weekly incidence of Influenza-Like Illness (ILI). The vertical dashed line marks the forecast origin (split). Solid lines denote the point forecasts, while the corresponding shaded regions represent the 80% prediction intervals, illustrating the models' capacity to capture complex epidemic peaks and quantify uncertainty.

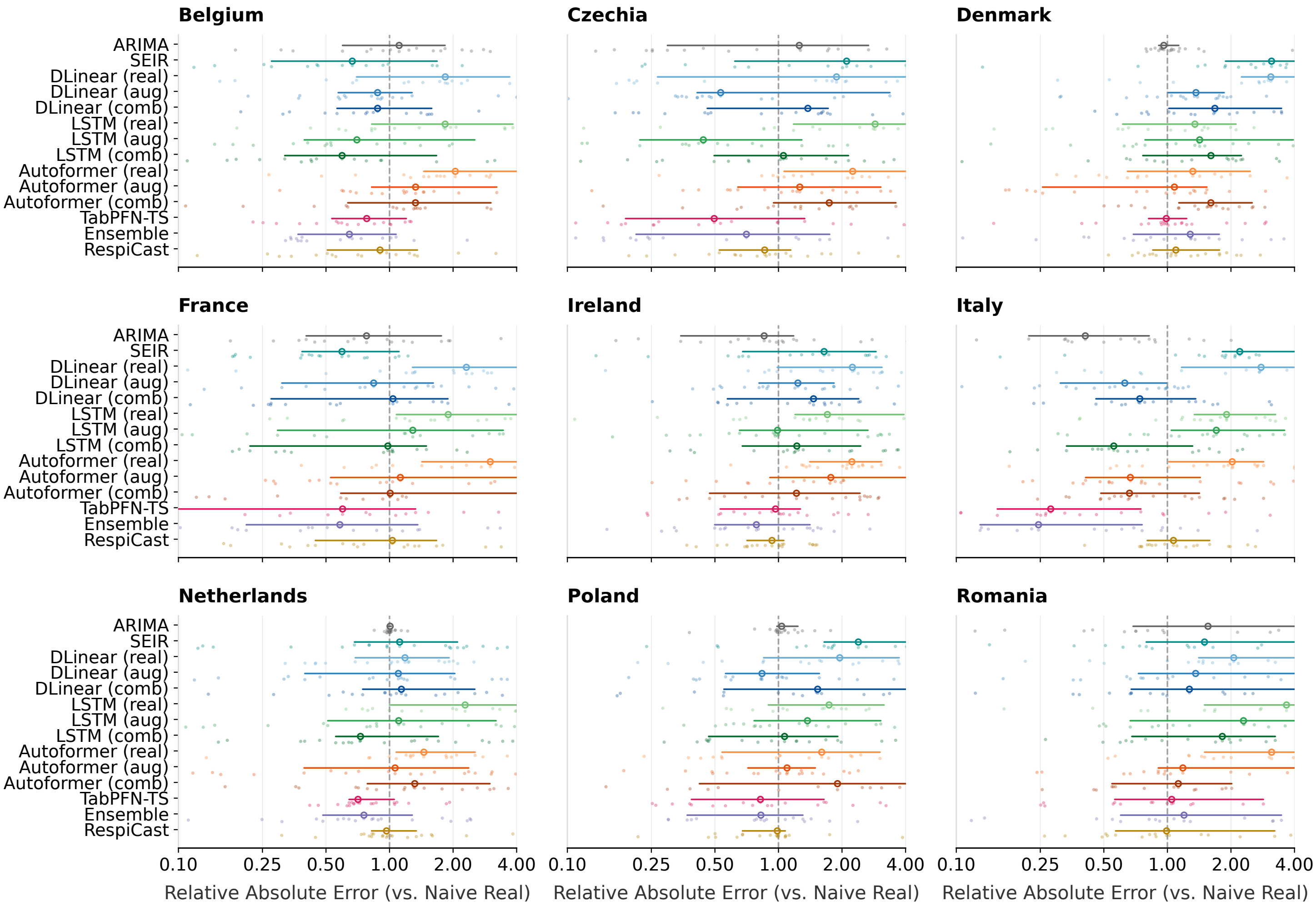


Figure S5: **Point forecast accuracy at Horizon 1 (i.e., 1-week-ahead).** Distribution of the Relative Absolute Error (Relative AE) across nine European countries. The vertical dashed line at 1.0 marks the naive baseline; values < 1.0 indicate a reduction in absolute prediction error. For deep learning models, suffixes denote training data strategies: **(real)** for real data only, **(aug)** for endogenous data, and **(comb)** for exogenous data. The 'Ensemble' strategy combines the best-performing methods across all respective data configurations. The x-axis is on a logarithmic scale.

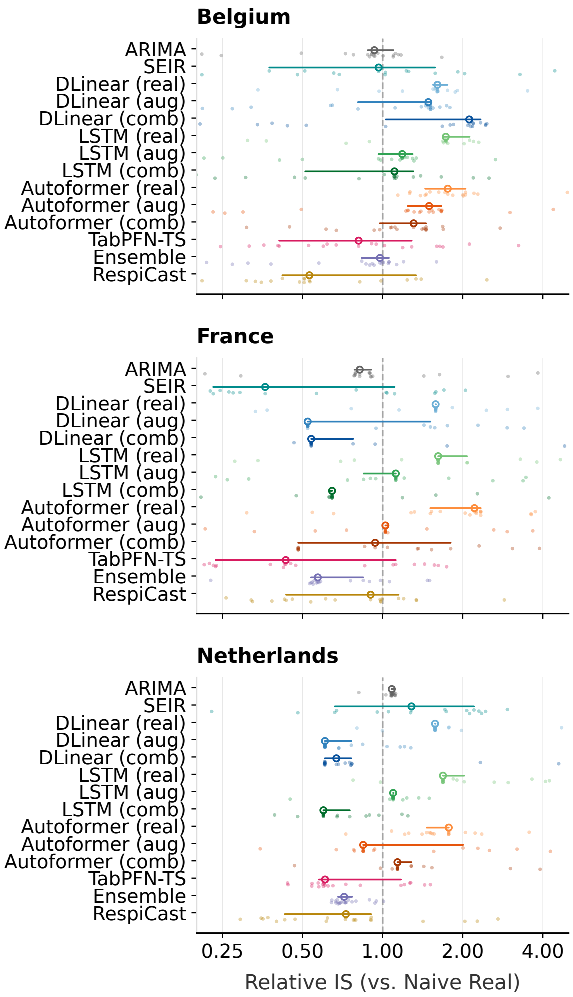


Figure S6: **Probabilistic forecasting performance at Horizon 1 (i.e., 1-week-ahead).** Distribution of the Relative Interval Score (Relative IS) across nine European countries. The vertical dashed line at 1.0 represents the naive baseline; values $< 1.0$ indicate improved probabilistic accuracy. Model suffix definitions and plotting conventions are identical to those in Fig. S5. The 'Ensemble' strategy combines the best-performing methods across all respective data configurations. The x-axis is on a logarithmic scale.

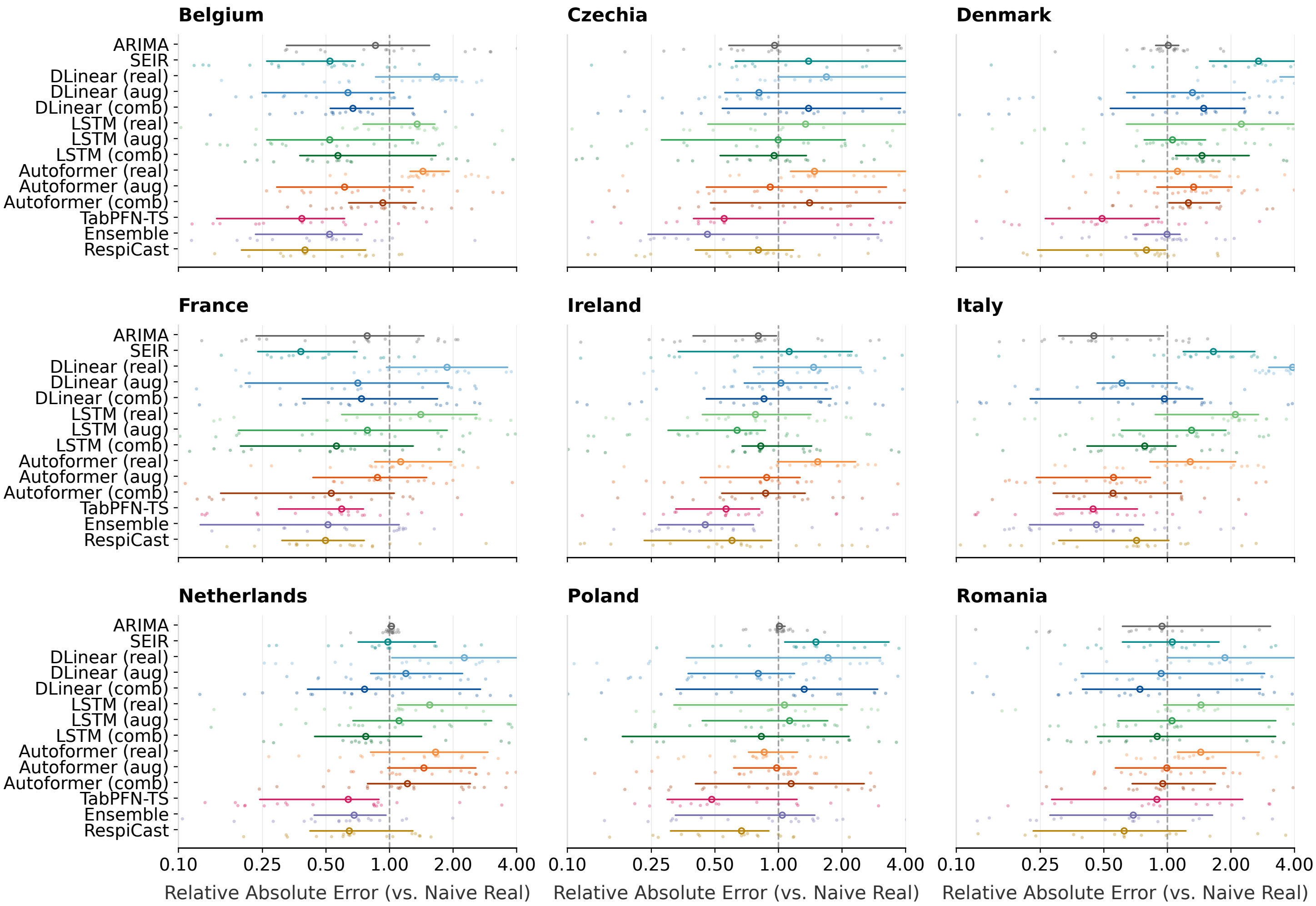


Figure S7: **Point forecast accuracy at Horizon 2 (i.e., 2-week-ahead).** Distribution of the Relative Absolute Error (Relative AE) across nine European countries. The vertical dashed line at 1.0 marks the naive baseline; values < 1.0 indicate a reduction in absolute prediction error. Model suffix definitions and plotting conventions are identical to those in Fig. S5. The 'Ensemble' strategy combines the best-performing methods across all respective data configurations. The x-axis is on a logarithmic scale.

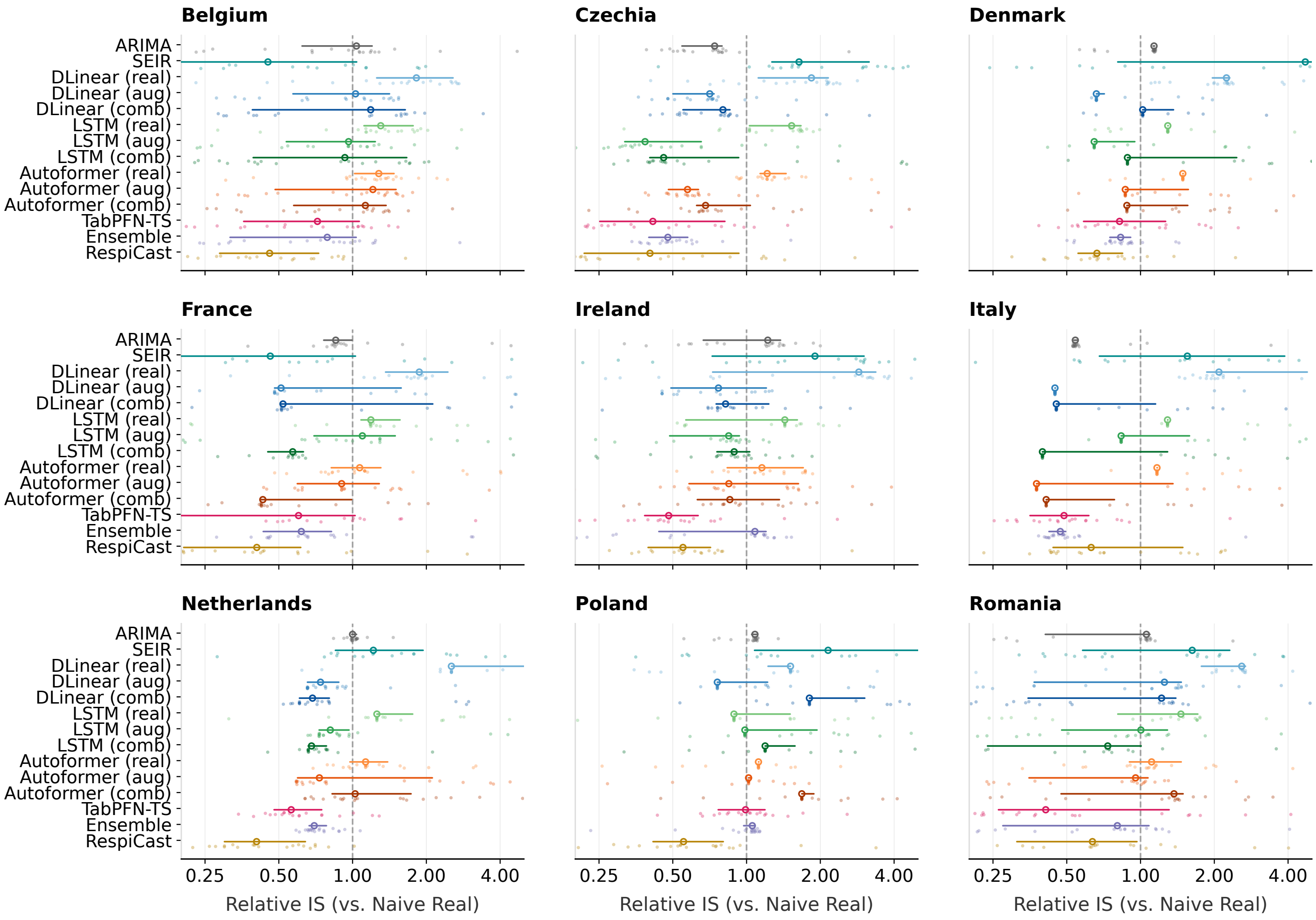


Figure S8: **Probabilistic forecasting performance at Horizon 2 (i.e., 2-week-ahead).** Distribution of the Relative Interval Score (Relative IS) across nine European countries. The vertical dashed line at 1.0 represents the naive baseline; values < 1.0 indicate improved probabilistic accuracy. Model suffix definitions and plotting conventions are identical to those in Fig. S5. The 'Ensemble' strategy combines the best-performing methods across all respective data configurations. The x-axis is on a logarithmic scale.

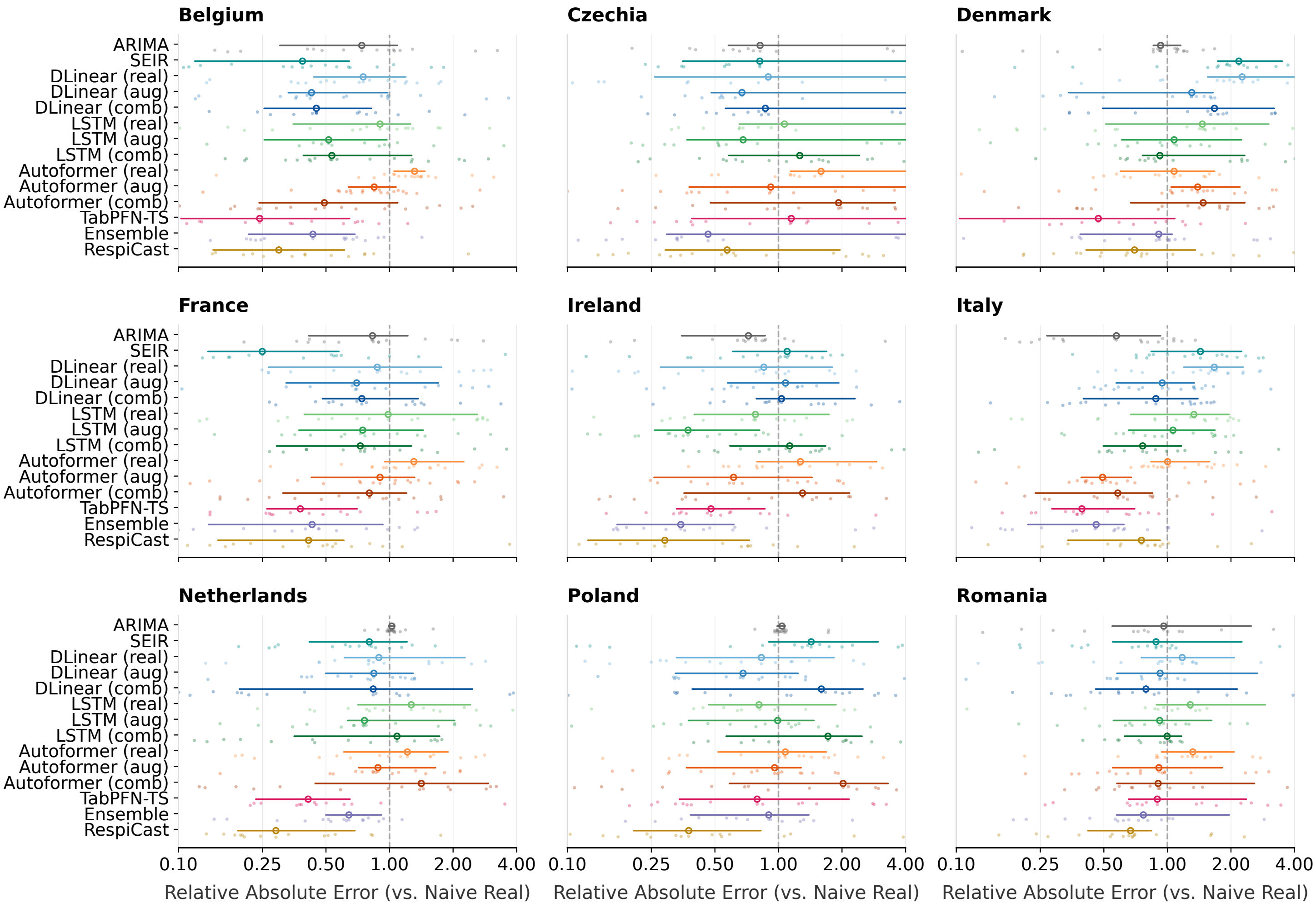


Figure S9: **Point forecast accuracy at Horizon 3 (i.e., 3-week-ahead).** Distribution of the Relative Absolute Error (Relative AE) across nine European countries. The vertical dashed line at 1.0 marks the naive baseline; values < 1.0 indicate a reduction in absolute prediction error. Model suffix definitions and plotting conventions are identical to those in Fig. S5. The 'Ensemble' strategy combines the best-performing methods across all respective data configurations. The x-axis is on a logarithmic scale.

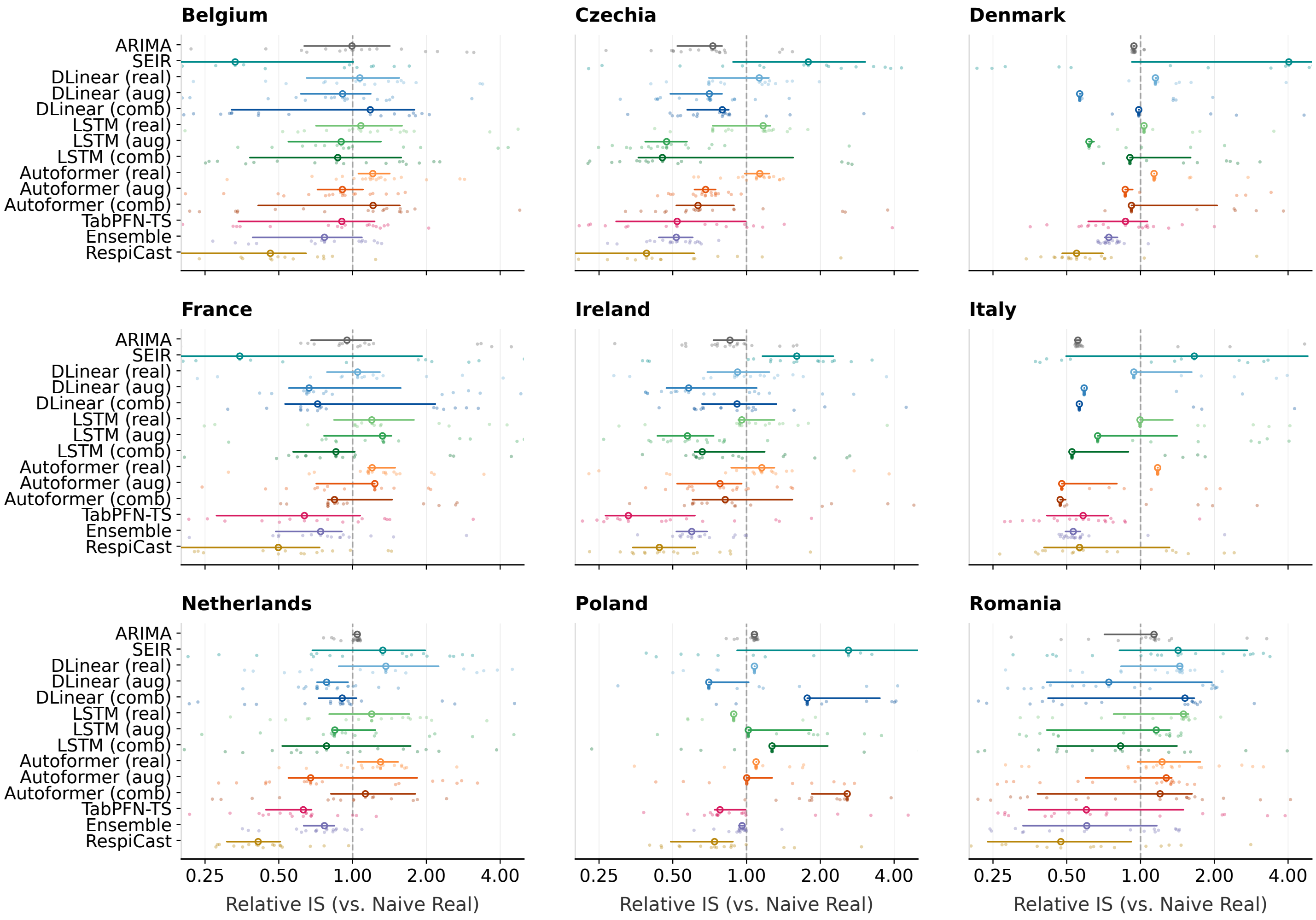


Figure S10: **Probabilistic forecasting performance at Horizon 3 (i.e., 3-week-ahead).** Distribution of the Relative Interval Score (Relative IS) across nine European countries. The vertical dashed line at 1.0 represents the naive baseline; values < 1.0 indicate improved probabilistic accuracy. Model suffix definitions and plotting conventions are identical to those in Fig. S5. The 'Ensemble' strategy combines the best-performing methods across all respective data configurations. The x-axis is on a logarithmic scale.

| Country | Step | Naive | ARIMA | SEIR | DLinear | | | LSTM | | | Autoformer | | | TabPFN_ts | Respicast | Ensemble |
|---|---|---|---|---|---|---|---|---|---|---|---|---|---|---|---|---|
| | | | | | real | aug | combined | real | aug | combined | real | aug | combined | | | |
| Belgium | 1 | 0.2561 | 0.2413 | 0.2102 | 0.3735 | 0.2366 | 0.3798 | 0.5105 | 0.2716 | **0.2001** | 0.5203 | 0.3559 | 0.3878 | 0.2257 | 0.3146 | 0.2156 |
| | 2 | 0.4865 | 0.4195 | 0.2574 | 0.8665 | 0.3882 | 0.5403 | 0.5934 | 0.3657 | 0.3085 | 0.6429 | 0.3717 | 0.5294 | **0.2318** | 0.296 | 0.2748 |
| | 3 | 0.6452 | 0.5041 | 0.2987 | 0.6542 | 0.4386 | 0.5932 | 0.609 | 0.4184 | 0.4088 | 0.7929 | 0.4844 | 0.4154 | 0.2813 | **0.2698** | 0.2968 |
| | 4 | 0.8114 | 0.6046 | 0.3147 | 0.8653 | 0.4768 | 0.6137 | 0.6197 | 0.4551 | 0.4616 | 0.936 | 0.5748 | 0.617 | 0.3747 | **0.2743** | 0.3593 |
| Czechia | 1 | 0.2756 | 0.228 | 0.3183 | 0.4606 | 0.2121 | 0.2741 | 0.5494 | 0.1894 | 0.2348 | 0.5806 | 0.3517 | 0.3942 | **0.137** | 0.248 | 0.1742 |
| | 2 | 0.4848 | 0.3743 | 0.4867 | 0.8604 | 0.3722 | 0.4171 | 0.5809 | 0.2768 | 0.4172 | 0.6844 | 0.3096 | 0.4514 | 0.2909 | 0.2792 | **0.2212** |
| | 3 | 0.6757 | 0.5527 | 0.6346 | 0.6423 | 0.5335 | 0.5896 | 0.6809 | 0.4445 | 0.6091 | 0.8858 | 0.4019 | 0.5736 | 0.5158 | **0.2986** | 0.3466 |
| | 4 | 0.8659 | 0.627 | 0.7733 | 0.8077 | 0.6321 | 0.7113 | 0.665 | 0.6166 | 0.7757 | 0.9246 | 0.5459 | 0.6271 | 0.7033 | **0.3167** | 0.5209 |
| Denmark | 1 | 0.1291 | 0.1292 | 0.371 | 0.3055 | 0.1432 | 0.214 | 0.1462 | 0.2027 | 0.1583 | 0.1737 | 0.1545 | 0.1952 | **0.1193** | 0.1387 | 0.1605 |
| | 2 | 0.171 | 0.173 | 0.4296 | 0.8669 | 0.1872 | 0.3272 | 0.2806 | 0.1898 | 0.2469 | 0.1898 | 0.2248 | 0.2236 | **0.1191** | 0.1264 | 0.1563 |
| | 3 | 0.208 | 0.2098 | 0.4976 | 0.4719 | 0.2169 | 0.3785 | 0.2427 | 0.2286 | 0.2853 | 0.2558 | 0.3113 | 0.2992 | **0.1225** | 0.1369 | 0.1581 |
| | 4 | 0.2236 | 0.2325 | 0.5602 | 0.2441 | 0.2606 | 0.3841 | 0.2654 | 0.2538 | 0.2832 | 0.248 | 0.2645 | 0.4061 | **0.1204** | 0.1565 | 0.1697 |
| France | 1 | 0.1917 | 0.1523 | 0.1629 | 0.3526 | 0.1453 | 0.2039 | 0.3929 | 0.2197 | 0.1669 | 0.4304 | 0.2572 | 0.2135 | 0.1298 | 0.2111 | **0.1265** |
| | 2 | 0.3715 | 0.279 | 0.1887 | 0.8152 | 0.2612 | 0.3589 | 0.5004 | 0.3373 | 0.2756 | 0.4556 | 0.2968 | 0.2508 | 0.2098 | 0.2024 | **0.1691** |
| | 3 | 0.5147 | 0.4085 | 0.2221 | 0.5589 | 0.3584 | 0.4582 | 0.5294 | 0.4462 | 0.3721 | 0.6819 | 0.4482 | 0.4723 | 0.2998 | **0.2156** | 0.2261 |
| | 4 | 0.6478 | 0.495 | 0.2552 | 0.7766 | 0.4333 | 0.4763 | 0.5586 | 0.5059 | 0.4386 | 0.7932 | 0.6039 | 0.6714 | 0.3596 | **0.2379** | 0.2602 |
| Ireland | 1 | 0.3081 | 0.2672 | 0.431 | 0.4668 | 0.3201 | 0.486 | 0.4511 | 0.3348 | 0.3337 | 0.6122 | 0.4506 | 0.2967 | 0.2602 | 0.3352 | **0.2582** |
| | 2 | 0.5031 | 0.3864 | 0.5211 | 0.7348 | 0.4489 | 0.6424 | 0.4506 | 0.3102 | 0.5164 | 0.6038 | 0.4271 | 0.4495 | **0.2773** | 0.3298 | 0.288 |
| | 3 | 0.6325 | 0.4362 | 0.6478 | 0.5306 | 0.5267 | 0.7706 | 0.5018 | 0.284 | 0.5988 | 0.753 | 0.3903 | 0.6577 | 0.2733 | 0.3206 | **0.2483** |
| | 4 | 0.8016 | 0.5093 | 0.7607 | 0.7998 | 0.5965 | 0.8376 | 0.54 | 0.386 | 0.7815 | 0.9381 | 0.5253 | 0.8048 | **0.2983** | 0.3326 | 0.3366 |
| Italy | 1 | 0.09494 | 0.05151 | 0.2794 | 0.2849 | 0.06138 | 0.061 | 0.22 | 0.1792 | 0.0623 | 0.2333 | 0.07996 | 0.09372 | 0.05157 | 0.1111 | **0.05012** |
| | 2 | 0.1817 | 0.1066 | 0.3564 | 0.8308 | 0.1364 | 0.1217 | 0.3403 | 0.2488 | 0.1335 | 0.2393 | 0.1221 | 0.12 | 0.08847 | 0.1275 | **0.08748** |
| | 3 | 0.2642 | 0.1628 | 0.4367 | 0.4727 | 0.2556 | 0.2118 | 0.3561 | 0.3416 | 0.2113 | 0.3036 | 0.1698 | 0.155 | **0.1323** | 0.1698 | 0.1377 |
| | 4 | 0.3385 | 0.2292 | 0.516 | 0.3992 | 0.3825 | 0.3682 | 0.3753 | 0.4301 | 0.3261 | 0.3973 | 0.1992 | 0.1925 | **0.1869** | 0.2197 | 0.2082 |
| Netherlands | 1 | 0.2308 | 0.23 | 0.2591 | 0.2876 | 0.2472 | 0.26 | 0.5093 | 0.3103 | 0.202 | 0.4163 | 0.2881 | 0.3517 | **0.1874** | 0.2544 | 0.1943 |
| | 2 | 0.3506 | 0.3504 | 0.3353 | 0.6769 | 0.3739 | 0.3959 | 0.4957 | 0.4509 | 0.2791 | 0.535 | 0.4345 | 0.4734 | **0.1784** | 0.2154 | 0.2622 |
| | 3 | 0.5015 | 0.5024 | 0.4304 | 0.506 | 0.4655 | 0.4466 | 0.5737 | 0.5118 | 0.5068 | 0.6181 | 0.501 | 0.6353 | **0.2084** | 0.2171 | 0.3668 |
| | 4 | 0.6174 | 0.6194 | 0.4986 | 0.6332 | 0.5686 | 0.5629 | 0.5509 | 0.6164 | 0.705 | 0.5827 | 0.5771 | 0.7075 | 0.2879 | **0.2056** | 0.4443 |
| Poland | 1 | 0.1278 | 0.1363 | 0.2909 | 0.1962 | 0.125 | 0.2231 | 0.1724 | 0.1832 | 0.1302 | 0.1882 | 0.145 | 0.1915 | 0.1175 | **0.1124** | 0.1186 |
| | 2 | 0.2134 | 0.214 | 0.3501 | 0.4146 | 0.1843 | 0.3383 | 0.2398 | 0.2394 | 0.22 | 0.1997 | 0.192 | 0.294 | 0.1545 | **0.1127** | 0.1972 |
| | 3 | 0.2368 | 0.2425 | 0.3921 | 0.2567 | 0.1893 | 0.3792 | 0.2199 | 0.2218 | 0.3209 | 0.2281 | 0.233 | 0.4586 | 0.218 | **0.1154** | 0.2064 |
| | 4 | 0.2533 | 0.2613 | 0.4541 | 0.2498 | 0.198 | 0.4104 | 0.2394 | 0.2123 | 0.4313 | 0.2842 | 0.2549 | 0.6935 | 0.2855 | **0.1214** | 0.2295 |
| Romania | 1 | 0.264 | 0.2522 | 0.3333 | 0.4123 | 0.2621 | 0.2643 | 0.6749 | 0.4714 | 0.3357 | 0.6227 | 0.3416 | **0.2423** | 0.2999 | 0.3236 | 0.2581 |
| | 2 | 0.5078 | 0.4616 | 0.4034 | 0.9445 | 0.4594 | 0.419 | 0.7706 | 0.5824 | 0.4201 | 0.6767 | 0.4222 | 0.4011 | 0.5078 | **0.3257** | 0.3661 |
| | 3 | 0.6976 | 0.6539 | 0.4945 | 0.8377 | 0.6457 | 0.6339 | 0.8266 | 0.5681 | 0.5183 | 0.82 | 0.5679 | 0.599 | 0.7249 | **0.3797** | 0.5514 |
| | 4 | 0.8715 | 0.8621 | 0.5969 | 0.8531 | 0.7566 | 0.7523 | 0.8197 | 0.6118 | 0.5976 | 0.9779 | 0.6842 | 0.7105 | 0.8576 | **0.4632** | 0.6436 |

Table S3: One-table summary across 9 countries, 4 horizons, and metric wMAPE.

#### S7. Forecasting trajectories across all horizons

To visually diagnose model behaviour across the full operational spectrum, Figures S11 to S46 display the epidemic trajectories (Steps 1 through 4) for all nine countries. These plots contrast the point forecasts and 80% prediction intervals of the models.

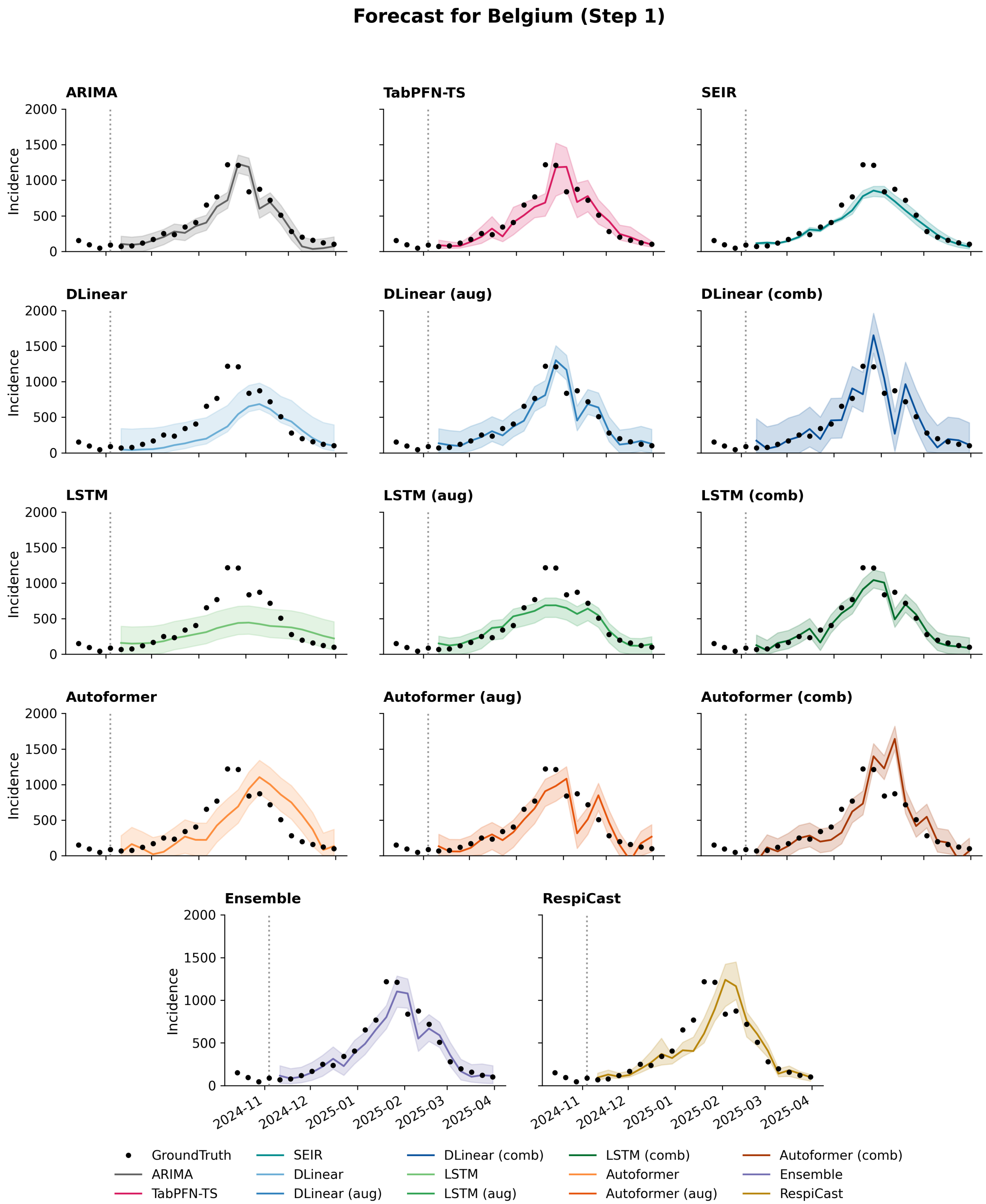


Figure S11: Model forecasts for Belgium incidence at forecasting horizon 1. Black dots denote the observed ground truth, coloured lines represent point forecasts, and shaded regions indicate predictive uncertainty.

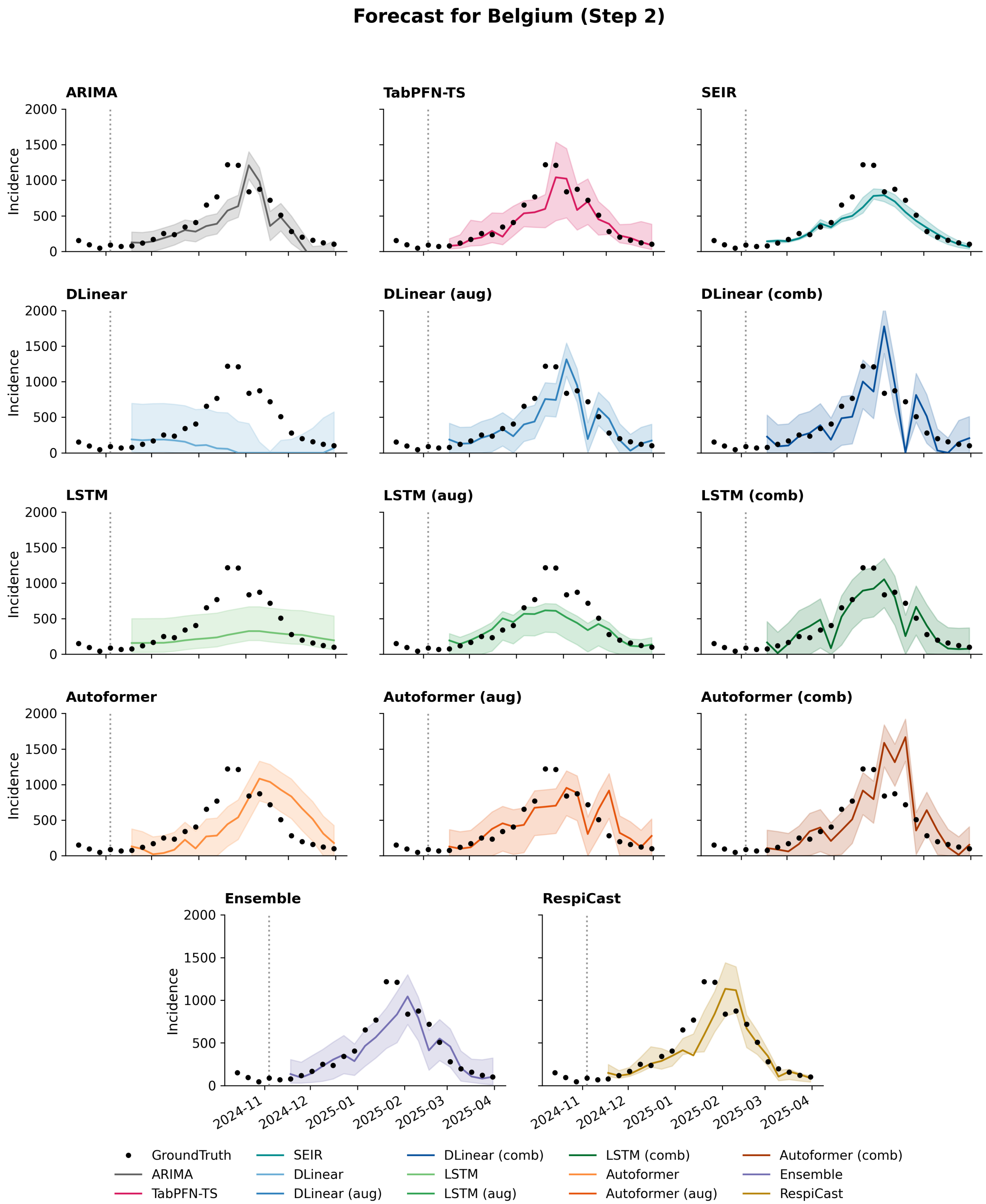


Figure S12: Model forecasts for Belgium incidence at forecasting horizon 2. Black dots denote the observed ground truth, coloured lines represent point forecasts, and shaded regions indicate predictive uncertainty.

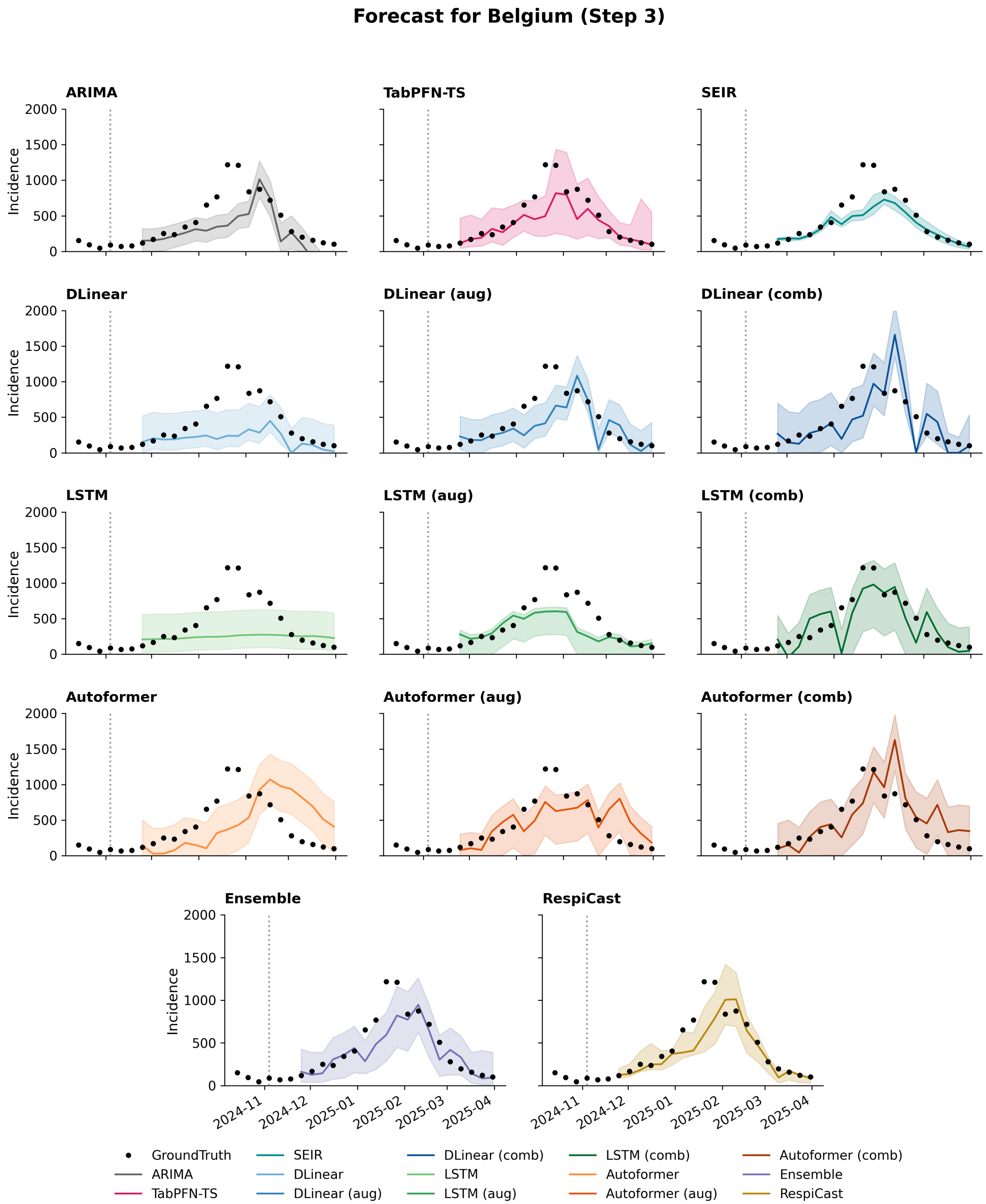


Figure S13: Model forecasts for Belgium incidence at forecasting horizon 3. Black dots denote the observed ground truth, coloured lines represent point forecasts, and shaded regions indicate predictive uncertainty.

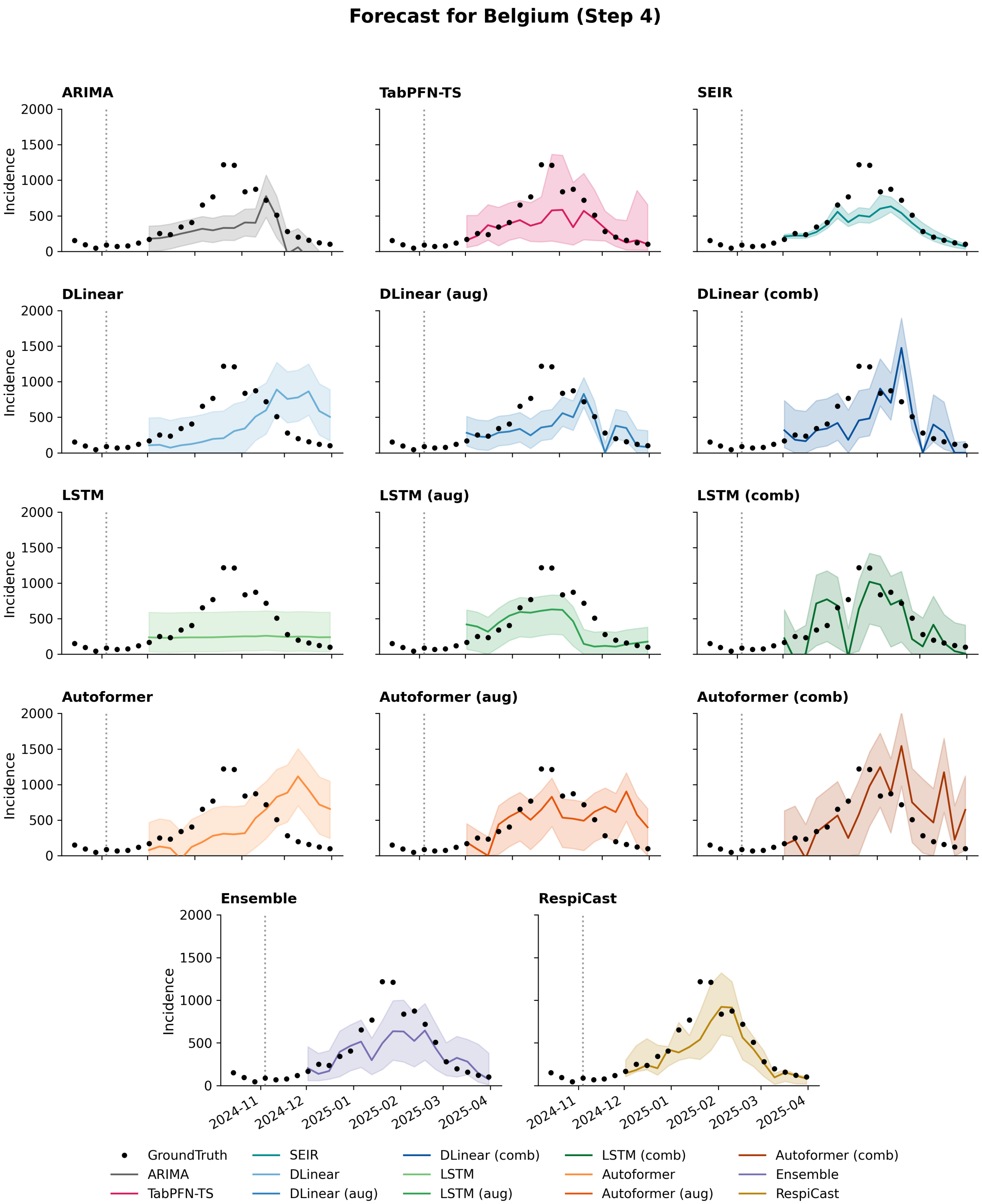


Figure S14: Model forecasts for Belgium incidence at forecasting horizon 4. Black dots denote the observed ground truth, coloured lines represent point forecasts, and shaded regions indicate predictive uncertainty.

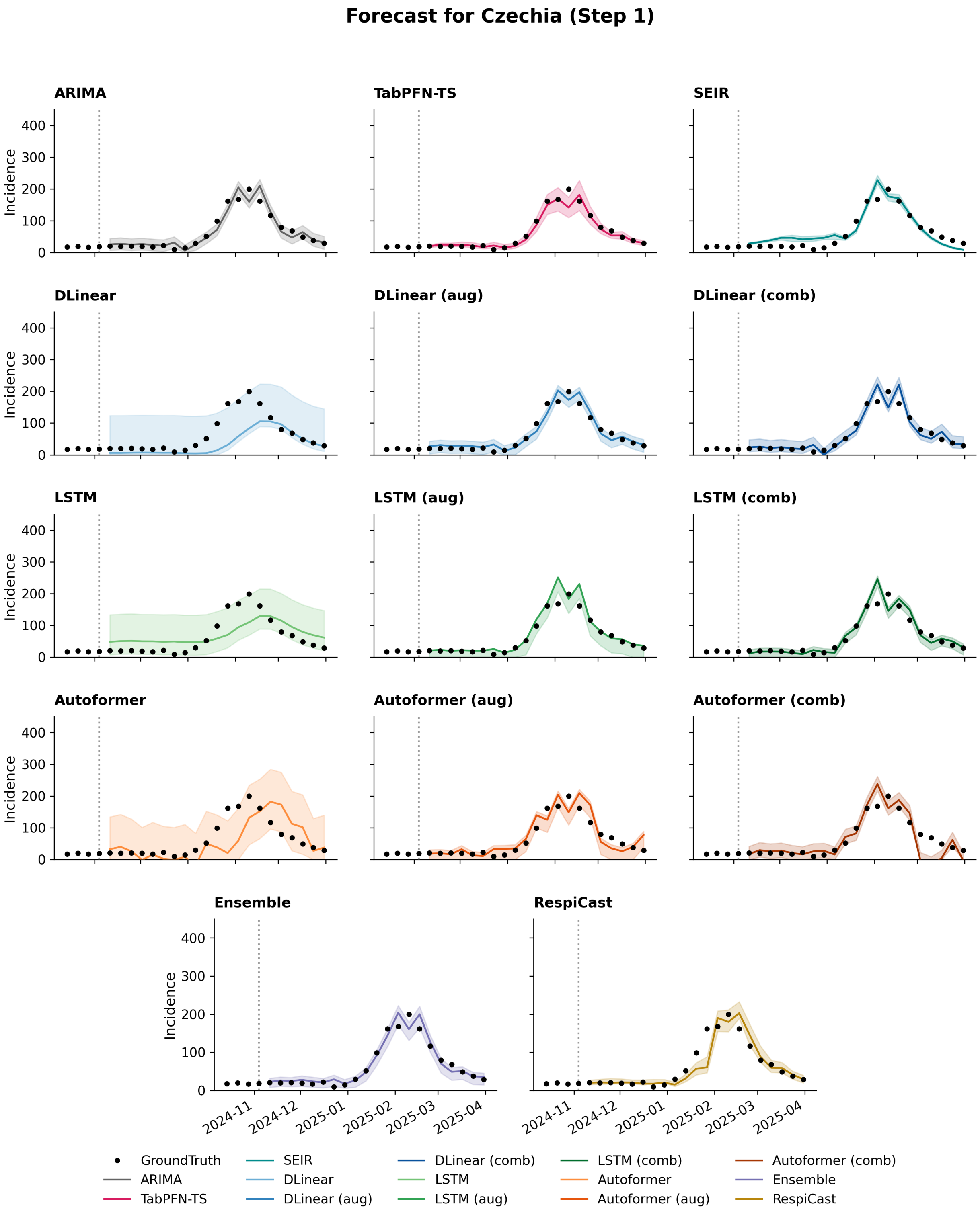


Figure S15: Model forecasts for Czechia incidence at forecasting horizon 1. Black dots denote the observed ground truth, coloured lines represent point forecasts, and shaded regions indicate predictive uncertainty.

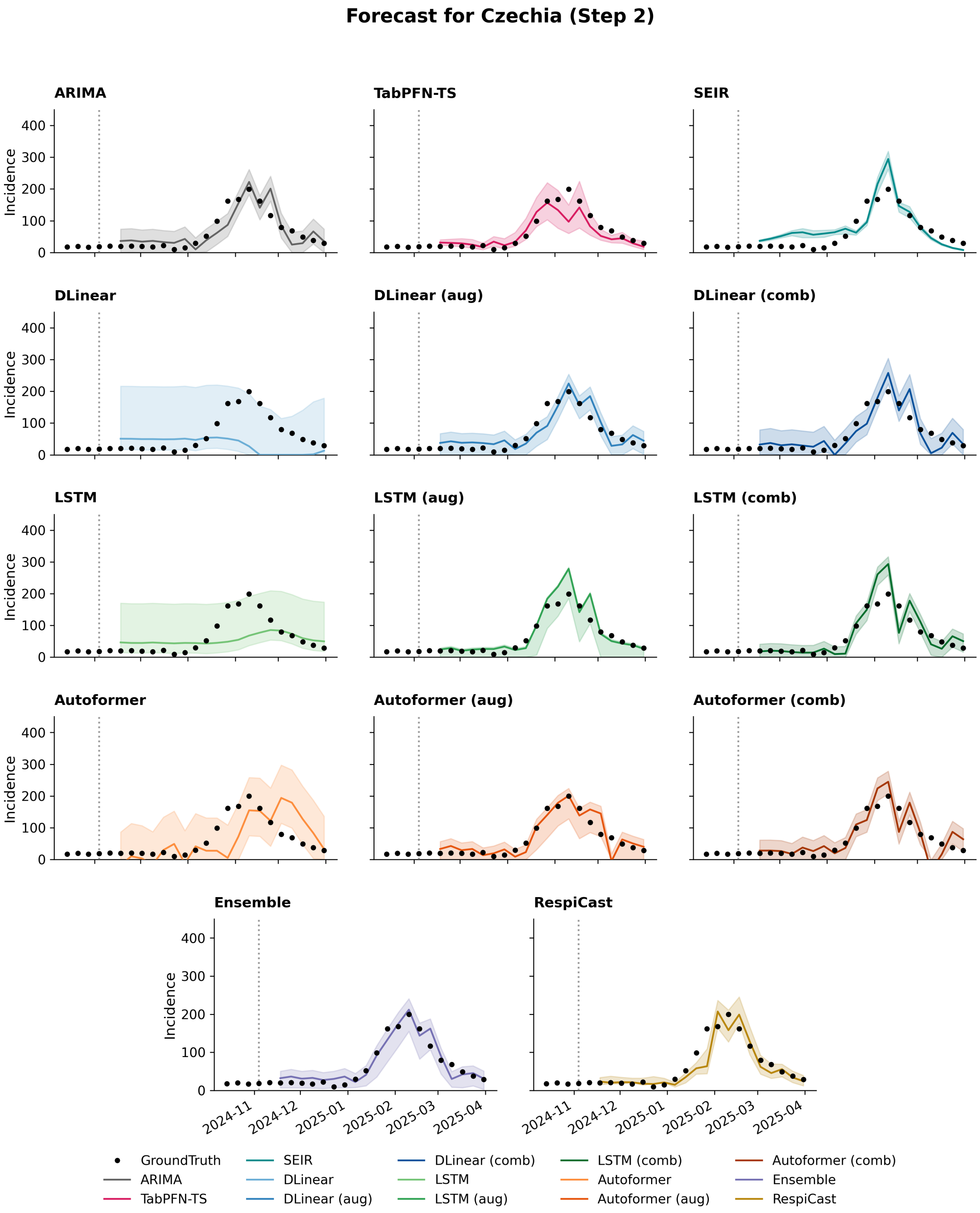


Figure S16: Model forecasts for Czechia incidence at forecasting horizon 2. Black dots denote the observed ground truth, coloured lines represent point forecasts, and shaded regions indicate predictive uncertainty.

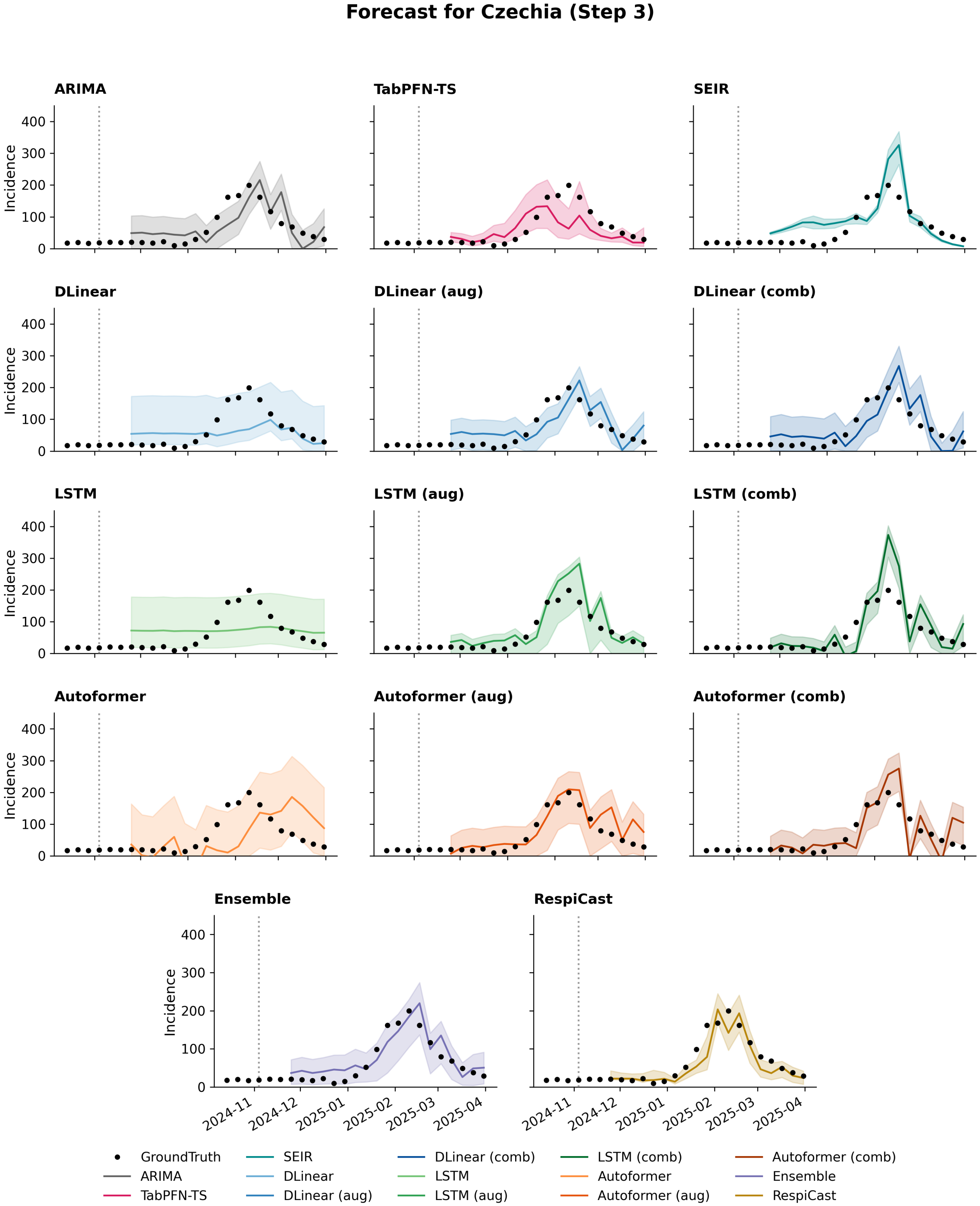


Figure S17: Model forecasts for Czechia incidence at forecasting horizon 3. Black dots denote the observed ground truth, coloured lines represent point forecasts, and shaded regions indicate predictive uncertainty.

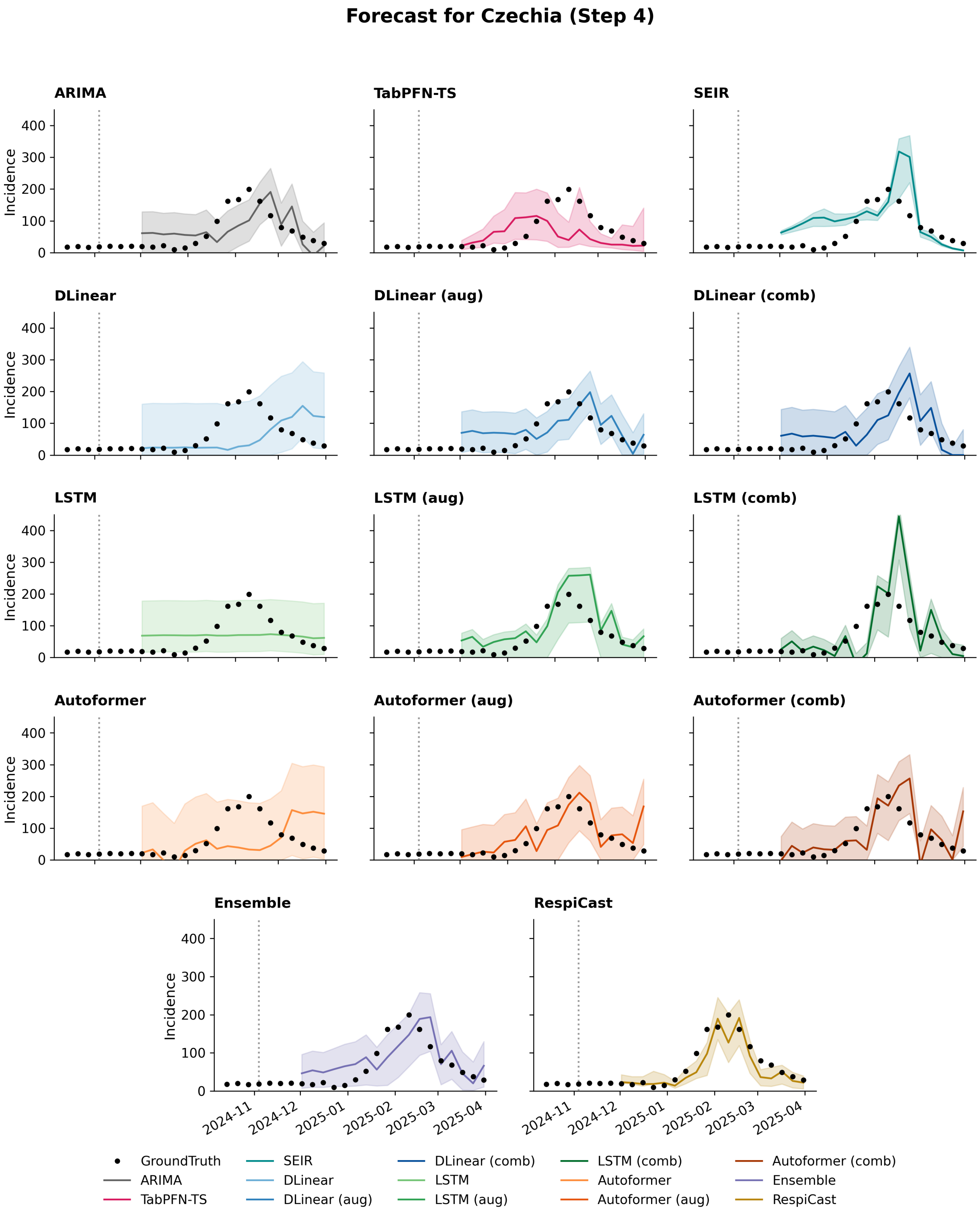


Figure S18: Model forecasts for Czechia incidence at forecasting horizon 4. Black dots denote the observed ground truth, coloured lines represent point forecasts, and shaded regions indicate predictive uncertainty.

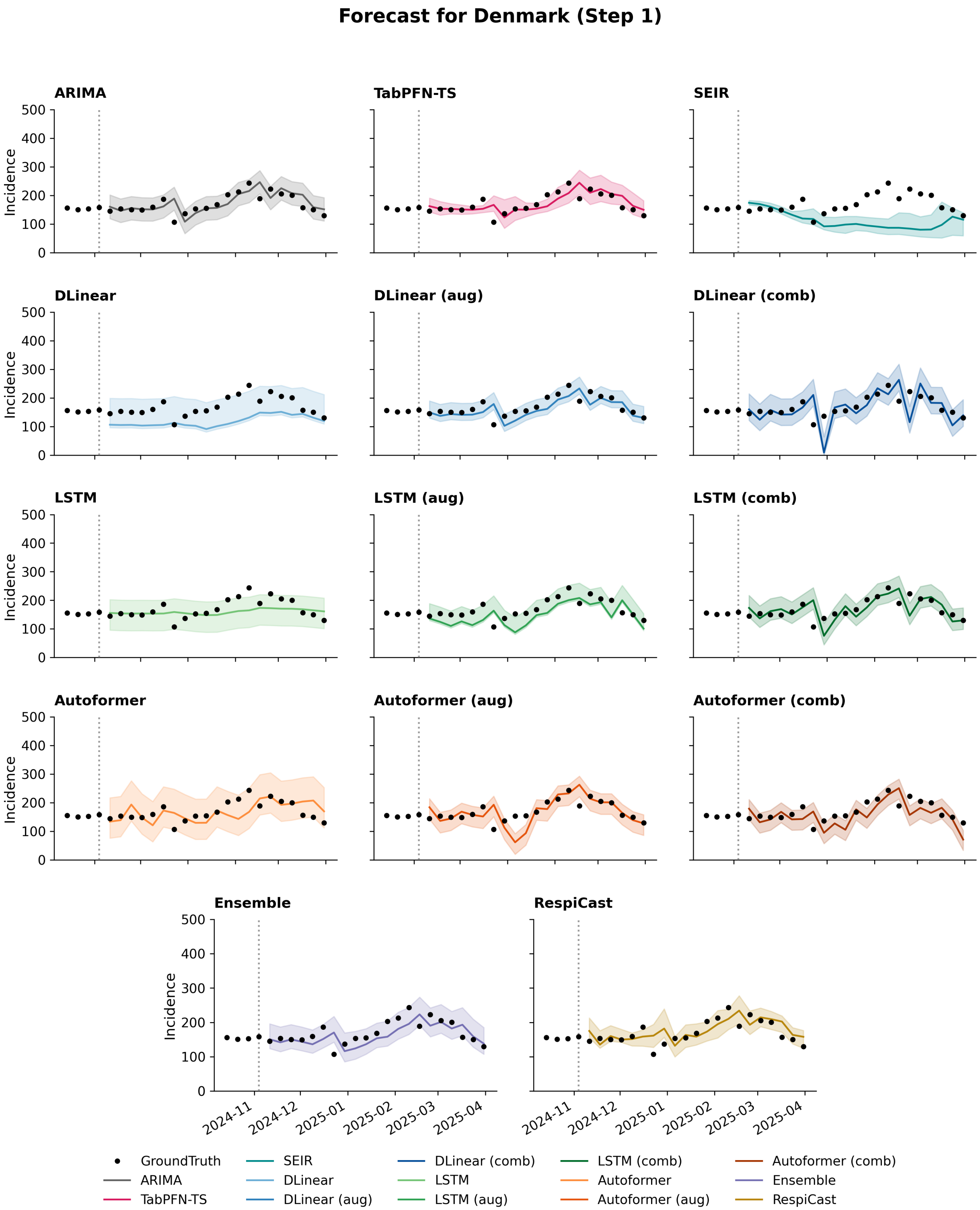


Figure S19: Model forecasts for Denmark incidence at forecasting horizon 1. Black dots denote the observed ground truth, coloured lines represent point forecasts, and shaded regions indicate predictive uncertainty.

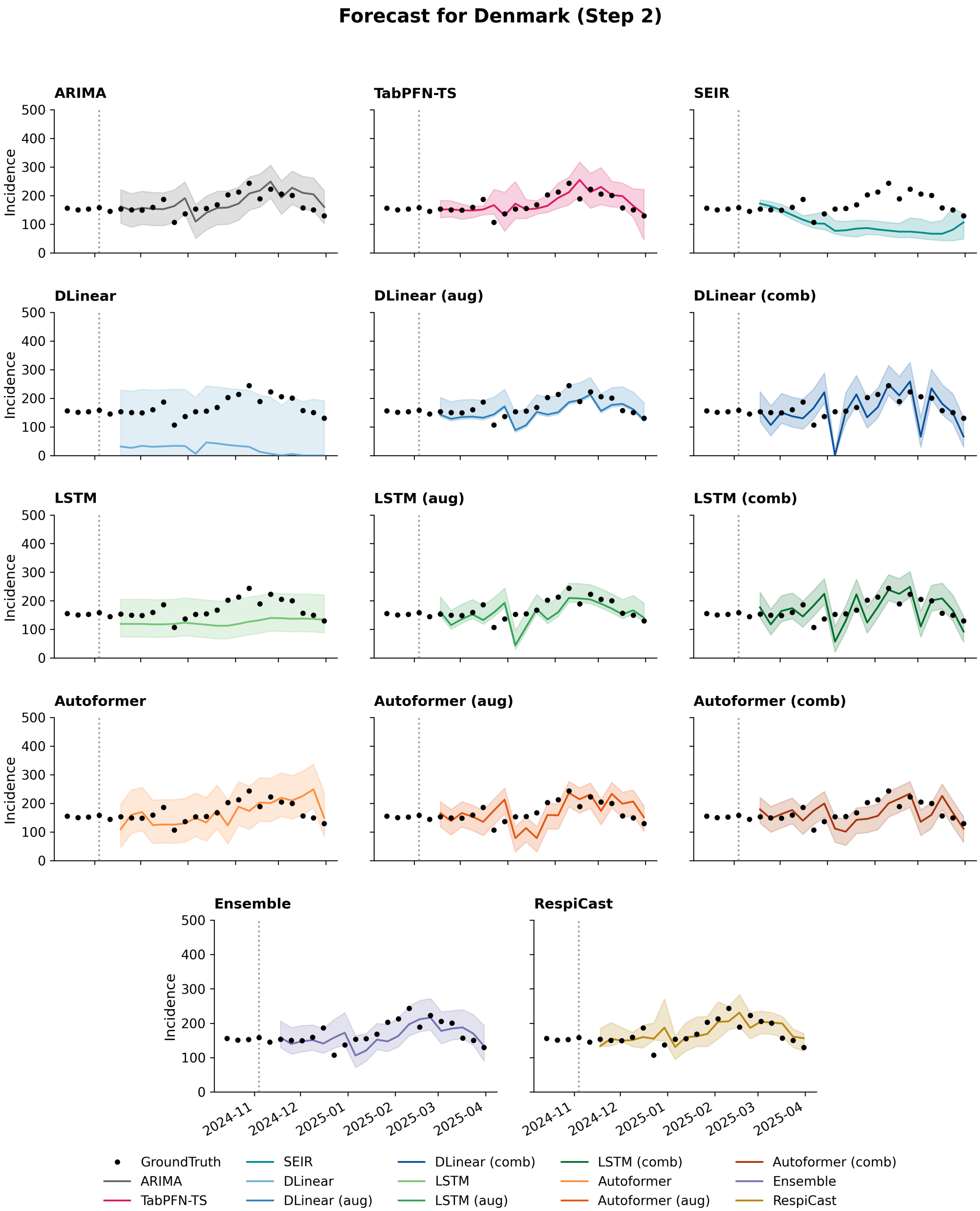


Figure S20: Model forecasts for Denmark incidence at forecasting horizon 2. Black dots denote the observed ground truth, coloured lines represent point forecasts, and shaded regions indicate predictive uncertainty.

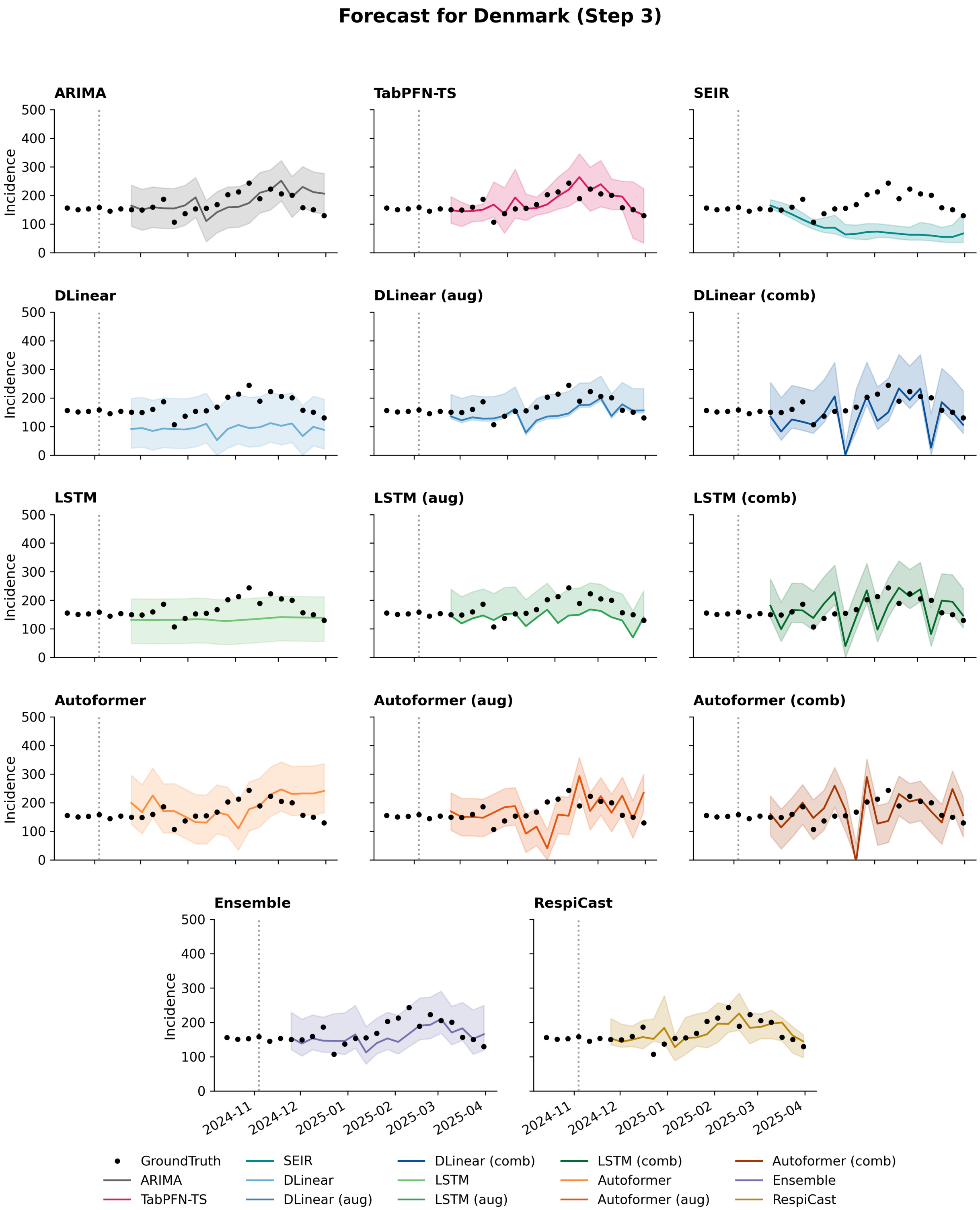


Figure S21: Model forecasts for Denmark incidence at forecasting horizon 3. Black dots denote the observed ground truth, coloured lines represent point forecasts, and shaded regions indicate predictive uncertainty.

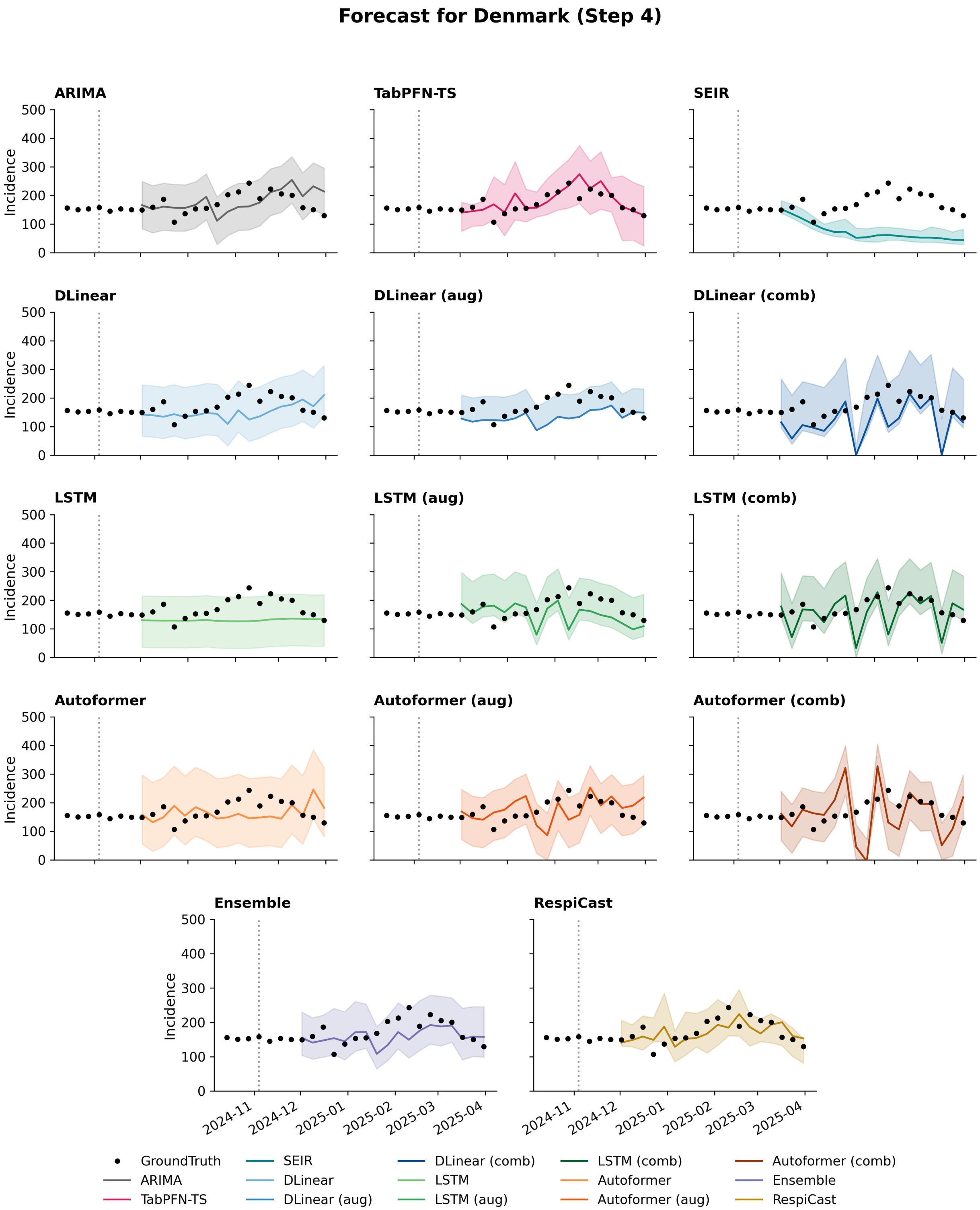


Figure S22: Model forecasts for Denmark incidence at forecasting horizon 4. Black dots denote the observed ground truth, coloured lines represent point forecasts, and shaded regions indicate predictive uncertainty.

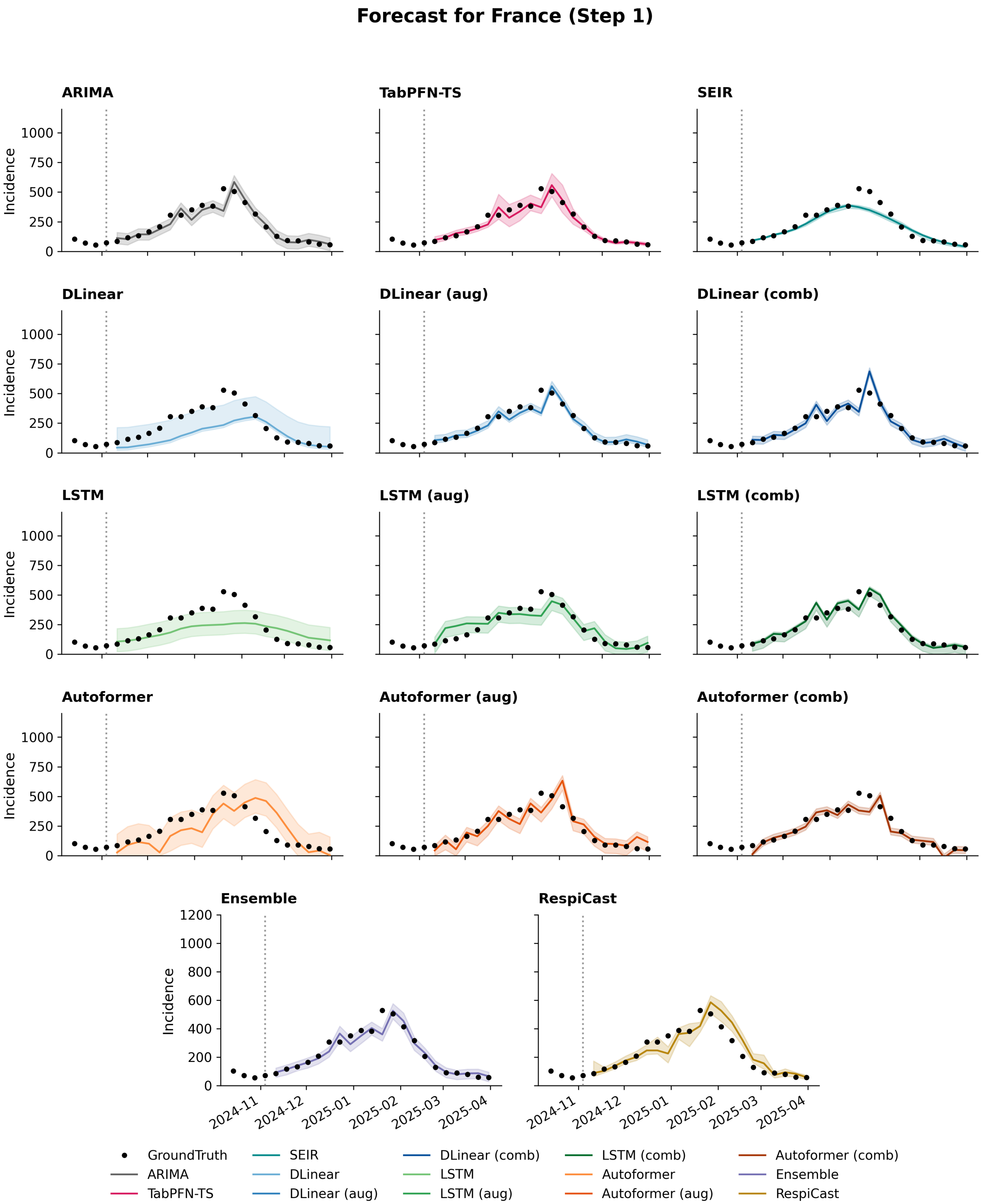


Figure S23: Model forecasts for France incidence at forecasting horizon 1. Black dots denote the observed ground truth, coloured lines represent point forecasts, and shaded regions indicate predictive uncertainty.

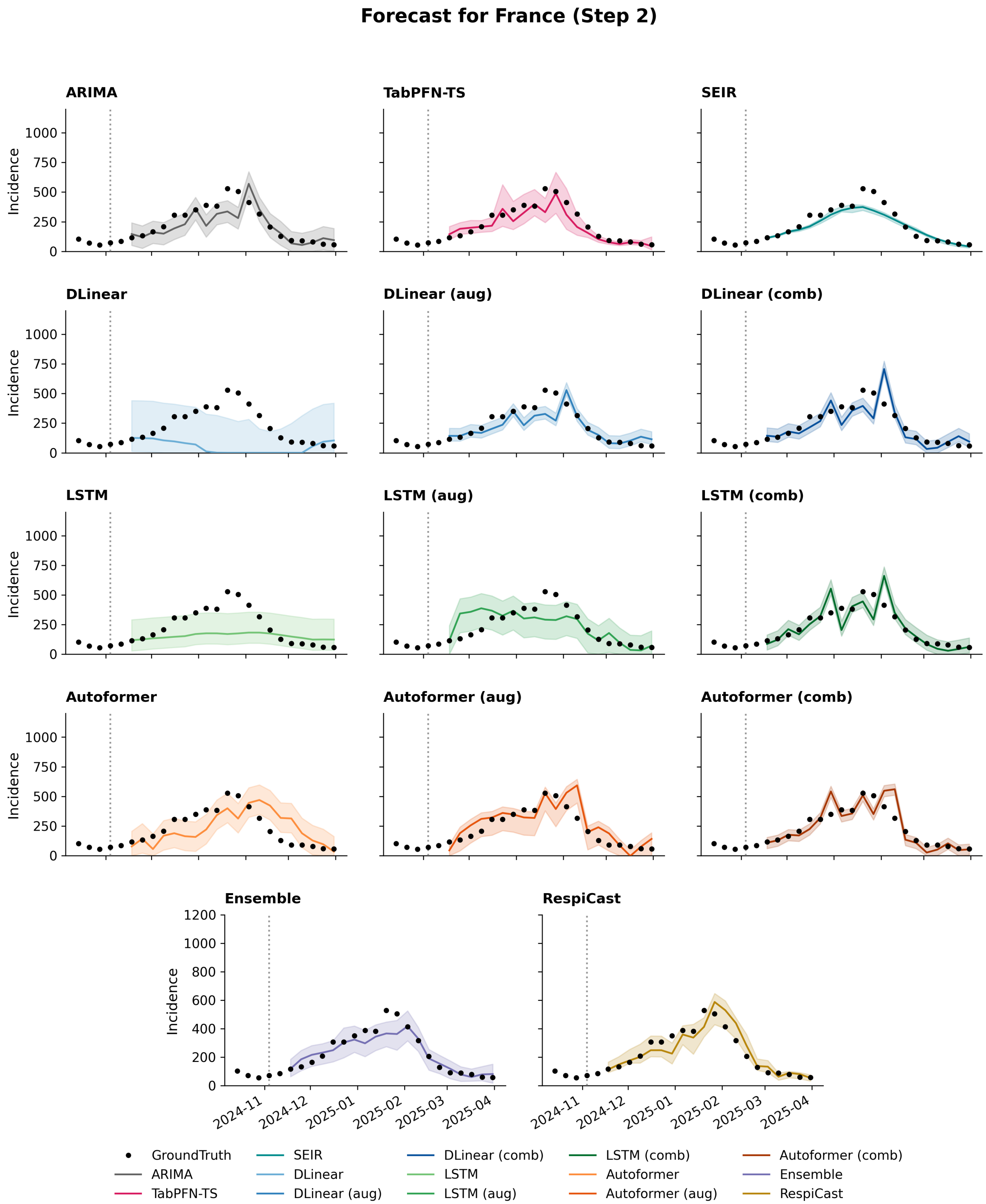


Figure S24: Model forecasts for France incidence at forecasting horizon 2. Black dots denote the observed ground truth, coloured lines represent point forecasts, and shaded regions indicate predictive uncertainty.

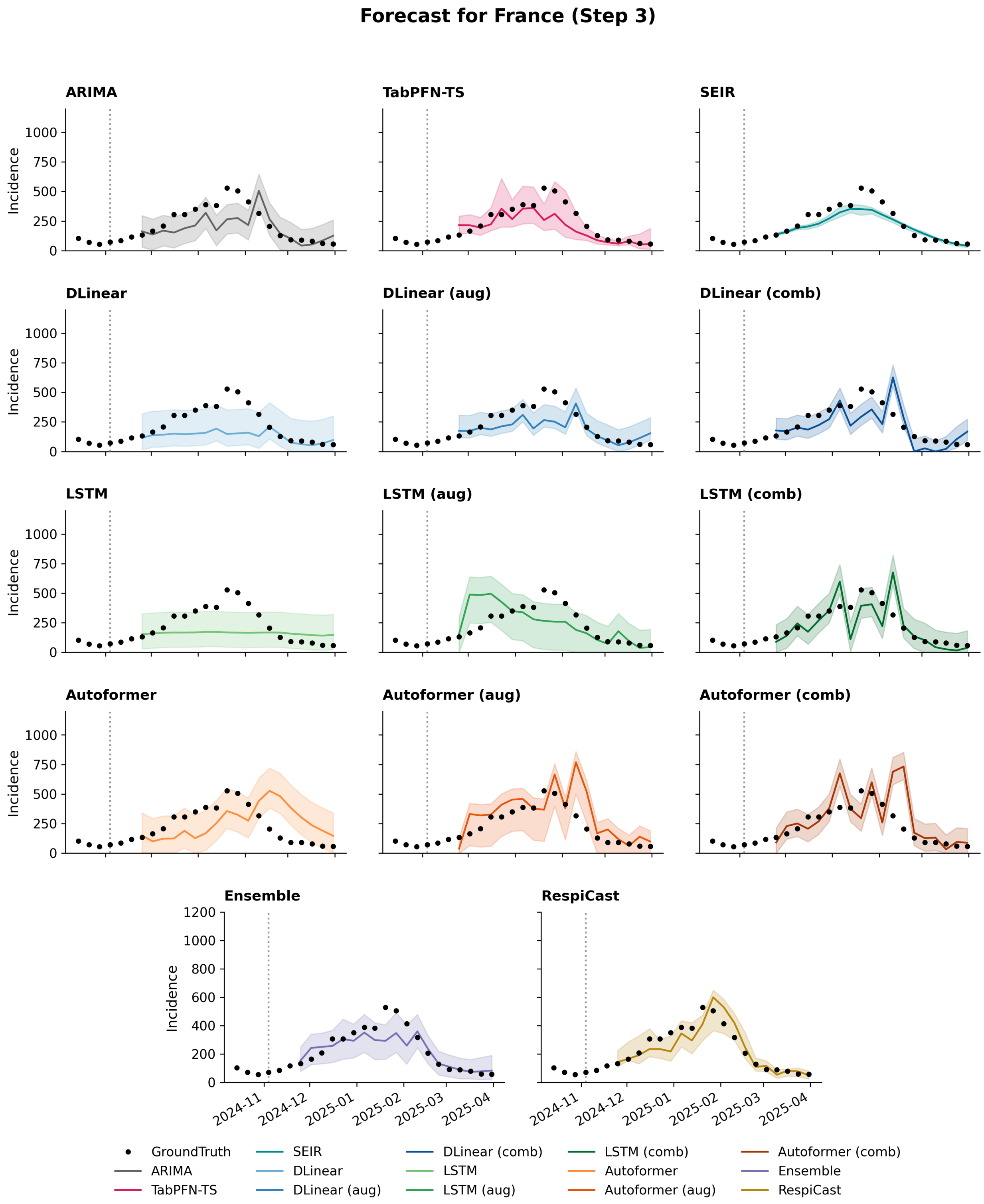


Figure S25: Model forecasts for France incidence at forecasting horizon 3. Black dots denote the observed ground truth, coloured lines represent point forecasts, and shaded regions indicate predictive uncertainty.

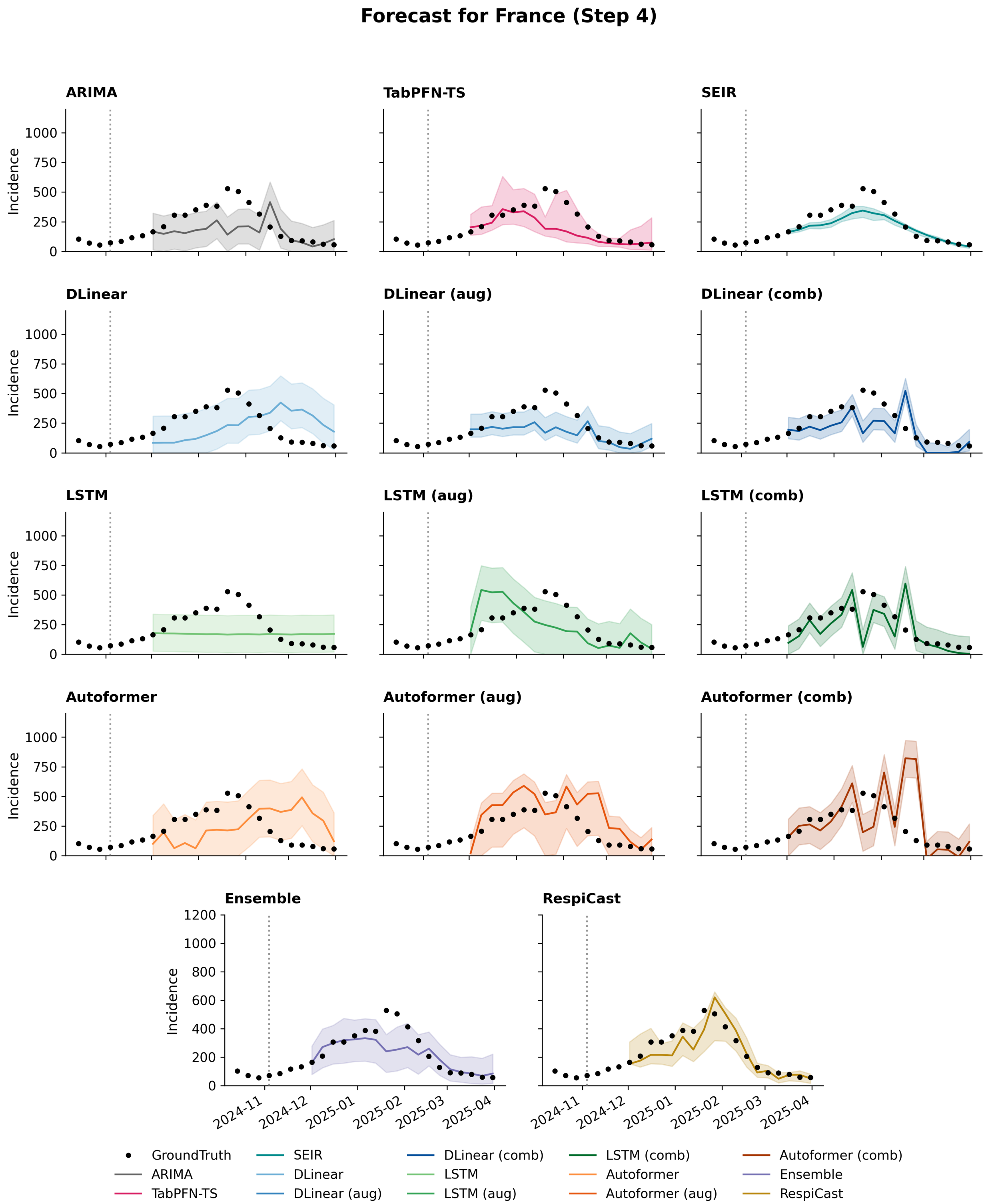


Figure S26: Model forecasts for France incidence at forecasting horizon 4. Black dots denote the observed ground truth, coloured lines represent point forecasts, and shaded regions indicate predictive uncertainty.

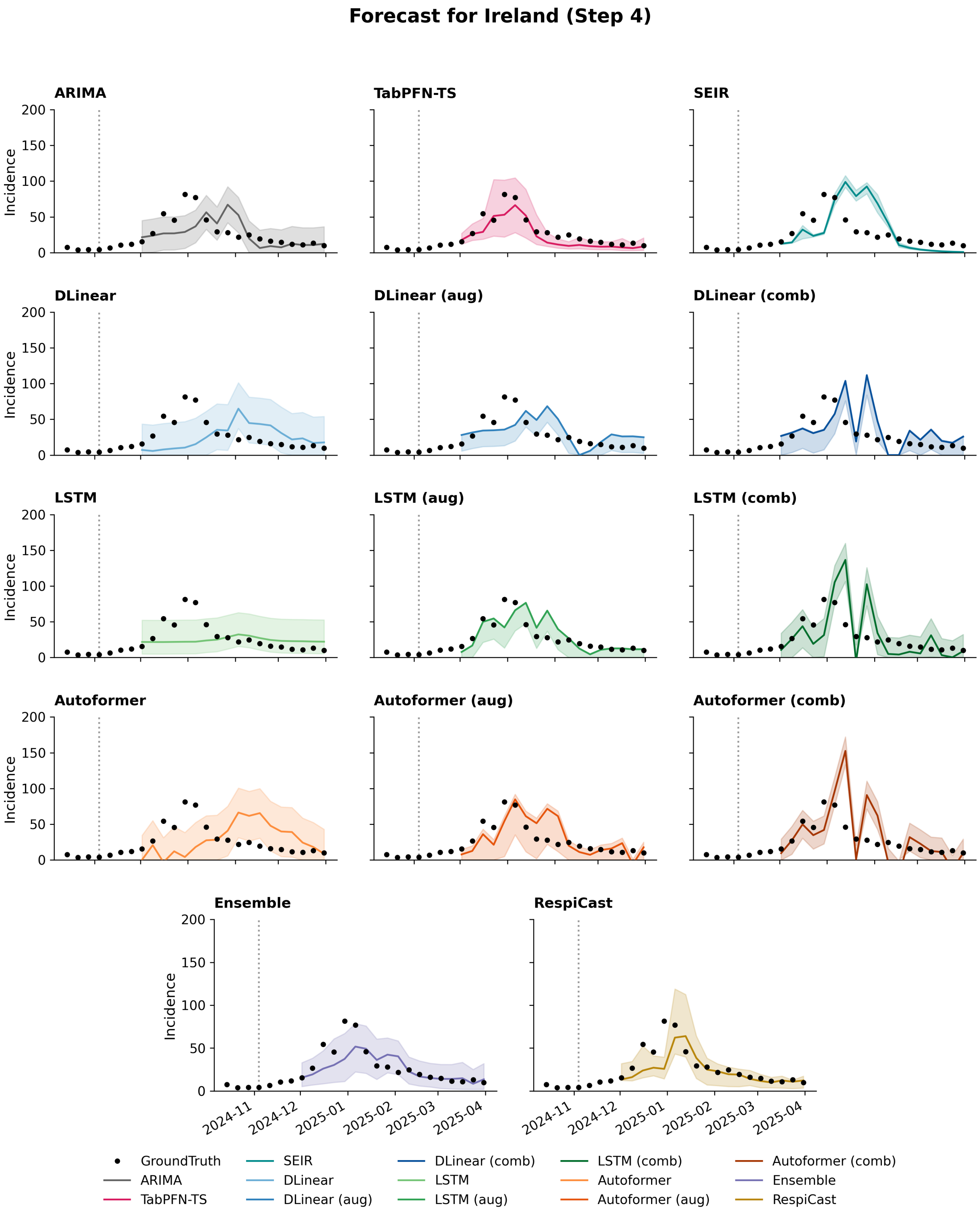


Figure S27: Model forecasts for Ireland incidence at forecasting horizon 1. Black dots denote the observed ground truth, coloured lines represent point forecasts, and shaded regions indicate predictive uncertainty.

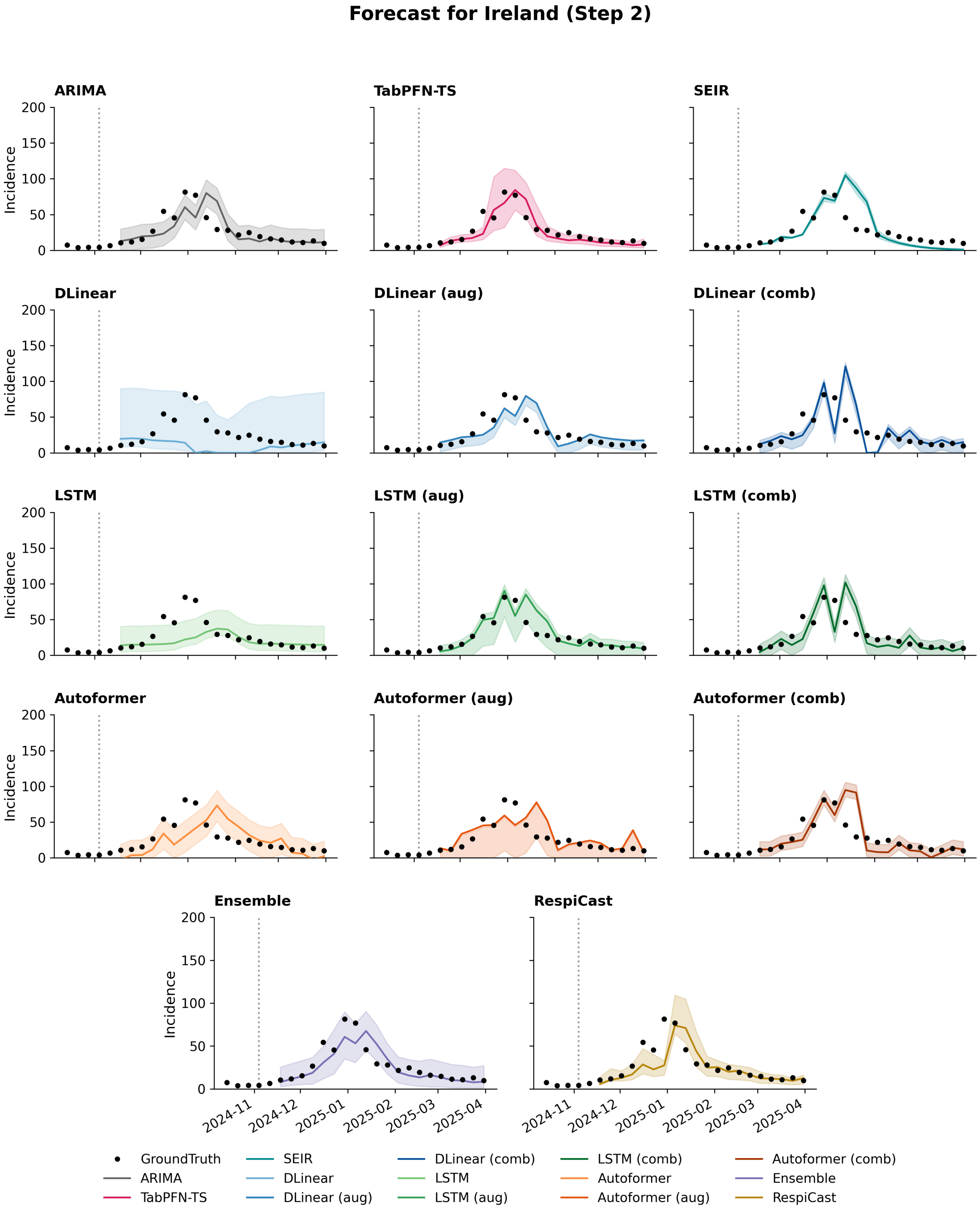


Figure S28: Model forecasts for Ireland incidence at forecasting horizon 2. Black dots denote the observed ground truth, coloured lines represent point forecasts, and shaded regions indicate predictive uncertainty.

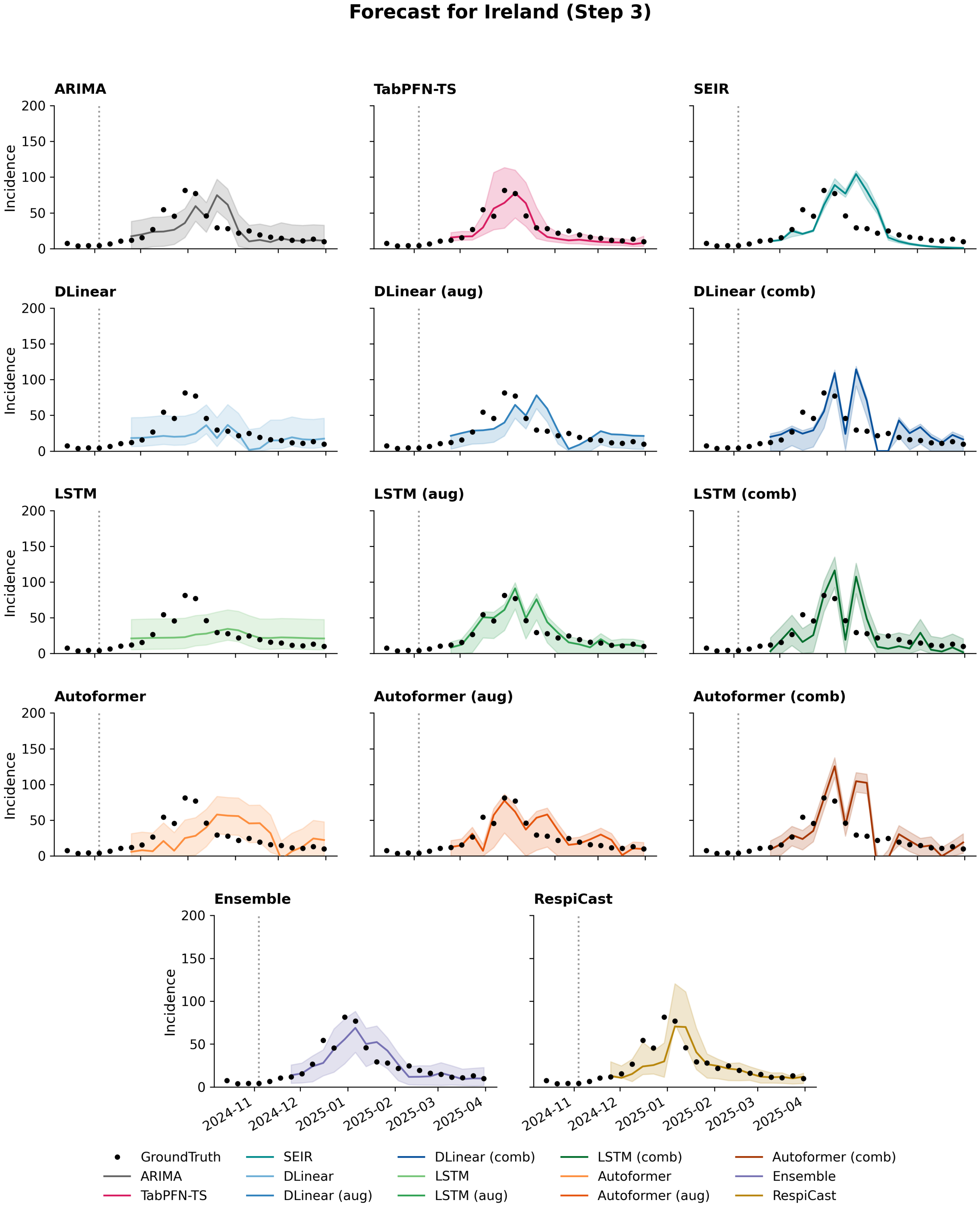


Figure S29: Model forecasts for Ireland incidence at forecasting horizon 3. Black dots denote the observed ground truth, coloured lines represent point forecasts, and shaded regions indicate predictive uncertainty.

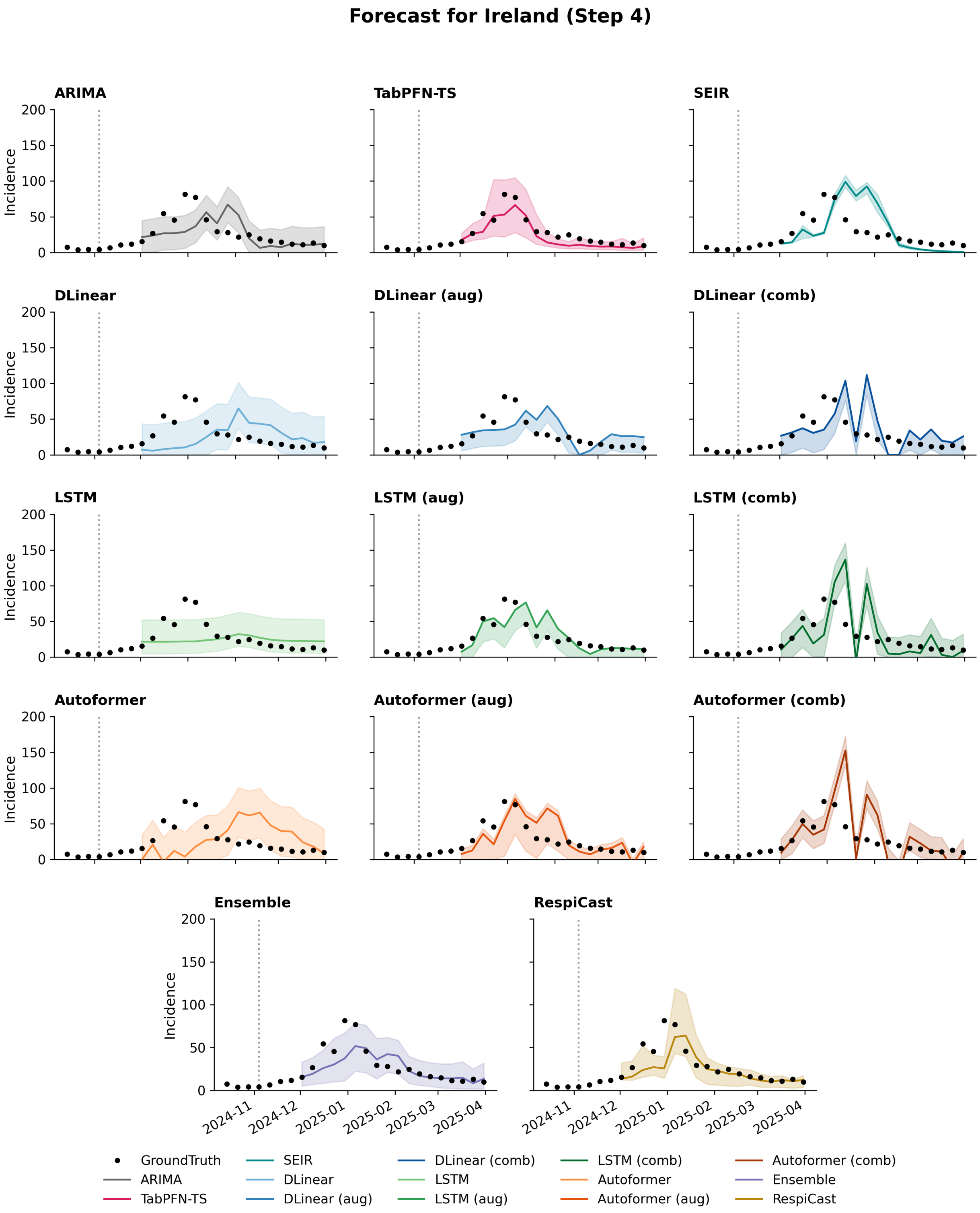


Figure S30: Model forecasts for Ireland incidence at forecasting horizon 4. Black dots denote the observed ground truth, coloured lines represent point forecasts, and shaded regions indicate predictive uncertainty.

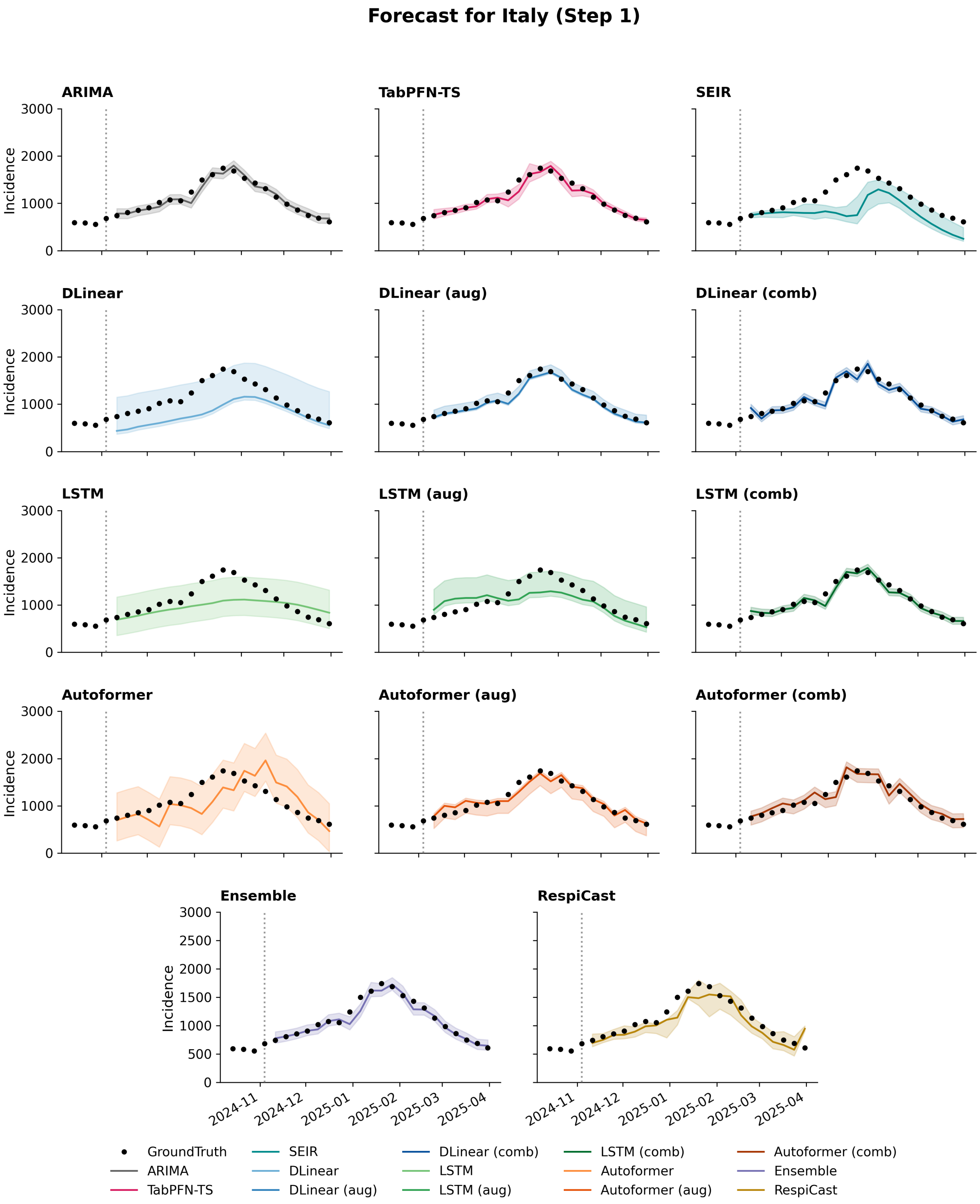


Figure S31: Model forecasts for Italy incidence at forecasting horizon 1. Black dots denote the observed ground truth, coloured lines represent point forecasts, and shaded regions indicate predictive uncertainty.

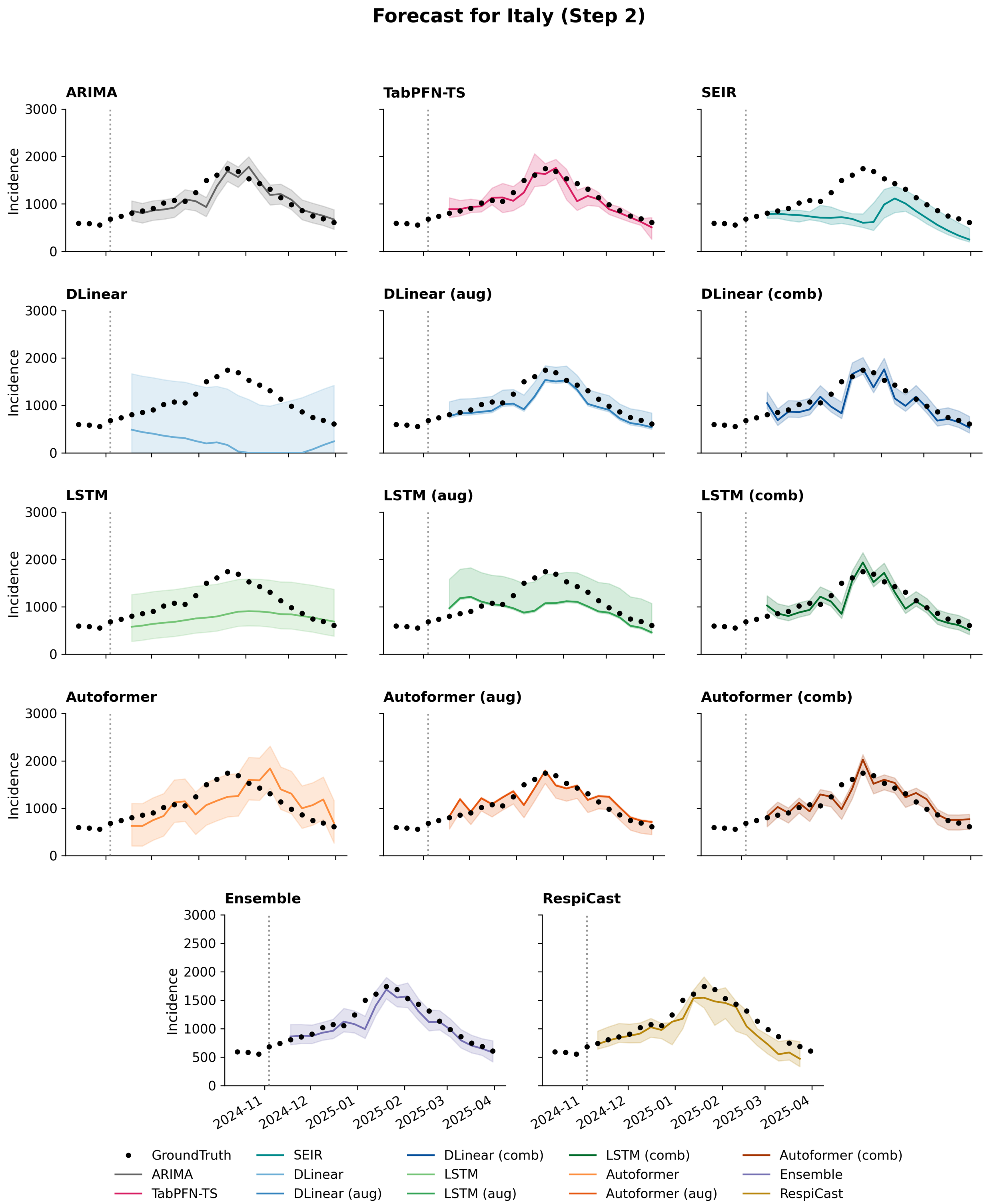


Figure S32: Model forecasts for Italy incidence at forecasting horizon 2. Black dots denote the observed ground truth, coloured lines represent point forecasts, and shaded regions indicate predictive uncertainty.

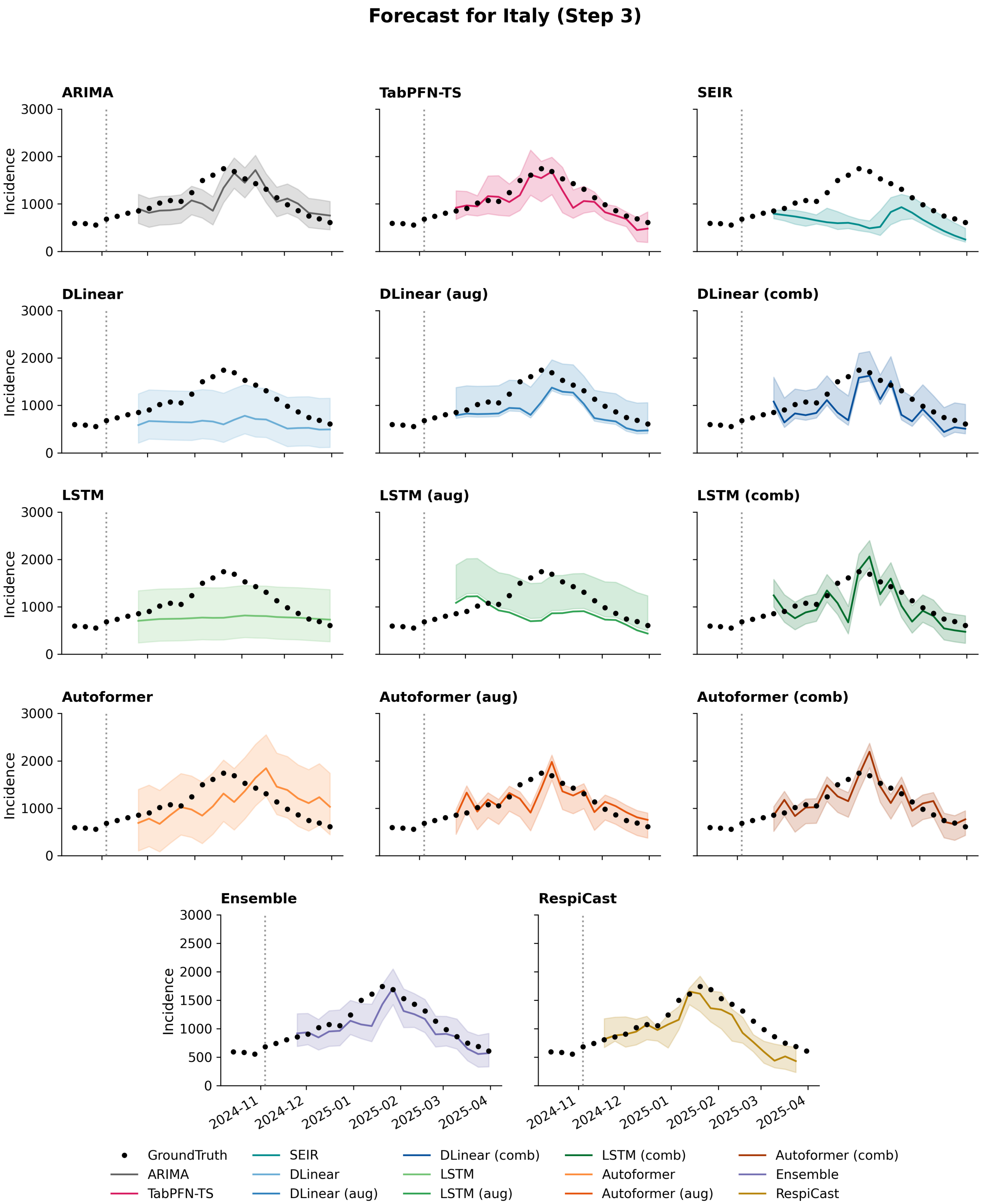


Figure S33: Model forecasts for Italy incidence at forecasting horizon 3. Black dots denote the observed ground truth, coloured lines represent point forecasts, and shaded regions indicate predictive uncertainty.

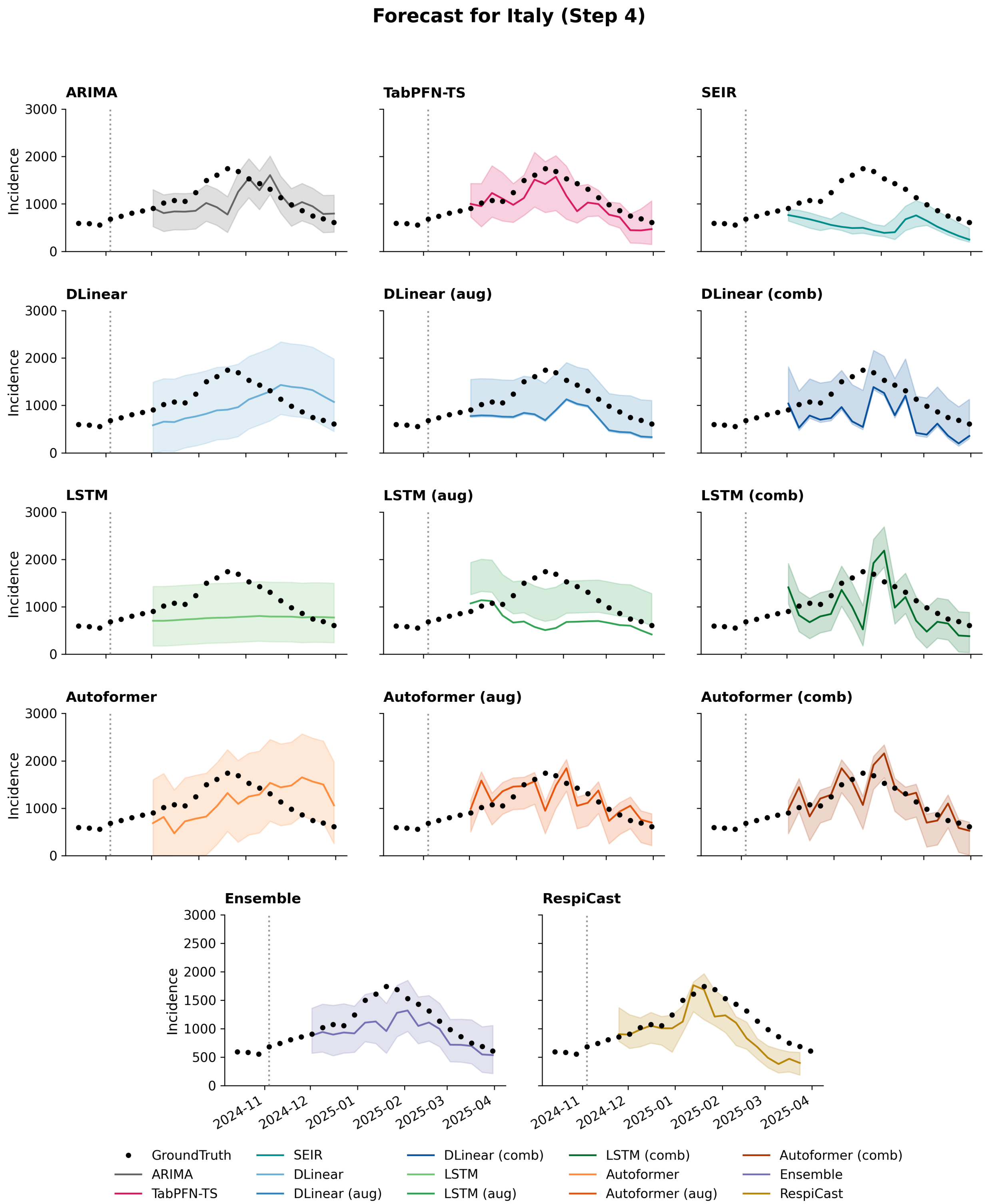


Figure S34: Model forecasts for Italy incidence at forecasting horizon 4. Black dots denote the observed ground truth, coloured lines represent point forecasts, and shaded regions indicate predictive uncertainty.

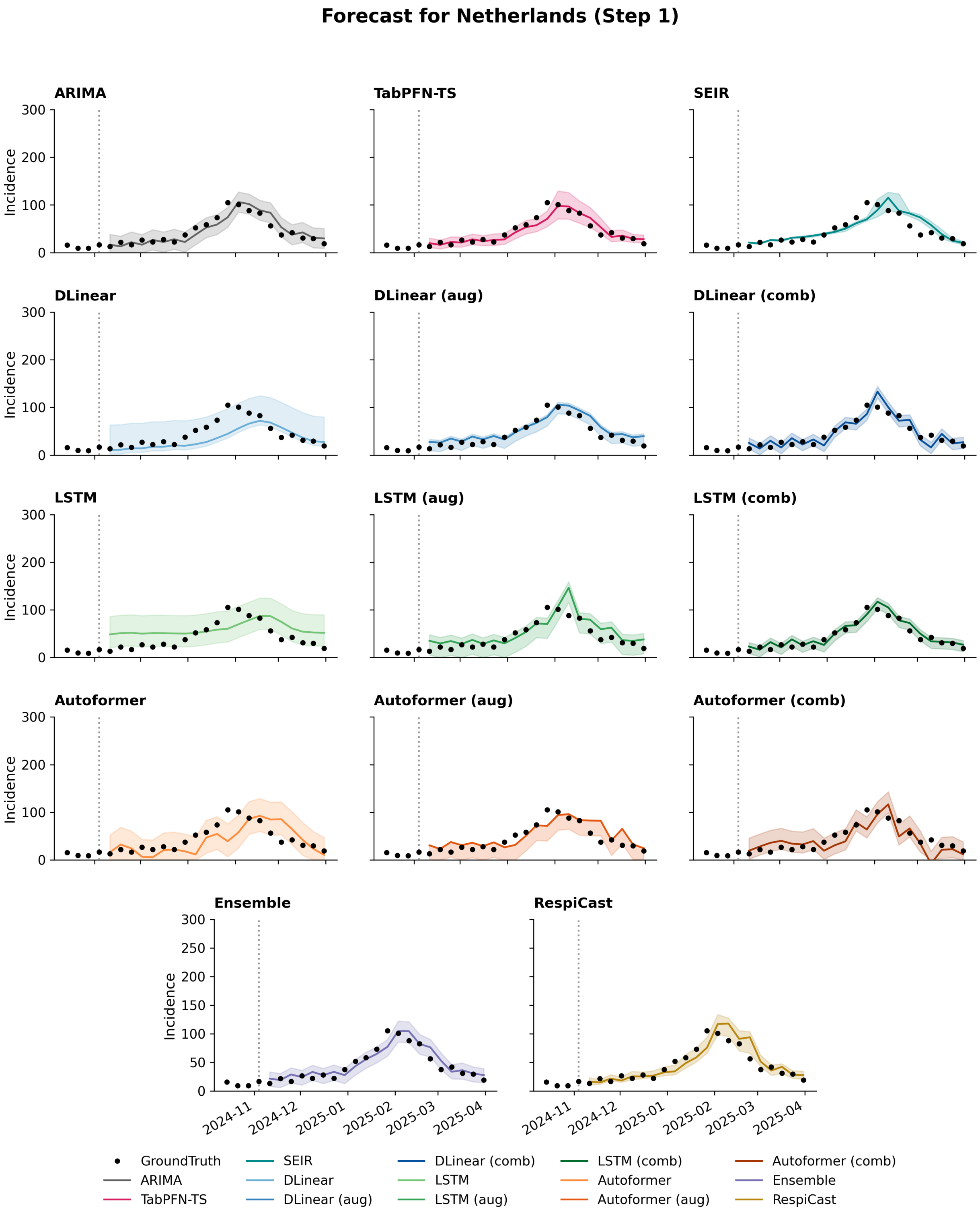


Figure S35: Model forecasts for Netherlands incidence at forecasting horizon 1. Black dots denote the observed ground truth, coloured lines represent point forecasts, and shaded regions indicate predictive uncertainty.

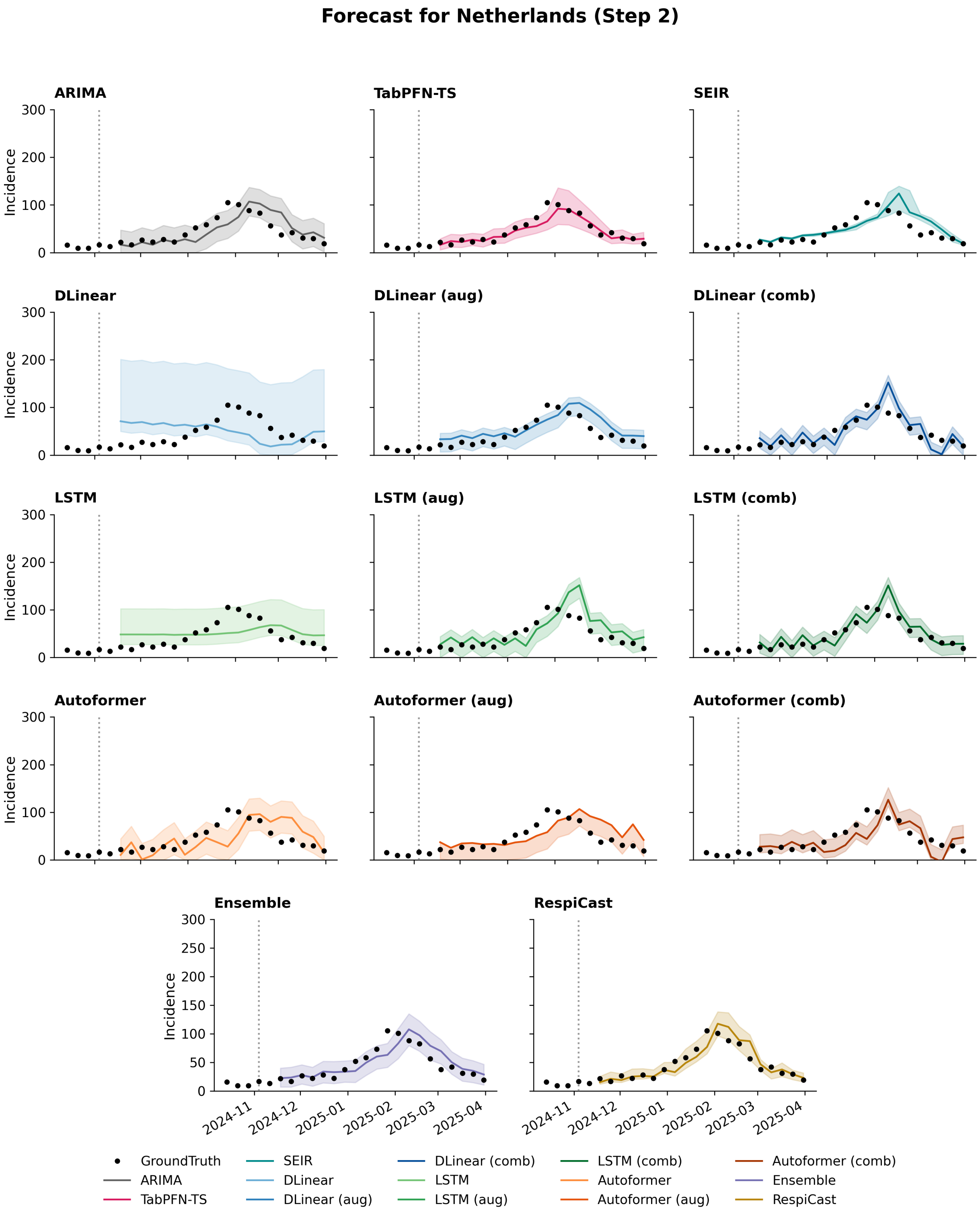


Figure S36: Model forecasts for Netherlands incidence at forecasting horizon 2. Black dots denote the observed ground truth, coloured lines represent point forecasts, and shaded regions indicate predictive uncertainty.

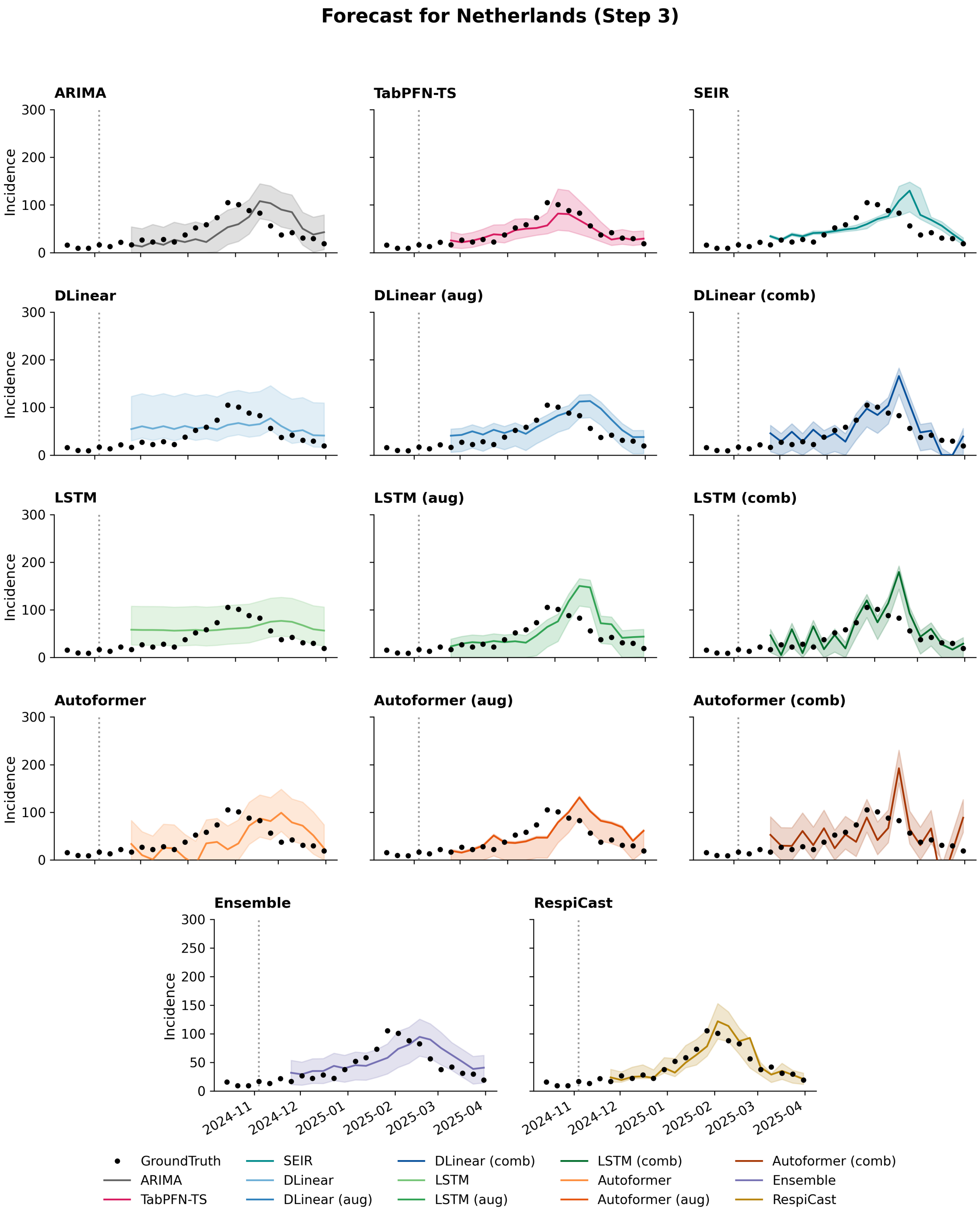


Figure S37: Model forecasts for Netherlands incidence at forecasting horizon 3. Black dots denote the observed ground truth, coloured lines represent point forecasts, and shaded regions indicate predictive uncertainty.

Forecast for Netherlands (Step 4)

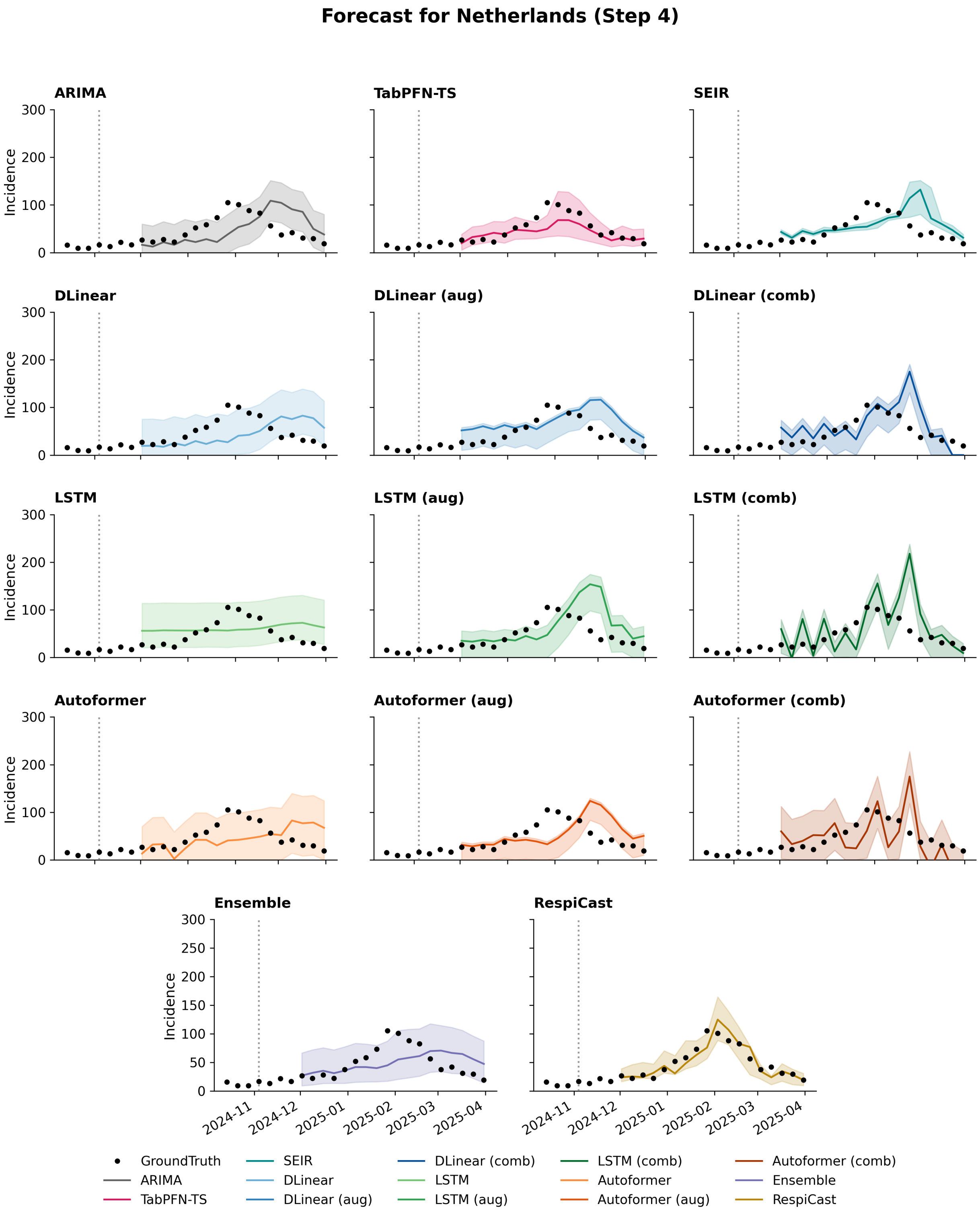


Figure S38: Model forecasts for Netherlands incidence at forecasting horizon 4. Black dots denote the observed ground truth, coloured lines represent point forecasts, and shaded regions indicate predictive uncertainty.

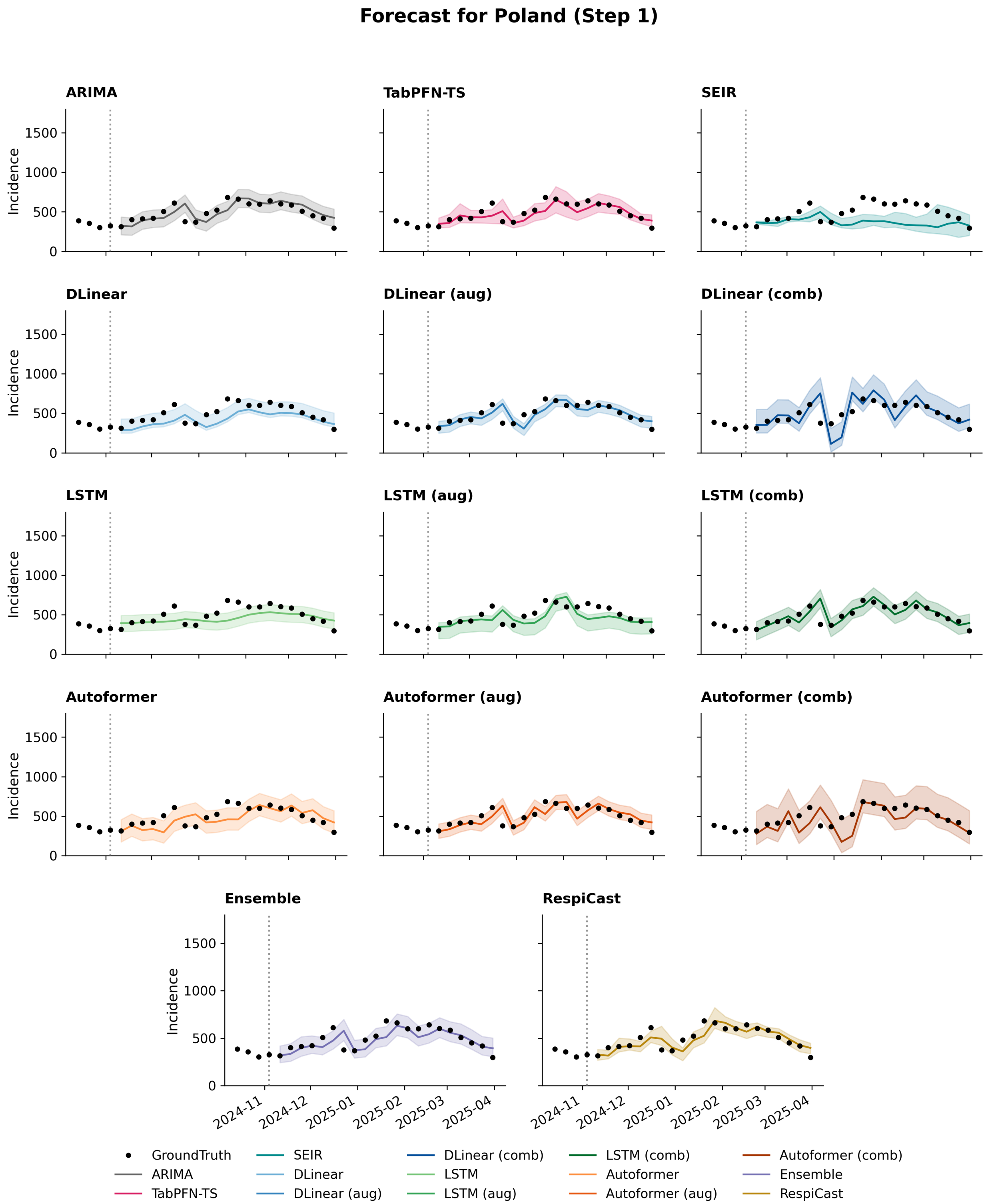


Figure S39: Model forecasts for Poland incidence at forecasting horizon 1. Black dots denote the observed ground truth, coloured lines represent point forecasts, and shaded regions indicate predictive uncertainty.

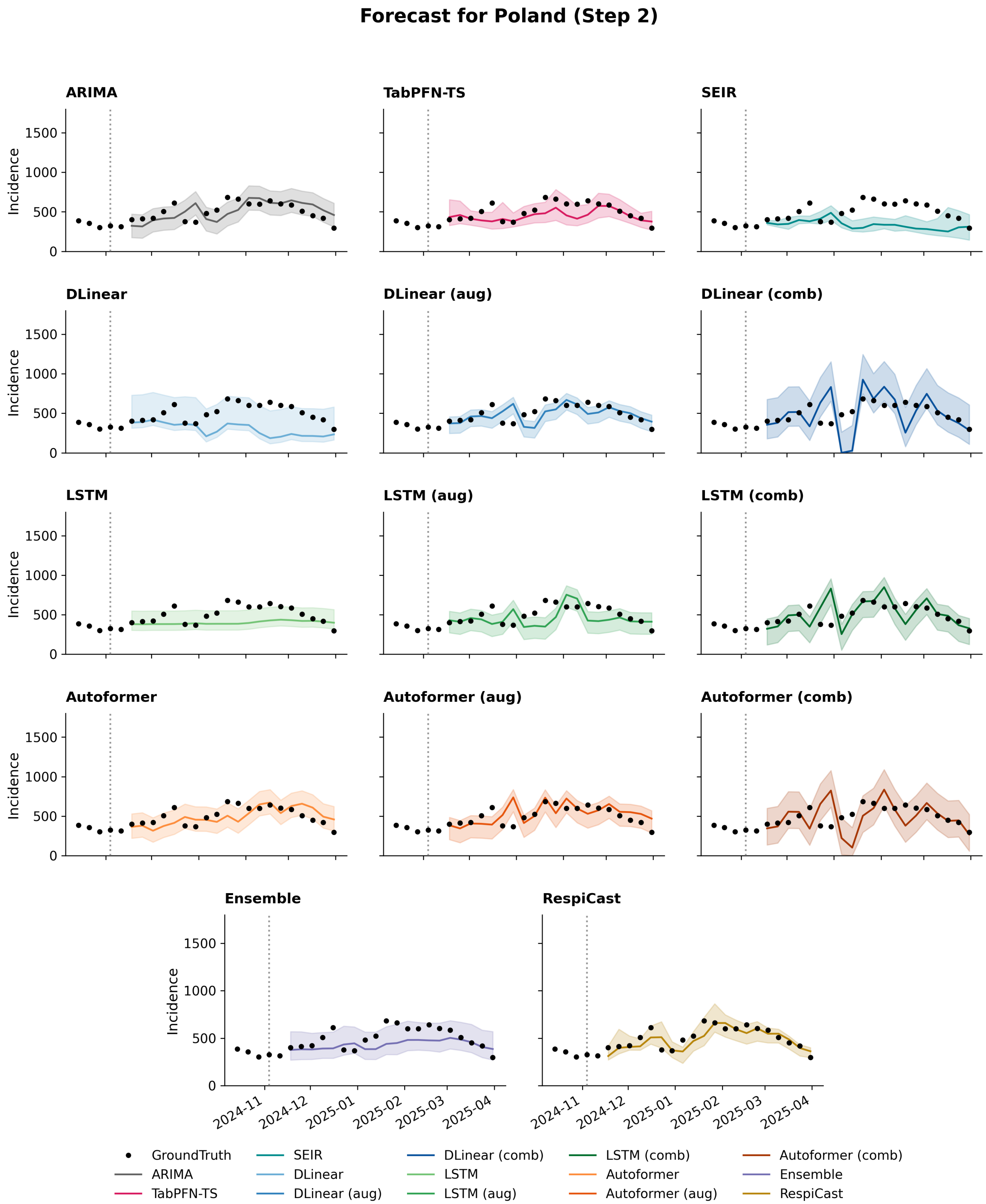


Figure S40: Model forecasts for Poland incidence at forecasting horizon 2. Black dots denote the observed ground truth, coloured lines represent point forecasts, and shaded regions indicate predictive uncertainty.

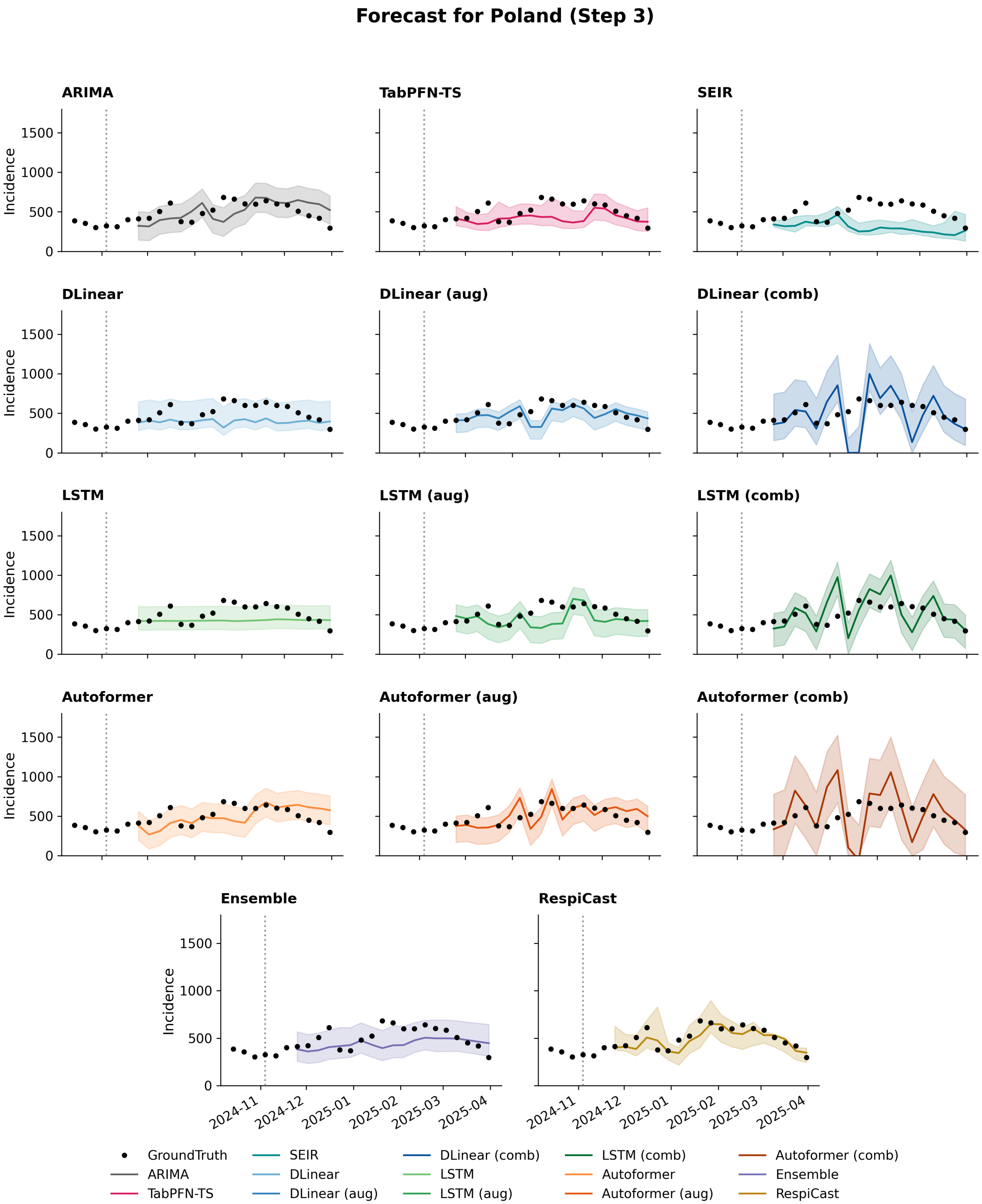


Figure S41: Model forecasts for Poland incidence at forecasting horizon 3. Black dots denote the observed ground truth, coloured lines represent point forecasts, and shaded regions indicate predictive uncertainty.

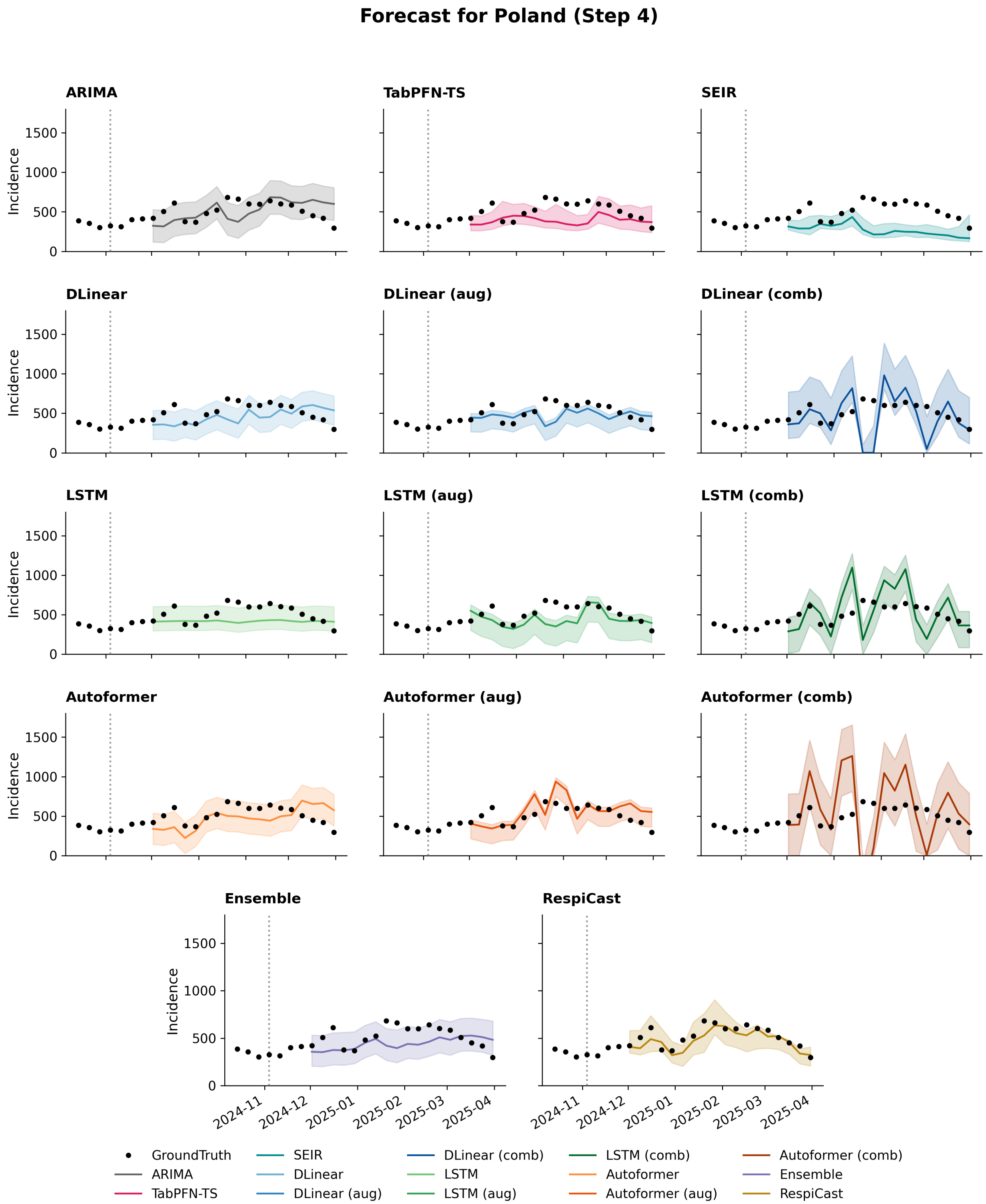


Figure S42: Model forecasts for Poland incidence at forecasting horizon 4. Black dots denote the observed ground truth, coloured lines represent point forecasts, and shaded regions indicate predictive uncertainty.

#### Forecast for Romania (Step 1)

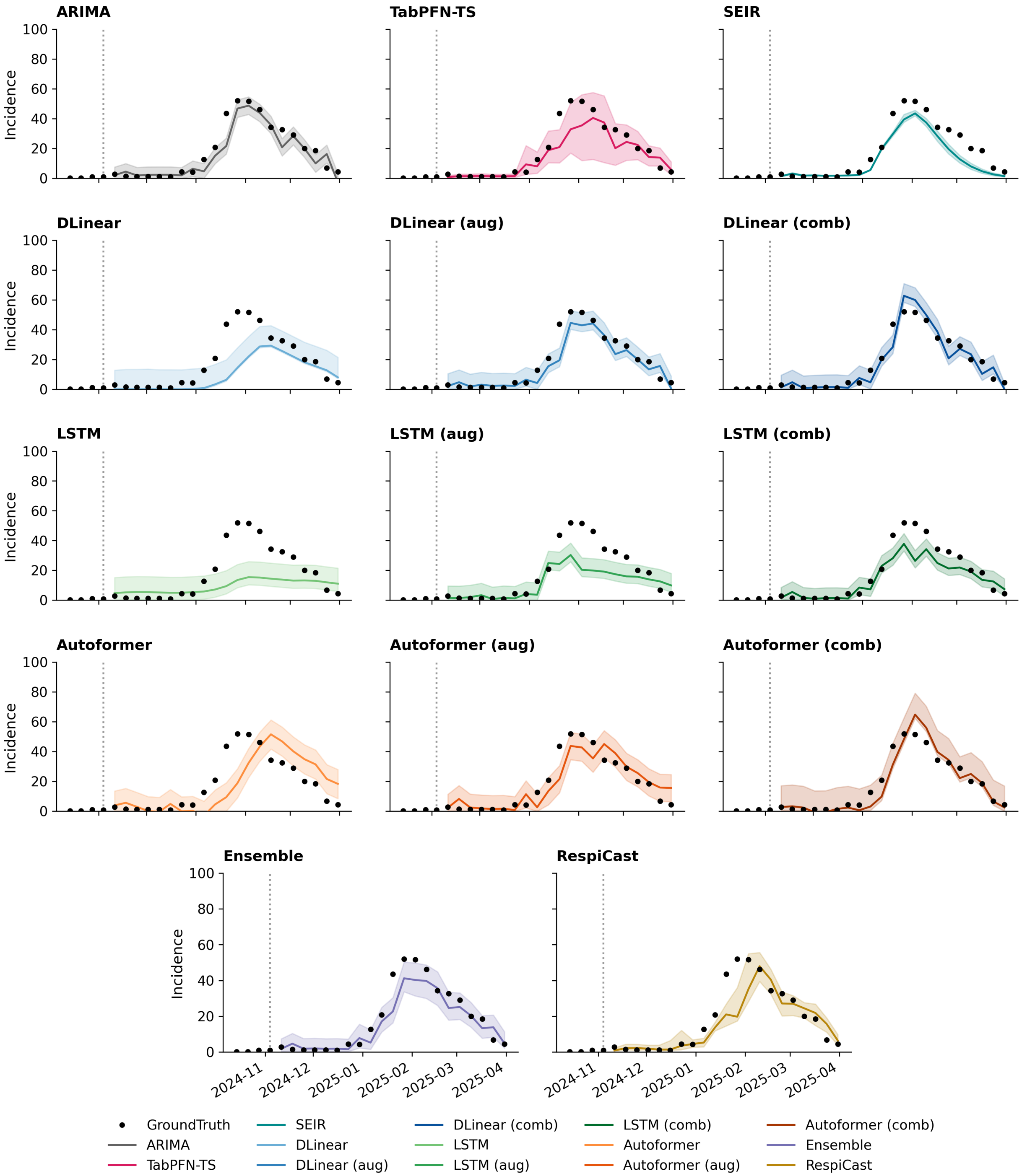


Figure S43: Model forecasts for Romania incidence at forecasting horizon 1. Black dots denote the observed ground truth, coloured lines represent point forecasts, and shaded regions indicate predictive uncertainty.

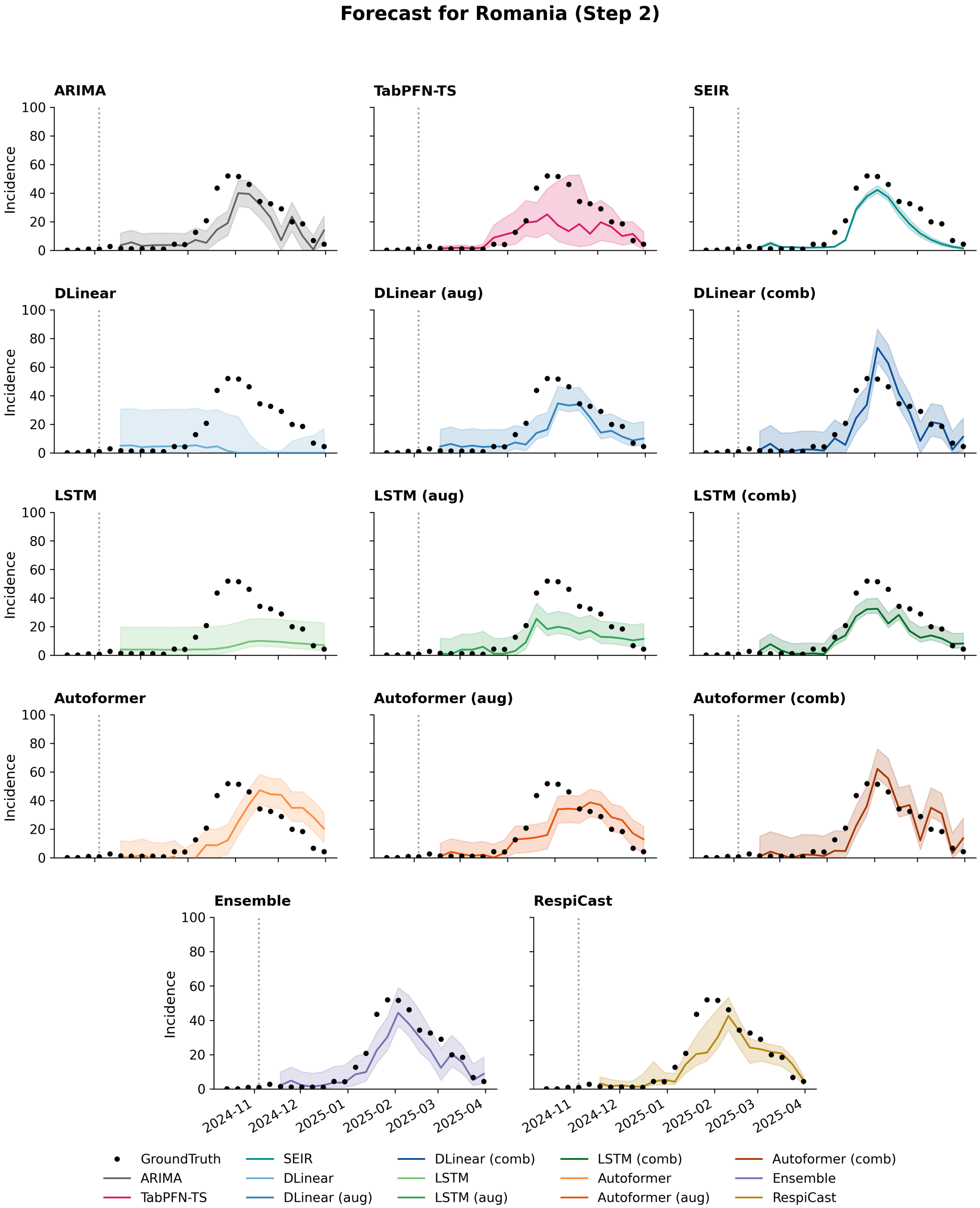


Figure S44: Model forecasts for Romania incidence at forecasting horizon 2. Black dots denote the observed ground truth, coloured lines represent point forecasts, and shaded regions indicate predictive uncertainty.

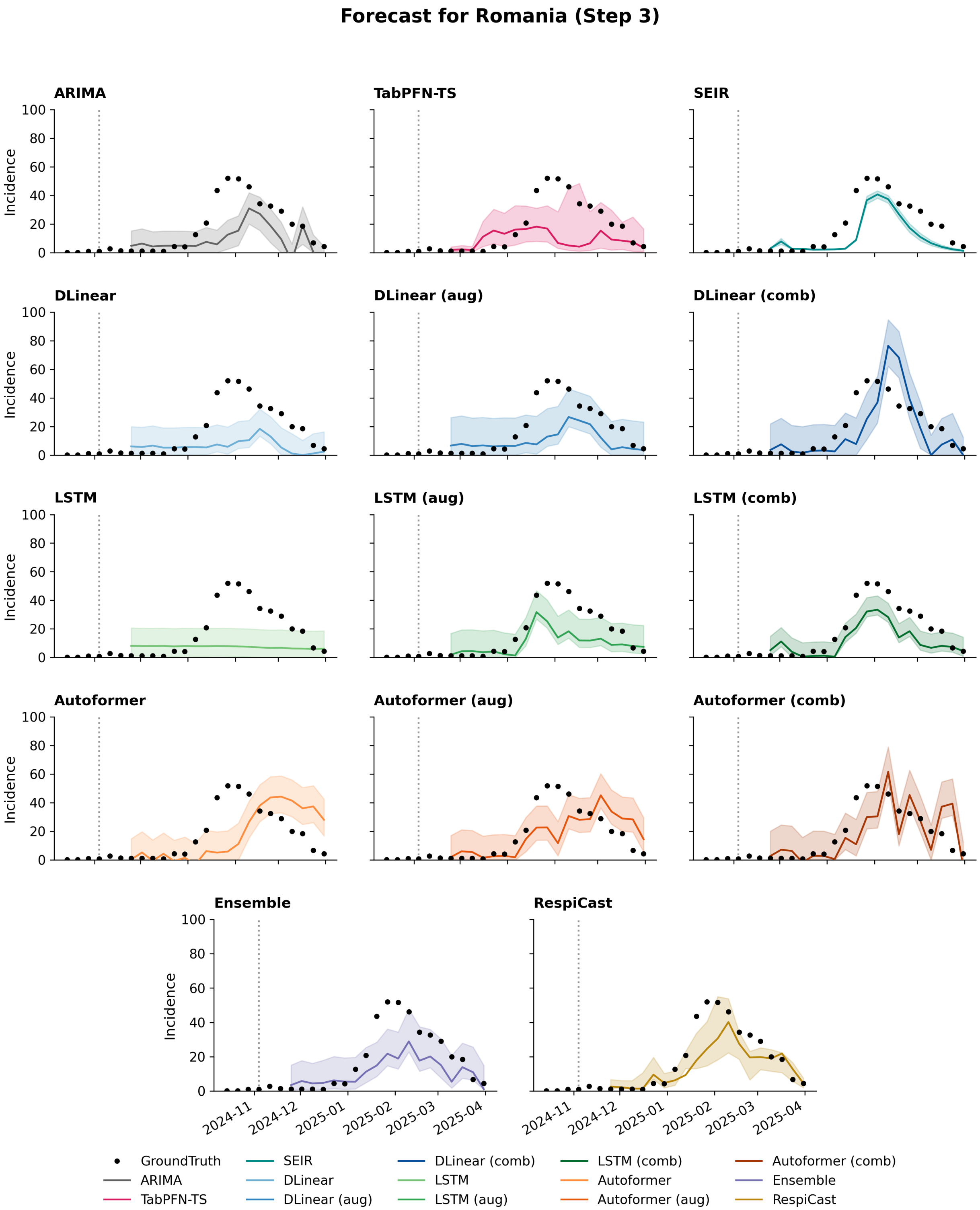


Figure S45: Model forecasts for Romania incidence at forecasting horizon 3. Black dots denote the observed ground truth, coloured lines represent point forecasts, and shaded regions indicate predictive uncertainty.

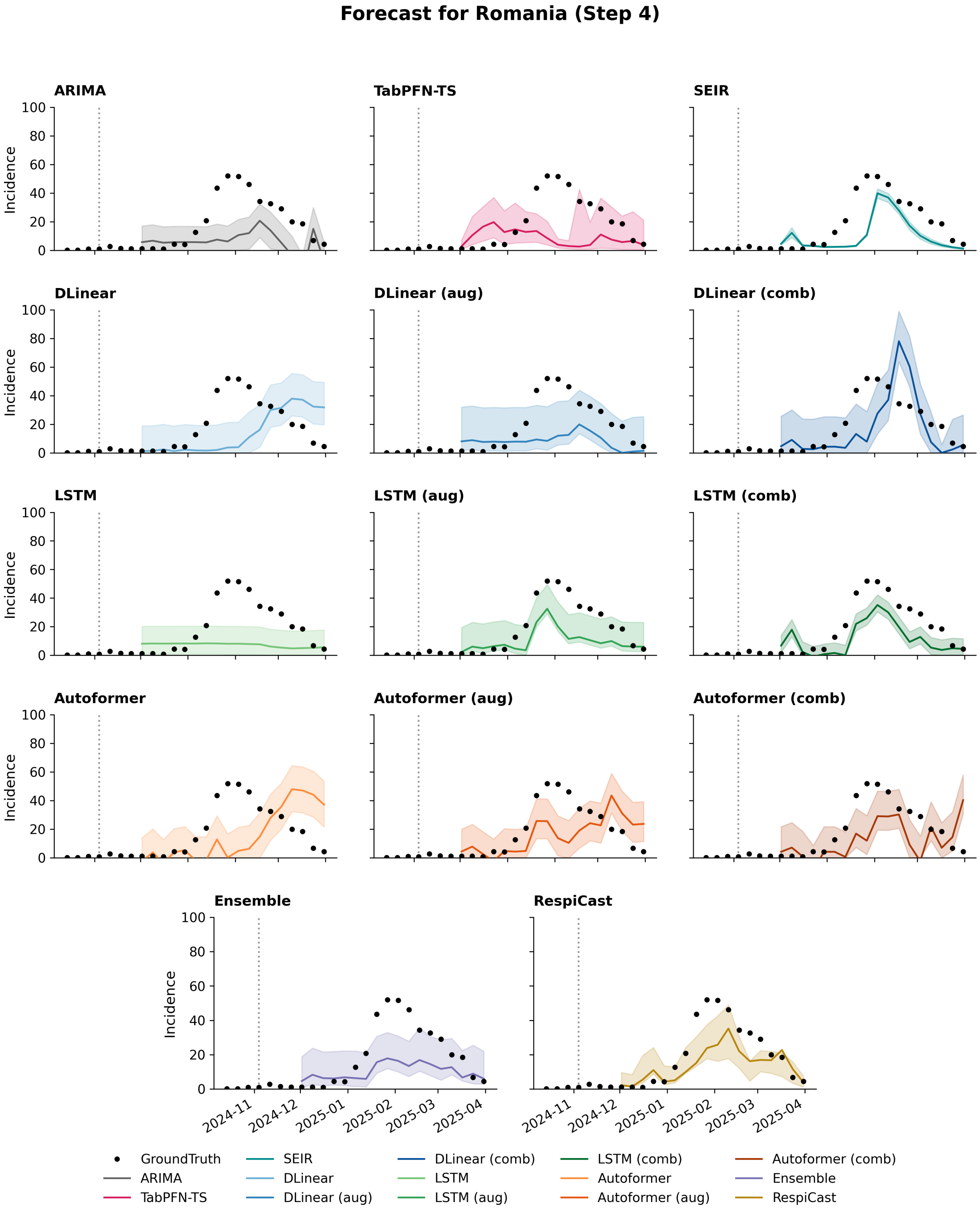


Figure S46: Model forecasts for Romania incidence at forecasting horizon 4. Black dots denote the observed ground truth, coloured lines represent point forecasts, and shaded regions indicate predictive uncertainty.

[11] N Gozzi, C Gioannini, P Milano, I Vismara, L Rossi, M Quaggiotto, S Fiandrino, D Paolotti, A Vespignani, H Johnson, et al. Performance evaluation of respicast ensemble forecasts for primary care syndromic indicators of viral respiratory disease in europe during the 2023/24 winter season. 2025.

[12] Kiesha Prem, Alex R Cook, and Mark Jit. Projecting social contact matrices in 152 countries using contact surveys and demographic data. *PLoS computational biology*, 13(9):e1005697, 2017.

[13] Tina Toni, David Welch, Natalja Strelkowa, Andreas Ipsen, and Michael PH Stumpf. Approximate bayesian computation scheme for parameter inference and model selection in dynamical systems. *Journal of the Royal Society Interface*, 6(31):187–202, 2009.

[14] Yannik Schälte, Emmanuel Klinger, Emad Alamoudi, and Jan Hasenauer. pyabc: Efficient and robust easy-to-use approximate bayesian computation. *Journal of Open Source Software*, 7(74):4304, 2022.

[15] Matthew Biggerstaff, Simon Cauchemez, Carrie Reed, Manoj Gambhir, and Lyn Finelli. Estimates of the reproduction number for seasonal, pandemic, and zoonotic influenza: a systematic review of the literature. *BMC infectious diseases*, 14(1):1–20, 2014.

[16] Elizabeth B White, Lauren Grant, Josephine Mak, Lauren Olsho, Laura J Edwards, Allison Naleway, Jefferey L Burgess, Katherine D Ellingson, Harmony Tyner, Manjusha Gaglani, et al. Influenza vaccine effectiveness against illness and asymptomatic infection in 2022–2023: A prospective cohort study. *Clinical infectious diseases*, 80(4):893–900, 2025.
